# Polygenic regression uncovers trait-relevant cellular contexts through pathway activation transformation of single-cell RNA sequencing data

**DOI:** 10.1101/2023.03.04.23286805

**Authors:** Yunlong Ma, Chunyu Deng, Yijun Zhou, Yaru Zhang, Fei Qiu, Dingping Jiang, Gongwei Zheng, Jingjing Li, Jianwei Shuai, Yan Zhang, Jian Yang, Jianzhong Su

**Affiliations:** School of Biomedical Engineering, School of Ophthalmology & Optometry and Eye Hospital, Wenzhou Medical University, Wenzhou, 325027, China; Oujiang Laboratory, Zhejiang Lab for Regenerative Medicine, Vision and Brain Health, Wenzhou 325101, Zhejiang, China; School of Life Science and Technology, Harbin Institute of Technology, Harbin 150080, China; School of Life Sciences, Westlake University, Hangzhou, 310012, Zhejiang, China; Westlake Laboratory of Life Sciences and Biomedicine, Hangzhou, Zhejiang 310024, China

**Keywords:** GWAS, Single-cell RNA sequencing, Genetic variants, Risk genes, Trait-relevant cell types

## Abstract

Advances in single-cell RNA sequencing (scRNA-seq) techniques have accelerated functional interpretation of disease-associated variants discovered from genome-wide association studies (GWASs). However, identification of trait-relevant cell populations is often impeded by inherent technical noise and high sparsity in scRNA-seq data. Here, we developed scPagwas, a computational approach that uncovers trait-relevant cellular context by integrating pathway activation transformation of scRNA-seq data and GWAS summary statistics. scPagwas effectively prioritizes trait-relevant genes, which facilitates identification of trait-relevant cell types/populations with high accuracy in extensive simulated and real datasets. Cellular-level association results identified a novel subpopulation of naïve CD8+ T cells related to COVID-19 severity, and oligodendrocyte progenitor cell and microglia subsets with critical pathways by which genetic variants influence Alzheimer’s disease. Overall, our approach provides new insights for the discovery of trait-relevant cell types and improves the mechanistic understanding of disease variants from a pathway perspective.

## Introduction

Genome-wide association study (GWAS) data on complex diseases and numerous genotype-phenotype associations have tremendously accumulated in the past decades^1-4^. However, functional interpretation of these variants identified by GWASs remains challenging. It is still unclear how these variants regulate key biological pathways in relevant tissues/cell types to mediate disease development. The advent of single-cell RNA sequencing (scRNA-seq) technology has provided an unprecedented opportunity to characterize cell populations and states from heterogeneous tissues^5^. Unveiling trait-relevant cell populations from scRNA-seq data is crucial for exploring the mechanistic etiology of complex traits (including diseases)^6^. Thus, linking scRNA-seq data with genotype-phenotype association information from GWASs has considerable potential to provide new insights into the polygenic architecture of complex traits at a high resolution^7^.

Several studies have revealed significant enrichment of complex traits in relevant tissue types by integrating tissue-specific gene expression profiles with GWAS summary statistics^8-10^. Inspired by these tissue-type enrichment methods, several methods^11-15^, including LDSC-SEG, RolyPoly, and MAGMA-based approaches, have been employed to incorporate GWAS and scRNA-seq data to identify predefined cell types associated with complex traits. However, these approaches largely neglect the considerable intra-heterogeneity within each cell type and thus are not suitable for making inferences at single-cell resolution. Recently, scDRS^16^ was developed to distinguish disease-associated cell populations at the single-cell level; however, its accuracy relies heavily on a set of disease-specific genes identified from GWAS data using gene-based association test methods^17-20^, such as MAGMA^20^. Although the gene-scoring methods focus on the top significant genotype-phenotype associations and have been applied to bulk tissue or aggregated data analysis, it is still challenging to use these methods to make accurate per-cell-based inferences in scRNA-seq data. The top genetic association signals at specific loci may be absent from most cells because of the extensive sparsity and technical noise in single cell data^21,22^. To date, no method exists to optimize effective and robust trait-relevant genes applicable to scRNA-seq data for accurate inference of disease-associated cells at a fine-grained resolution.

Dynamic cell activities and states are often caused by the combined actions of interacting genes in a given pathway or biological process^23^. Compared with leveraging the expression level of individual genes, pathway activity scoring methods that collapse the functional actions of different genes involved in the same biological pathways can prominently enhance statistical power and biological interpretation for determining particular cellular functions or states^21,24-26^. Recent studies have shown that such pathway-based scoring methods exhibit a greater reduction of technical and biological confounders of scRNA-seq data^27-29^. Moreover, multiple lines of evidence have suggested that clinically informative variants associated with complex diseases mainly occur in systems of closely interacting genes, and even variants with weak association signals clustered in the same biological pathway could provide critical information to understand the genetic basis of complex diseases^30,31^. Thus, integrating coordinated transcriptional features in biological pathways from scRNA-seq data and polygenic risk signals from GWAS summary statistics is a promising approach to prioritize trait-relevant genes and distinguish critical cells by which genetic variants influence diseases.

Here, we present a pathway-based polygenic regression method (scPagwas) that integrates scRNA-seq and GWAS data for the discovery of cellular context critical for complex diseases and traits. scPagwas performs a linear regression of GWAS signals on pathway activation transformed from scRNA-seq data to identify a set of trait-relevant genes, which are subsequently used to infer the most trait-relevant cell subpopulations. We show that scPagwas outperforms the state-of-the-art methods using extensive simulated and real scRNA-seq datasets. Through scPagwas-based analyses of different diseases, we provide new biological insights into how disease-associated naïve CD8+ T cells are involved in COVID-19 severity and subsets of microglia and oligodendrocyte progenitor cells (OPCs) can contribute to Alzheimer’s disease (AD) risk.

## Results

### Overview of scPagwas

Given extensive evidence^11,13,32,33^ indicating a positive correlation between genetic associations for a trait of interest and expression levels of genes in bulk tissue or specific cell type, we apply this principle to scRNA-seq data and take advantage of gene expression signatures shared in a biological pathway. scPagwas first links single-nucleotide polymorphisms (SNPs) in the GWAS summary data to each pathway by annotating SNPs to their proximal genes of the corresponding pathway (Figure 1A). In each cell, scPagwas calculates the correlation between the genetic effects of SNPs and gene expression levels within a given pathway to estimate regression coefficient *τ* (Figure 1B), which reflects the strength of association between pathway-specific gene expression activity and the variance of SNP effects^34^. Meanwhile, scPagwas transforms the normalized gene-by-cell matrix to a pathway activity score (PAS)-by-cell matrix, which is constructed using the first principal component (PC1) of gene expression in each pathway via the singular value decomposition (SVD) method^22,35^ (Figure 1C, see Methods).

**Figure 1.**
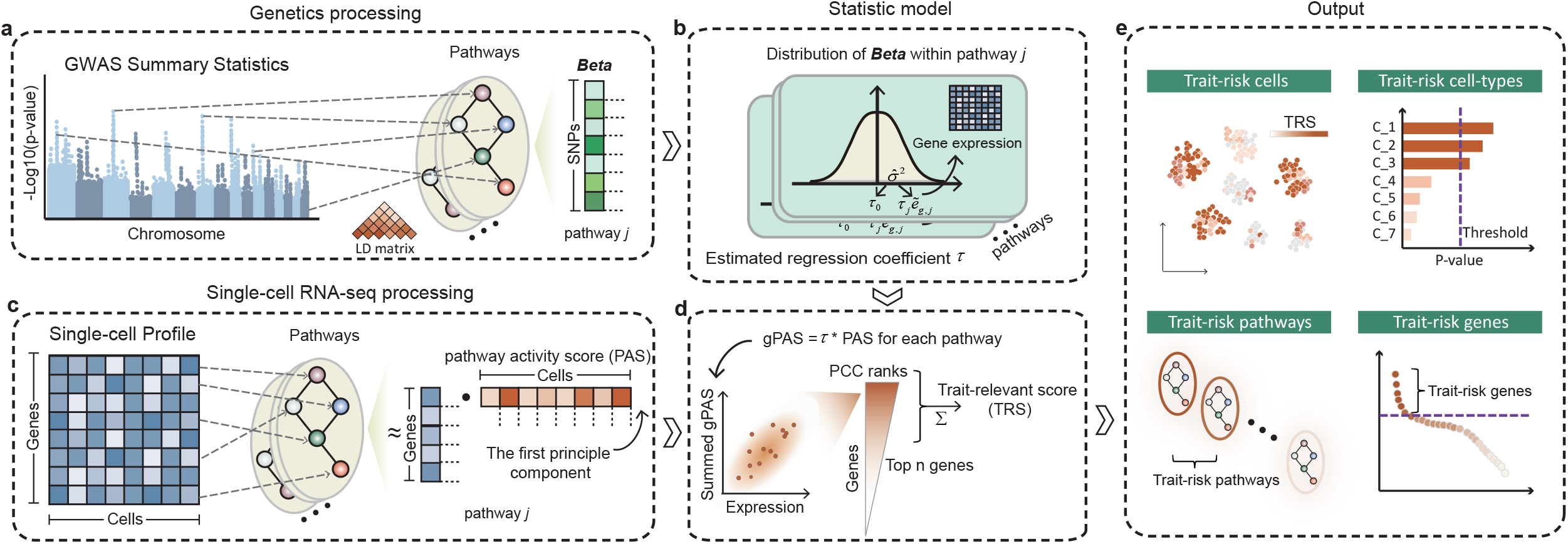
Overview of scPagwas approach. A. Linking single-nucleotide polymorphisms (SNPs) from GWAS summary statistics into corresponding pathway. The linkage disequilibrium (LD) matrix for SNPs is calculated based on the 1,000 Genomes Project Phase 3 Panel. B. Statistical model. The pathway-specific polygenic regression analysis between the SNP effect sizes and the adjusted gene expression within a given pathway is used to infer an estimated coefficient *τ* for each pathway. C. Transforming gene-by-cell matrix to pathway activity score (PAS)-by-cell matrix via using the singular value decomposition (SVD) method. The first principal component (PC1) represents the PAS for each pathway. D. The Pearson correlation model. The genetically-associated PAS (gPAS) for each pathway is defined as the product between the estimated coefficient *τ* and weighted PAS (see Methods). The bottom panel represents the Pearson correlation analysis of the summed gPASs of all pathways in a given cell with the expression of a given gene for all individual cells, and ranking the Pearson correlation coefficients (PCCs) to prioritize top trait-relevant genes. Then, scPagwas uses the cell-scoring method in Seurat to collapse the expression of top *n* trait-relevant genes (default top 1,000 genes) for calculating the trait-relevant score (TRS) of each cell. E. scPagwas outputs. The typical outputs includes (i) trait-relevant cells, (ii) trait-relevant cell types, and (iii) trait-relevant pathways/genes.

Following previous studies^13,34^, we compute the product of 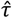 and PAS for each pathway, hereinafter referred to as genetically-associated PAS (gPAS), to capture the pathway-based genetic variances of traits of interest at the single-cell level (Figure 1D). Then, we use the central limit theorem method^36^ to identify significant trait-relevant pathways based on the ranking of gPASs of pathways across individual cells within each cell type (see Supplementary Methods). In the meanwhile, we compute the sum of gPASs over all pathways in each cell, correlate it with the expression level of each gene across cells, and prioritize the trait-relevant genes by ranking the correlations (Figure 1D). Finally, a trait-relevant score (TRS) of each cell is computed by averaging the expression level of the trait-relevant genes and subtracting the random control cell score via the cell-scoring method used in Seurat^37^ (see Methods). By treating the set of cells in a predefined cell type as a pseudo-bulk transcriptomic profile, scPagwas can also be employed to infer significant trait-relevant cell-types using the block bootstrap method.

The input of scPagwas includes gene sets of pathways, a gene-by-cell matrix of scRNA-seq data, and summary statistics from a GWAS or meta-analysis for a quantitative trait or disease (case-control study). The typical output includes 1) per cell-based TRSs and the corresponding P values; 2) trait-associated cell types from the block bootstrap analysis; 3) trait-relevant pathways based on the ranked gPASs; 4) trait-relevant genes based on the ranked PCCs (Figure 1E).

### scPagwas effectively identifies trait-relevant genes

Because trait-relevant genes are vital for inferring the TRS of each cell, we compared the biological functions of the top 1,000 trait-relevant genes identified from scPagwas with those identified with the widely-used gene-scoring method, MAGMA^20^, and three other well-established eQTL-based methods including TWAS^17^, S-PrediXcan^19^, and S-MultiXcan^18^. A panel of highly-heritable hematopoietic traits was used for benchmark analysis (Supplementary Tables S1-S2). We found that trait-relevant genes identified by scPagwas were more highly enriched in functional gene sets related to blood cell traits than those identified by the other four gene-scoring methods (Figure 2A and Supplementary Table S3). For example, the lymphocyte count-relevant genes prioritized by scPagwas showed highly significant enrichment in biological processes related to immune response, including T cell activation, adaptive immune response, leukocyte differentiation, and leukocyte cell-cell adhesion, whereas those prioritized by MAGMA lacked enrichment in any functional term (false discovery rate, FDR < 0.01, Figure 2B).

**Figure 2.**
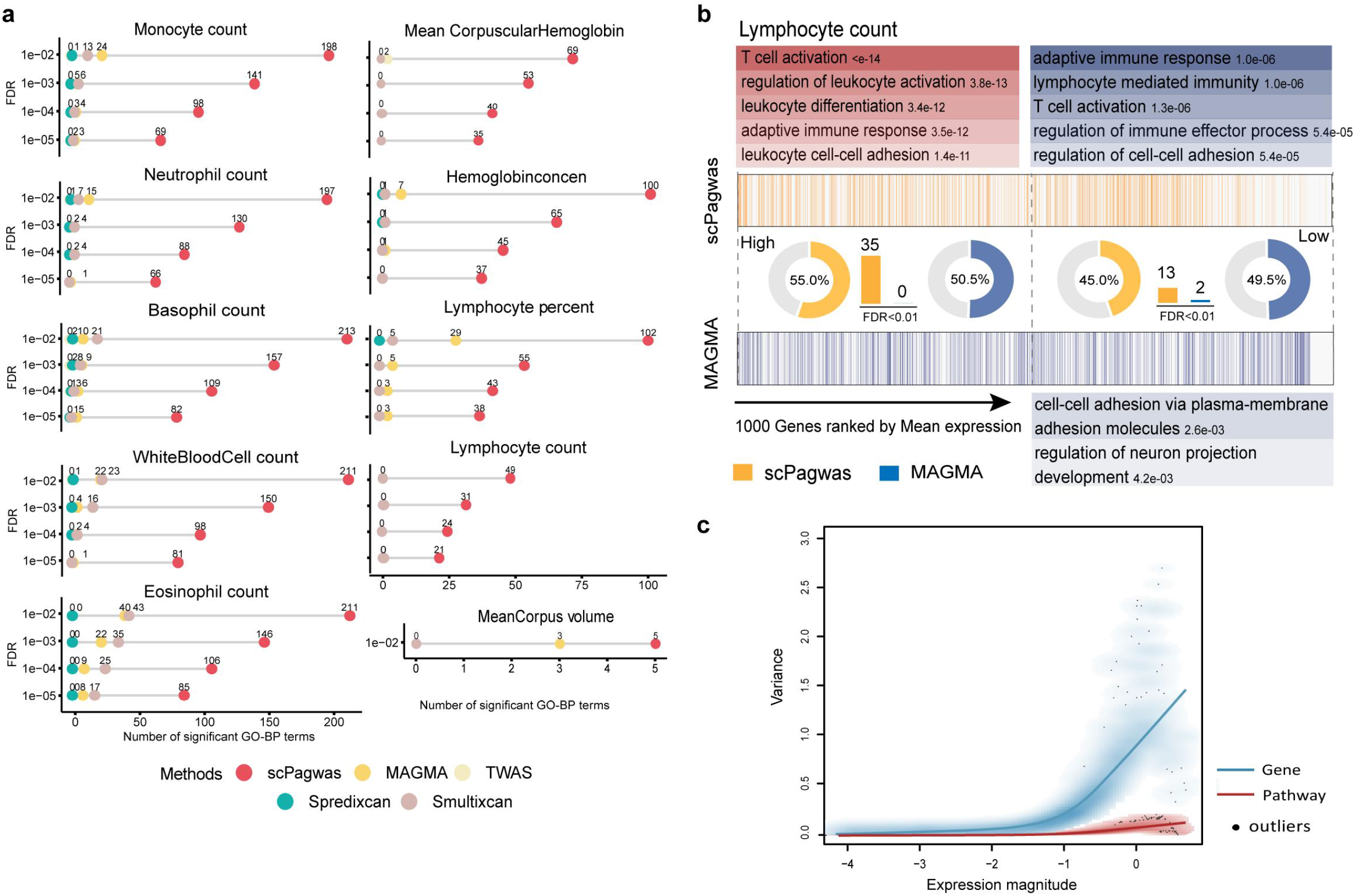
The reproducible and functional results of scPagwas. A. GO-term enrichment analyses of top-ranked 1,000 genes from scPagwas and other four gene-based methods (i.e., MAGMA, TWAS, S-PrediXcan, and S-MultiXcan) for 10 highly-heritable blood cell traits. Different color dots represent number of significant GO-terms of biological processes (BP, FDR < 0.01) enriched by top-ranked genes from scPagwas and other four methods (see Supplementary Table S3). B. Example of the distribution of scPagwas-identified risk genes and MAGMA-identified risk genes ranked by their average expression across all cells for the lymphocyte count trait. The percentages of risk genes for top-half (over-expression genes) and bottom-half (down-expression genes) cells are shown in the plot accordingly. GO enrichment results of the lymphocyte count trait classified by two groups of over-expression genes (FDR < 0.01, 35 significant GO-terms enriched by scPagwas vs. 0 GO-terms by MAGMA) and down-expression genes (FDR < 0.01, 13 significant GO-terms enriched by scPagwas vs. 2 GO-terms by MAGMA) are shown in the plot. C. Plot demonstrating the variance of gene-level expression magnitude and pathway-level expression magnitude in the BMMC scRNA-seq dataset (n = 35,582 cells). Fitted line with red color represents pathway-level expression magnitude, which shows a mean-variance fit that demonstrates the relationship between average expression of genes in a given pathway (x axis) and its corresponding variance (y axis). Fitted line with blue color represents gene-level expression magnitude, which shows a mean-variance fit that demonstrates the relationship between average gene expression (x axis) and gene variance (y axis). The black dots in the plot indicate outliers. See also Supplementary Figure S2.

In scRNA-seq data, the sparsity and technical noise of individual genes can lead to high computational costs and inadequate association inference at single-cell level^21,22^. Using five distinct scRNA-seq datasets, we found that the use of pathway information with scPagwas could remarkably reduce the sparsity compared with that of individual gene-based evaluations (Supplementary Figure S1). The average expression magnitudes of individual genes showed a strong positive correlation with their variances (Figure 2C, blue line). In contrast, pathway activation scores transformed from transcriptome profiles significantly reduced the technical noise of variances of single-cell data, which facilitated the identification of biologically-relevant genes and improved downstream analyses^38^ (Figure 2C and Supplementary Figure S2). These results demonstrate that scPagwas not only reduces sparsity and technical noise but also prioritizes more functional genes associated with trait of interest for per-cell-based inference to identify trait-relevant cells.

### Assessment of scPagwas in discerning trait-relevant cells

We first assessed the power and precision of scPagwas in identifying trait-relevant cells using a real GWAS dataset and simulated scRNA-seq datasets. We adopted a highly heritable and relatively simple trait, monocyte count, for benchmark analysis, with the GWAS summary statistics from a large-scale study (N = 563,946, Supplementary Table S1). We synthesized an scRNA-seq dataset from fluorescence-activated cell-sorted bulk hematopoietic populations as the ground truth (see Methods), which contained a known relevant cell type (monocytes, n = 1,000 cells) and non-relevant cell types (T and B cells, dendritic cells (DCs), and natural killer (NK) cells, n = 1,000 cells in total, Figure 3A). We found that scPagwas (using the cell-scoring method of Seurat^37^ by default) could accurately distinguish monocyte count-relevant cells from all simulated cells (precision = 95.9%, Figure 3B). We further examined whether the scPagwas-identified trait-relevant genes could improve the power of the latest cell-scoring method, scDRS^16^, by comparing the results with those using the default gene-based method, MAGMA. The precision of scDRS in identifying the trait-relevant cells increased from 0.940 when using the the top 1,000 genes prioritized by MAGMA to 0.957 when using the top 1,000 genes prioritized by scPagwas (Figure 3C-D).

**Figure 3.**
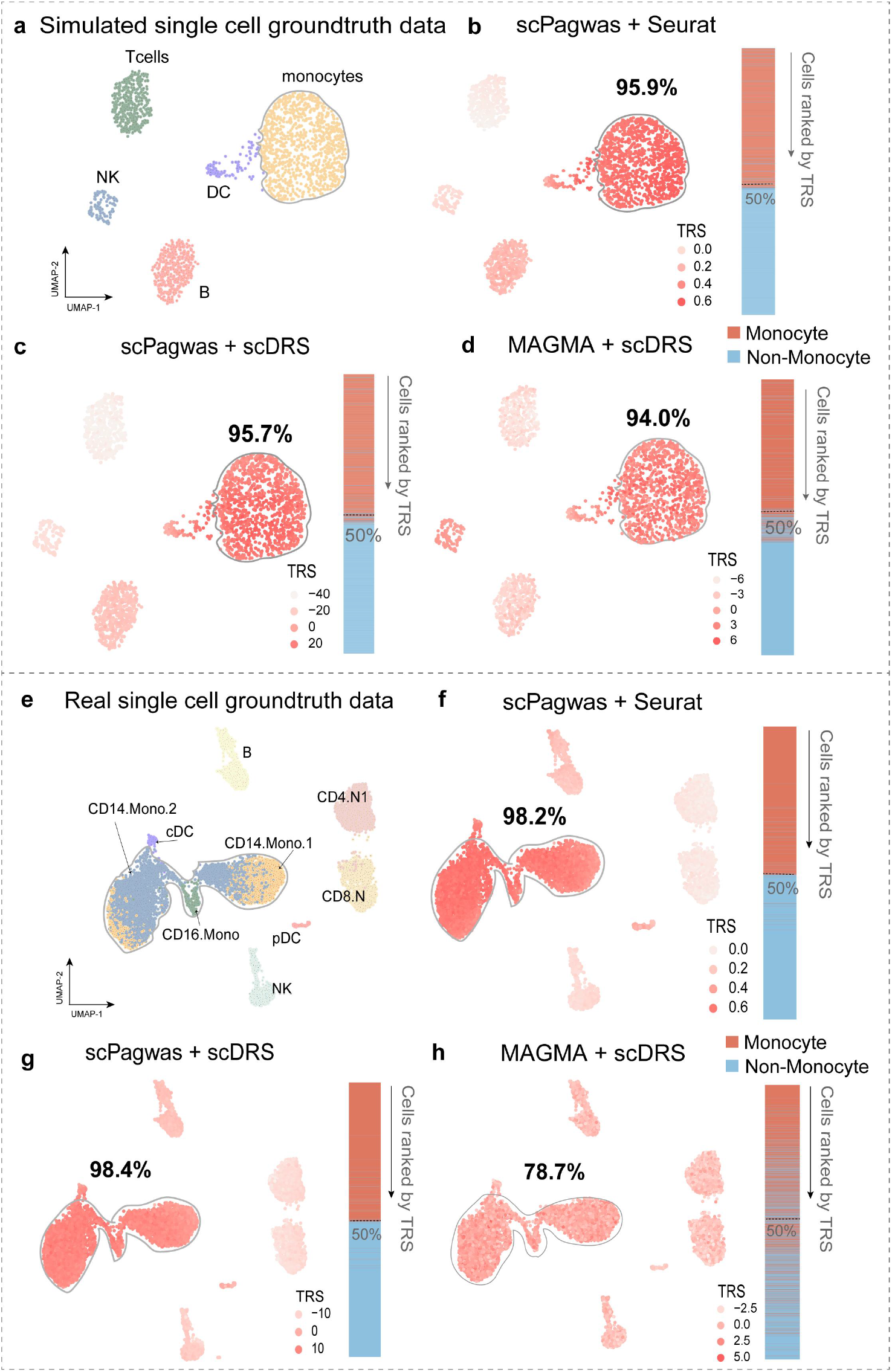
Assessment of the performance of scPagwas in both simulated and real scRNA-seq datasets. A. UMAP embedding plot shows the cellular component of a synthesized ground truth scRNA-seq dataset (monocytes: n = 1,000 cells, and T, B, DC, and NK: n = 1,000 cells in total). E. UMAP plot shows the cellular component of a real ground truth scRNA-seq dataset. The real BMMC scRNA-seq dataset contains 10,000 cells with seven cell types: monocytes (11_CD14.Mono.1, 12_CD14.Mono.2, and 13_CD16.Mono, n = 5,000 cells), DC (09_pDC and 10_cDC, n = 200 cells), T cells (19_CD8.N and 20_CD4.N1, n = 3,000 cells), B cells (17_B, n = 1,000 cells), and NK cells (25_NK, n = 800 cells). B, C, F, G) Illustration of the performance of top 1,000 scPagwas-identified genes for identifying monocyte count trait-relevant cells based on two cell-scoring methods (i.e., Seurat and scDRS) in the synthesized (B, C) and real (F, G) scRNA-seq datasets. D, H). Illustration of the performance of top-ranked 1,000 putative disease genes identified by the gene-scoring method of MAGMA for identifying monocyte count trait-relevant cells based on the scDRS method in the synthesized (D) and real (H) scRNA-seq datasets. The UMAP projections of every cell colored by its TRS. The vertical bar exhibits cells descendingly ranked according to their corresponding TRSs (top-ranked 1,000 genes), where red color indicates monocyte cells and blue color indicates non-monocyte cells. The accuracy of each method represents the percentage of monocyte count trait-related cells (i.e., monocytes) for top-half cells that are ranked by TRS for all cells in a descending manner. See also Supplementary Figures S3-S4.

Moreover, compared with the gene-based methods that incorporate eQTL information (i.e., S-MultiXcan, S-PrediXcan, and TWAS), the scPagwas-identified trait-relevant genes considerably enhanced the performance of the scDRS in distinguishing cells relevant to the monocyte count trait (scPagwas precision = 95.7% vs other three methods precision = 62.9%–90%, Supplementary Figure S3A). Using the same simulated single-cell dataset, we further examined the lymphocyte count trait also with a large-scale GWAS dataset (N = 171,643 samples) to benchmark the performance of scPagwas against the other four gene-based methods when using scDRS to score cells. We observed a consistent result that scPagwas yielded the best performance (scPagwas precision = 81.8% vs other four methods precision < 50%, Supplementary Figure S4A). In addition, we evaluated the performance of scPagwas in identifying predefined cell types related to a trait of interest in simulated data (see Supplementary Methods). We observed that scPagwas could effectively identify trait-relevant cell types under different genetic architectures when the number of included pathways was more than 100 (Supplementary Figures S5–S6).

Next, we assessed whether scPagwas could distinguish monocyte count trait-related enrichment in a real ground truth scRNA-seq dataset (Figure 3E) that contained monocytes (CD14+ and CD16+ monocytes, n = 5,000 cells) and non-monocyte cells (T cells (n = 3,000), B cells (n = 1,000), DCs (n = 200), and NK cells (n = 800)) from a bone marrow mononuclear cell (BMMC) scRNA-seq dataset (Supplementary Table S2)^39^. Consistent with the simulation results, scPagwas robustly identified the known trait-relevant cell populations with higher precision using Seurat as the cell-scoring method (precision = 98.2%, Figure 3F). Compared with the default setting of scDRS that uses the top 1,000 MAGMA-identified genes, applying the top 1,000 scPagwas-identified genes to scDRS considerably enhanced the discovery of monocyte count-relevant cells with the improvement of the precision from 0.787 to 0.984 (Figure 3G–H).

Analogous to the simulation results, we only found a moderate enrichment of monocyte count-relevant cells by applying the top genes prioritized by the three eQTL-based methods (S-MultiXcan, S-PrediXcan, and TWAS) to scDRS analysis (precision = 61.7–71.0%, Supplementary Figure S3B). This observation remained reproducible for lymphocyte count with the inclusion of the same real ground truth scRNA-seq dataset (Supplementary Figure S4B). When further evaluating whether the number of included top trait-relevant genes influences the power of scoring trait-relevant cells, scPagwas using the scDRS method achieved a stable and robust performance after choosing the top 100 trait-relevant genes. In contrast, the use of scDRS with MAGMA resulted in a variable and moderate performance that was largely influenced by the number of included genes prioritized by MAGMA or the three other eQTL-based methods (Supplementary Figure S7A–B). Additionally, when applying genes prioritized by scPagwas to two other cell-scoring methods, e.g., Vision^29^ and AUCell^27^, we consistently found that these cell-scoring methods yielded a high performance in identifying cells relevant to monocyte count with either the simulated or real RNA-seq data (Supplementary Figure S3C-D). Taken together, our results reveal that scPagwas enables trait relevance to be accurately and robustly characterized at the single-cell resolution.

### scPagwas accurately identifies blood cell trait-relevant cell populations at distinct stages of human hematopoiesis

scPagwas was used to identify hematological trait-relevant cell populations in a large BMMC scRNA-seq dataset (n = 35,582 cells, Figure 4A) that contained the full spectrum of human hematopoietic differentiation from stem cells to their progeny^39^. To explore the genetic associations for 10 highly-heritable blood cell traits in various cellular contexts, we aggregated the TRSs of individual cells within the same annotated cell type to assess the enrichments of hematopoietic traits at distinct stages of human hematopoiesis using the unsupervised clustering method (Supplementary Table S1). According to the aggregation results, different cell populations from the same lineage were predisposed to have consistent associations across relevant traits (Figure 4B and Supplementary Figure S8A). For example, red blood cell traits, including hemoglobin concentration, mean corpuscular hemoglobin, and mean corpus volume, tended to have similar associations within the same module based on the TRS of the cell type, consistent with previous findings^22^.

**Figure 4.**
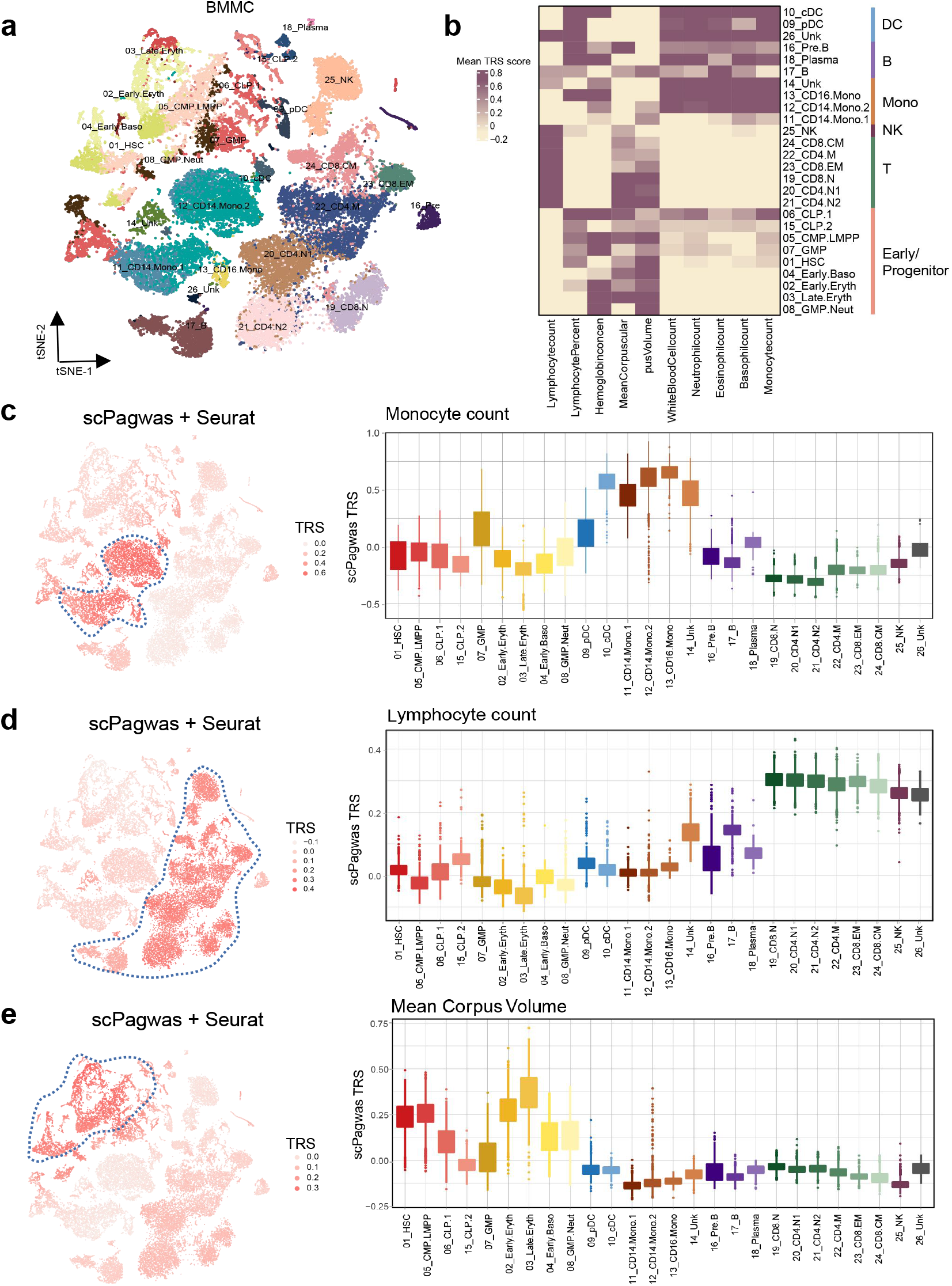
Application of scPagwas to multiple blood cell traits for identifying trait-relevant cells. The 10 hematological traits were analyzed using scPagwas (Seurat) on a large BMMC scRNA-seq dataset. A. The tSNE plot shows the cell type labels. B. The average TRSs for cells belonging to the same cell type are shown in the heatmap. Unsupervised clustering analysis was conducted and cell-type category were grouped into six main clusters, including DCs, B cells, Monocytes (Mono), NKs, T cells, and early/progenitor cells. cDC, classical dendritic cell; pDC, plasmacytoid dendritic cell; Unk, Unkown; CD4.M, CD4+ memory T cells; CD8.CM, CD8+central memory T cells; CD8.EM, CD8+effector memory T cells; CD8.N, CD8+ naïve T cells; CD4.N1/N2, CD4+naïve T cells; CLP, common lymphoid progenitor; CMP, common myeloid progenitor; GMP, granulocyte-macrophage progenitor; LMPP, lymphoid-primed multipotent progenitor; HSC, Hematopoietic stem cell; Baso, basophil; Eryth, erythrocyte; Neut, neutrophil. C-E) Per-cell TRS calculated by scPagwas (Seurat) for three representative traits including monocyte count (C), lymphocyte count (D), and mean corpus volume (E) are shown in tSNE coordinates (left) and per cell type (right). Boxplots (left to right: n = 1,425, 2,260, 903, 377, 2,097, 1,653, 446, 111, 1,050, 544, 325, 1,800, 4,222, 292, 420, 710, 1,711, 62, 1521, 2,470, 2,364, 3,539, 796, 2,080, 2,143, and 161 cells) show the median with interquartile range (IQR) (25-75%); whiskers extend 1.5× the IQR. See also Supplemenatry Figure S9.

The TRSs of cells for three representative traits are shown in low-dimensional t-distributed stochastic neighbor embedding (t-SNE) space (Figure 4C-E). Remarkably, cell lineages relevant to corresponding blood cell traits yielded considerably high TRSs under different conditions (Figure 4C–E and Supplementary Figure S8B-D), indicating that the cell specificity of these genetic effects was well captured by scPagwas. For monocyte count, scPagwas identified not only monocyte-related cell compartments with increased TRSs but also granulocyte-monocyte progenitor cells showing increased enrichment (Figure 4C). Furthermore, several cell compartments related to CD8+ T cells, CD4+ T cells, NK cells, and B cells yielded increased TRSs for lymphocyte count (Figure 4D), and early and late erythrocytes, CMP-LMMP, and hematopoietic stem cells exhibited increased TRSs for the mean corpus volume (Figure 4E).

When applying the top 1,000 scPagwas-prioritized genes to scDRS in the BMMC scRNA-seq dataset, cells relevant to three representative traits were enriched and had increased TRSs (Supplementary Figure S9A–C, left panel). However, the use of scDRS with the top 1,000 MAGMA-prioritized genes did not show such trait-relevant enrichment (Supplementary Figure S9A–C, right panel). Using an independent peripheral blood mononuclear cell (PBMC) scRNA-seq dataset with a larger number of cells (n = 97,039 cells)^40^, consistently, scPagwas using either cell scoring method, Seurat or scDRS, accurately distinguished monocyte and lymphocyte count-relevant cell compartments, whereas there was no specific trait relevance using scDRS with the top MAGMA-prioritized genes (Supplementary Figures S10–S11). Collectively, these results suggest that scPagwas can recapitulate known associations between blood cell traits and the cellular context and identify novel trait-associated cell subpopulations and states.

### scPagwas identifies novel immune subpopulations associated with severe COVID-19 risk

Understanding the effects of host genetic factors on immune responses to severe infection can contribute to the development of effective vaccines and therapeutics to control the COVID-19 pandemic. scPagwas was applied to discern COVID-19-associated immune cell types/subpopulations by integrating a large-scale GWAS summary dataset on severe COVID-19 (N = 7,885 cases and 961,804 controls) with a large PBMC scRNA-seq dataset (n = 469,453 cells) containing healthy controls and COVID-19 patients with various clinical severities (Supplementary Tables S1–S2). scPagwas identified that three immune cell types, including naïve CD8+ T cells (P = 4.6 × 10^−17^), megakaryocytes (P = 7.8 × 10^−6^), and CD16+ monocytes (P = 1.14 × 10^−4^), demonstrated significant associations with severe COVID-19 (FDR < 0.05, Figure 5A–D and Supplementary Table S4), whereas these three cell types only showed suggestive asssociations inferred by the three cell type-level inference methods (LDSC-SEG^11^, MAGMA-based approach^41^, and RolyPoly^13^) (see Supplementary Methods). Both CD16+ monocytes and megakaryocytes have been reported to be associated with aggressive cytokine storm among severe COVID-19 patients^42,43^.

**Figure 5.**
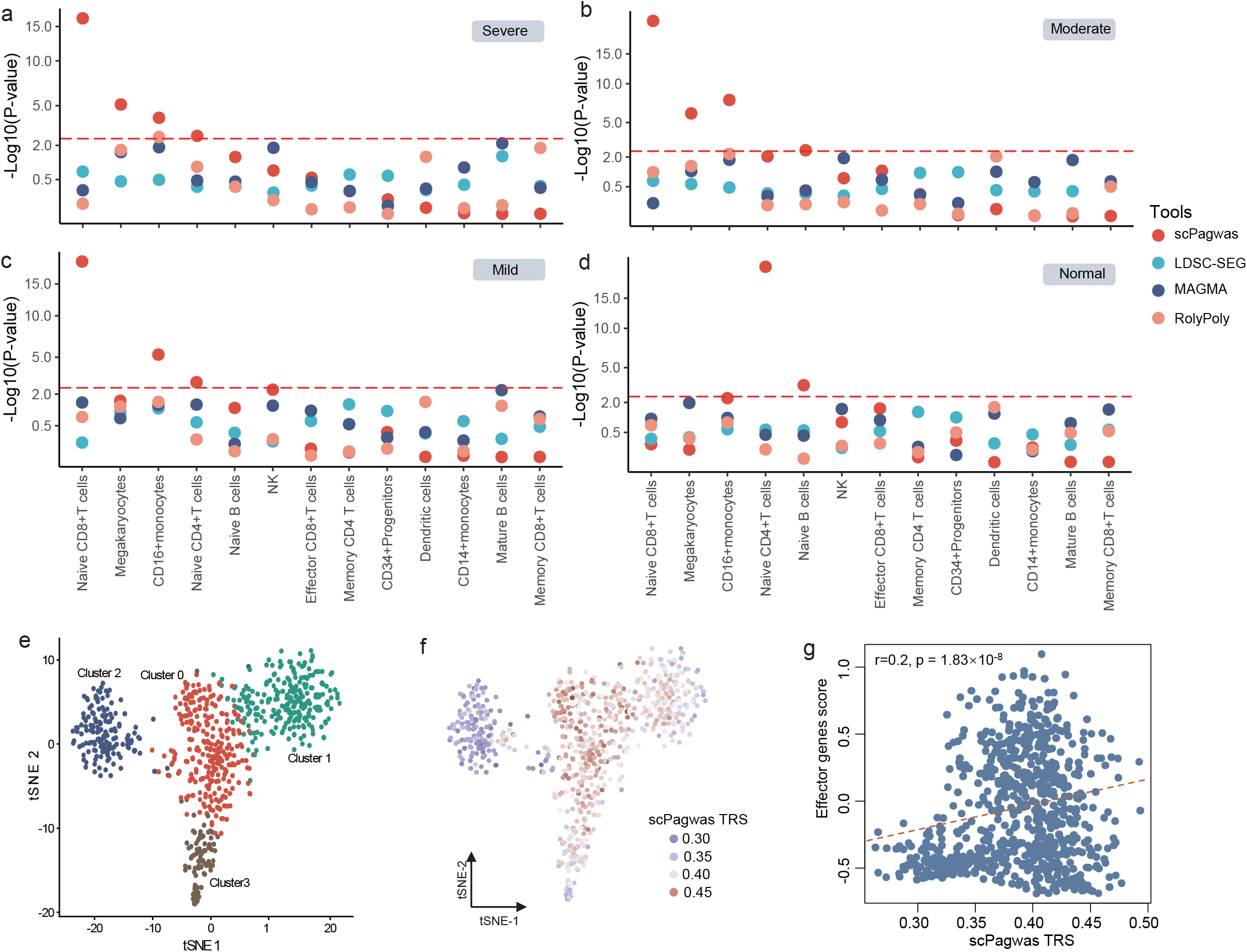
scPagwas identifies trait-relevant immune cell types and subpopulations for severe COVID-19. A-D. Benchmarking analysis of uncovering trait-relevant cell types by using scPagwas, LDSC-SEG, MAGMA-based approach, and RolyPoly for COVID-19 patients with various clinical severities of severe (A), moderate (B), mild (C), and healthy controls (D), respectively. The horizontal dashed red lines represent the significant threshold (Bonferroni corrected P < 0.05), and the horizontal dashed blue lines indicate the raw significant threshold (i.e., raw P < 0.05). E. The tSNE visualization of 766 naïve CD8+ T cells with four cell clusters. F. scPagwas TRS for the phenotype of severe COVID-19 risk is displayed for all naïve CD8+T cells in the tSNE plot. G. The correlation of scPagwas TRSs with molecular signature scores of effector marker genes across all naïve CD8+T cells. See also Supplementary Figures S12-S14.

Of note, scPagwas identified a novel cell subpopulation of naïve CD8+ T cells related to severe COVID-19 (Figure 5A–E). scPagwas identified that five biological pathways relevant to COVID-19 severities showed high specificity for naïve CD8+ T cells and included the prolactin signaling pathway, thyroid hormone signaling pathway, and type I diabetes mellitus (FDR < 0.05, Supplementary Figure S12), which have been reported to potentially play crucial roles in COVID-19^44-46^. Recent single-cell sequencing studies^47-49^ have demonstrated that naïve CD8+ T cells show prominent associations with COVID-19 severity. Moreover, naïve CD8+T cells are essential for recognizing newly-invaded viral antigens including SARS-CoV-2, leading to initiation of the adaptive immune response by differentiating naïve T cells into subpopulations of cytotoxic effector CD8+ T cells or memory CD8+ T cells^48,50,51^.

As shown in Figure 5E, the naïve CD8+T cells were grouped into four clusters. We found that trait-relevant cells with high scPagwas TRSs were mainly in clusters 0 and 1 (Figure 5F and Supplementary Figure S13). Of note, cluster 0 showed high expression of memory effector marker genes (*GZMK, AQP3, GZMA, PRF1*, and *GNLY*), while cluster 1 demonstrated high expression of exhaustive effector marker genes (*LAG3, TIGIT, GZMA, GZMB, PRDM1*, and *IFNG*) (Supplementary Figure S14). Further analysis showed that the molecular signature scores of the effector marker genes across cells were significantly positively correlated with the TRSs (Figure 5G and Supplementary Table S5), indicating that severe COVID-19-associated T cells tend to activate effector signatures involved in the anti-viral immune response. These new cell subpopulations may play important roles in modulating the immune response in severe COVID-19 patients.

### scPagwas distinguishes heterogeneous cell populations associated with AD

AD is a detrimental neurodegenerative disease that causes a gradual increase in neuronal death and loss of cognitive function. scPagwas was applied to uncover cell subpopulations associated with AD by integrating a human brain entorhinal cortex single-nucleus RNA-seq (snRNA-seq) dataset containing five brain cell types (n = 11,786 cells, Figure 6A and Supplementary Table S2) with an AD GWAS summary dataset (N = 21,982 cases and 41,944 controls, Supplementary Table S1). We found that OPCs and microglia with higher TRSs showed stonger enrichments in AD (Figure 6B). Consistently, at the cell type level, both OPCs and microglia were significantly associated with AD (FDR < 0.05, Figure 6C and Supplementary Table S6). For independent validation, three large single-cell datasets (Supplementary Table S1), including two human brain snRNA-seq datasets (n = 101,906 and 14,287 cells) and one mouse brain scRNA-seq dataset (n = 160,796 cells), were used for scPagwas analysis. These results also indicated that OPCs and microglia were significantly associated with AD (P < 0.05, Supplementary Table S7).

**Figure 6.**
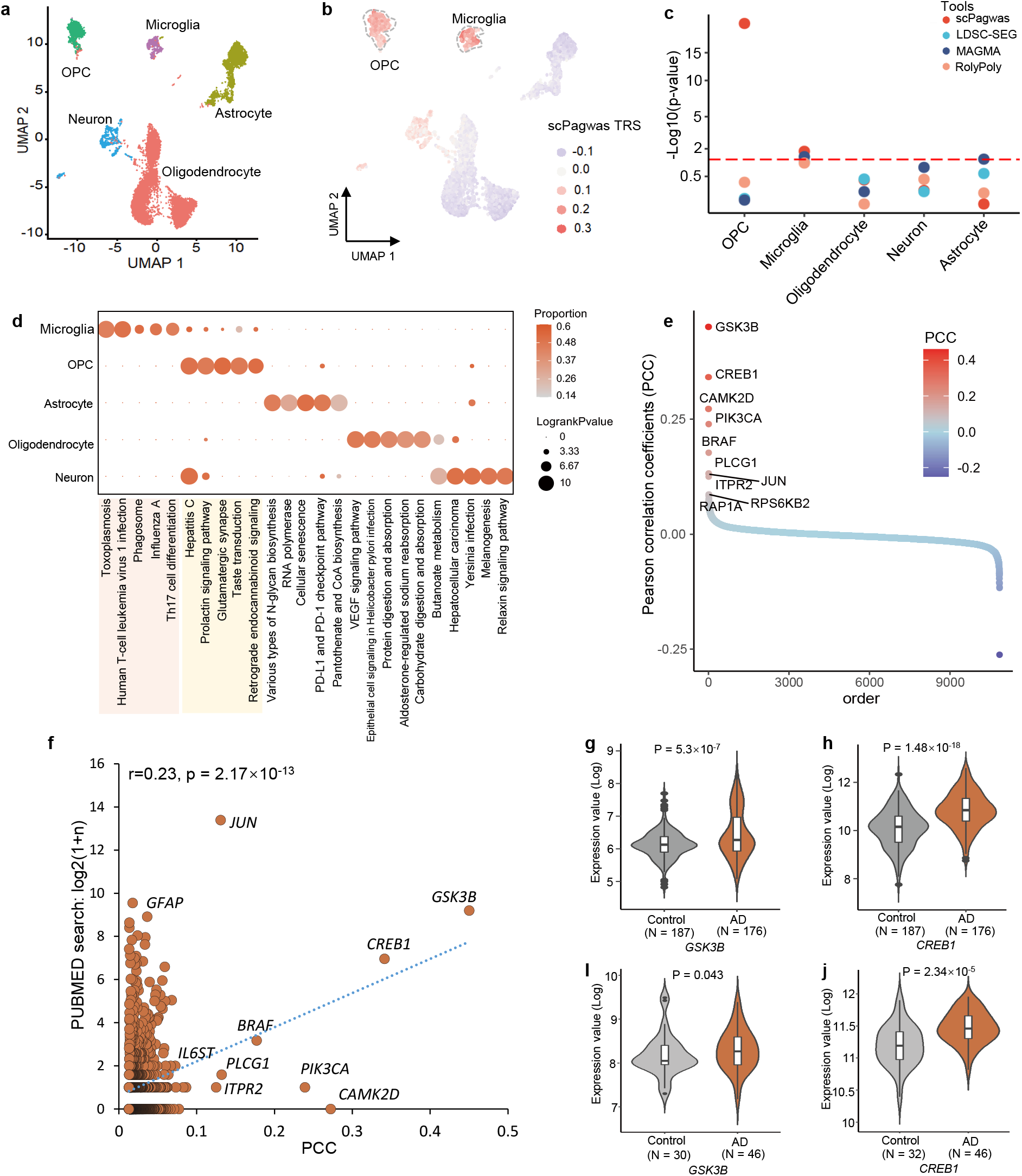
scPagwas discerns human brain cell types and subpopulations in association with AD. A. The UMAP plot of scRNA-seq profiles of 11,786 human brain cells containing five brain cell types. Cells are colored by the cell type annotation. B. scPagwas TRS for the phenotype of AD risk is displayed for all cells in the UMAP plot. OPC and microglial cells are highlighted with dashed lines. C. Benchmarking analysis of uncovering significant AD-associated cell types by using scPagwas, LDSC-SEG, MAGMA-based approach, and RolyPoly. The horizontal dashed red line represents the significant threshold (P < 0.05). D. Dot plot demonstrating the trait-relevant pathways across five brain cell types identified by scPagwas. Dot size represents the log-ranked P value for each pathway, and color intensity indicates the proportion of cells within each cell type genetically influenced by a given pathway (pathway-level coefficient beta > 0). E. Trait-relevant genes ranked by the Pearson correlation coefficients (PCCs) using scPagwas across all individual cells. F. Correlations of trait-relevant genes for AD ranked from scPagwas results and PubMed search results. The Pearson correlation is calculated between scPagwas results and PubMed search results (log2(n+1)). The top-ranking trait-relevant genes are labelled. G-H. Violin plots show the differential gene expression (DGE) analyses of *GSK3B* and *CREB1* between AD patients and controls in two independent bulk-based expression profiles. The two bulk transcriptomic datasets (GSE109887 with 78 samples, and GSE15222 with 363 samples) contain 222 AD patients and 219 controls. The two-sided Student’s T test was used for assessing the statistical significance. See also Supplementary Table S9.

Remarkably heterogeneous associations between OPCs and AD were detected by scPagwas (heterogeneous FDR = 3.33×10^−4^, Supplementary Figure S15), which is consistent with the recent finding of functionally diverse states of OPCs^52^. Disruption of OPCs is related to accelerated myelin loss and cognitive decline and is considered an early pathological sign of AD^53^. Analogous to our results, a recent genetic study demonstrated that OPCs exhibit significant associations with schizophrenia^54^, which was repeated by scPagwas using the same schizophrenia GWAS and scRNA-seq data as in the Agarwal et al. study (Supplementary Figure S16). Consistently, multiple lines of genetic evidence have indicated a critical role of microglia in the pathogenesis of AD^55-58^.

Moreover, the top significant trait-relevant pathways of the microglial association were related to immune pathways, including Th17 cell differentiation and influenza A (Figure 6D). Genes in the immune pathway of Th17 cell differentiation in disease-associated microglia have been identified as involved in AD risk^59^. The top-ranked significant pathways for OPC associations were related to brain development and synaptic transmitters, including glutamatergic synapses, taste transduction, and the prolactin signaling pathway (Figure 6D). Alteration of glutamatergic synapses has the potential to inhibit OPC proliferation and may be related to disruption of myelination, which is a prominent feature of AD^60^. These results suggest that these trait-relevant cell types could contribute to AD risk via distinct biological pathways.

We further identified the 1,000 top-ranked trait-relevant genes for AD by computing the correlation between the expression of a given gene and the summed gPASs of each cell across all 11,786 brain cells (see Methods and Figure 6E). To assess the association between these prioritized genes and AD, we adopted the RISmed method^61^, which searches for supporting evidence from reported studies in the PubMed database. A significant positive correlation was observed between the scPagwas results and PubMed search results (r = 0.23, P = 2.17×10^−13^, Figure 6F), which was notably higher than that from matched random gene sets (permuted P < 0.01, Supplementary Figure S17). These risk genes were significantly enriched in several important functional cellular components related to neurodegenerative diseases, including postsynaptic specialization and neuron-to-neuron synapses (FDR < 0.05, Supplementary Figure S18 and Table S8). To further evaluate the phenotypic associations of these top 1,000 scPagwas-identified risk genes, we leveraged two large and independent bulk-based expression profiles on AD patients (N = 222) and matched controls (N = 219). We found that 42.1% (421/1,000) of these genes were significantly up-regulated in AD patients (two-sided T test P < 0.05, Supplementary Table S9). Of note, this proportion was significantly higher than that of randomly selected length-matched genes (permuted P = 0.01, Supplementary Figure S19).

The highest-ranked genes, *CREB1* (P = 1.48×10^−18^ and 2.34×10^−5^) and *GSK3B* (P = 5.3×10^−7^ and 0.043), exhibited significantly higher expression in AD patients than in controls in both datasets (Figure 6G– J). Genetic variants in *CREB1* have been associated with brain-related phenotypes, including neuroticism^62^, major depressive disorder^63^, and cognitive performance^64^. Inhibition of *GSK3B* expression decreases microglial migration, inflammation, and inflammation-associated neurotoxicity^65^. In addition, activation of the kinase *GSK3B* promotes TAU phosphorylation, which corresponds to amyloid-β (Aβ) accumulation and Aβ-mediated neuronal death^66^. In summary, scPagwas not only identified subpopulations of microglia and OPC relevant to AD but also uncovered the key AD-associated pathways and risk genes.

## Discussion

Here, we introduce scPagwas, a pathway-based polygenic regression method that incorporates GWAS summary statistics and scRNA-seq data to identify trait-relevant individual cells. scPagwas exhibits well-calibrated and powerful performance benchmarked with extensive simulated and real datasets. scPagwas can capture the essential trait-relevant features of single-cell data and provide previously unrecognized functional insights by linking trait-relevant genetic signals to the cellular context. It should be noted that scPagwas does not require parameter tuning for cell type annotations and significantly enhances the discovery of trait-relevant enrichment at the single-cell resolution compared to existing methods^11-15^. scPagwas is suitable for analyzing genetic enrichment of rare or previously unknown cell populations in large-scale single-cell datasets^67^.

High sparsity and technical noise are the principal issues in analyzing single-cell sequencing data^22,24^. The activity of individual genes cannot represent cell functionality because it highly relies on the activity of other partner genes in a given pathway^23^. Additionally, the combination of biological functions of different genes in the same pathway has been reported to reduce inflated zero counts and technical noise^21,24-26,28^. Furthermore, disease-associated genetic variants that are mainly involved in systems of highly communicating genes and even variants with weak associations grouped in a given biological pathway could play important roles in uncovering the genetic mechanisms of complex diseases or traits^30,31^. Leveraging these pathway-based advantages, scPagwas could reduce the high sparsity and technical noise across millions of scRNA-seq profiles from different tissues and organs from mice and humans. Crucially, scPagwas not only recapitulated well-established cell type-disease associations, including the associations of immune cell types with hematological traits and microglia with AD, but also detected notable enrichments that have not been reported in previous studies and are biologically plausible, supporting the powerful potential of scPagwas for the discovery of novel mechanisms.

Based on the correlation between genetically-influenced pathway activity and gene expression, scPagwas prioritized more trait-relevant genes than other widely used gene-scoring methods, including MAGMA^20^, S-PrediXcan^19^, S-MultiXcan^18^, and TWAS^17^ (Figure 2). Although the use of scDRS with the MAGMA-identified genes was more powerful than with the genes identified by the other three methods (i.e., S-PrediXcan, S-MultiXcan, and TWAS), scPagwas yielded the best performance in distinguishing trait-relevant cells. When the top 1,000 scPagwas-identified trait-relevant genes were applied to scDRS, the precision for distinguishing trait-relevant cells was significantly enhanced, indicating that it is important to prioritize a group of robust trait-relevant genes for scoring cells. This explains why previous cell-scoring methods^16,27,29^ based on the top-ranked MAGMA genes only achieved moderate performance. Moreover, scPagwas could discern trait-relevant cell populations as well as early progenitor cells for blood cell traits, which is consistent with the fact that pathway-based scoring methods are useful in determining disease-associated but heterogeneous early developmental progenitor cells^28,68,69^.

Several limitations of this study should be noted. First, identification of a statistical association between complex diseases/traits and individual cells does not imply causality but may reflect indirect identification of causal associations, parallel to previous methods^11,13,14,16^. Nevertheless, even under such circumstances, the scPagwas-identified trait-relevant cells are inclined to be biologically relevant to the causal cells because of their similar genetic co-expression patterns. Second, for the current study, we selected canonical pathways identified in the Kyoto Encyclopedia of Genes and Genomes (KEGG) database^70^ because these pathways have been experimentally validated. Third, to be compatible with Seurat software^37^, the most extensively used tool for scRNA-seq data analysis, scPagwas by default employed the cell-scoring method of the *AddModuleScore* function of Seurat to directly compute the TRS, which makes it convenient to integrate scPagwas into existing scRNA-seq analysis pipelines. According to our current results, other state-of-the-art cell-scoring methods, including the scDRS^16^, AUCell^27^, and Vision^29^, also showed a good and robust performance for distinguishing trait-relevant cells when applying scPagwas-identified trait-relevant genes. Finally, we annotated SNPs into genes and their corresponding pathways based on the proximal distance of a 20 kb window. It may be possible to establish the link between SNPs and genes using other methods, such as functionally-informed SNP-to-gene linking approaches^71^, in the future.

In conclusion, scPagwas demonstrates promise for uncovering significant trait-relevant individual cells. Our pathway-based inference strategy will increase the identification of key cell subpopulations with reasonable biological interpretation for traits of interest. From a discovery viewpoint, the identification of reproducible trait-relevant individual cells will help to achieve the first step toward an in-depth experimental investigation of novel cell types or states with potential physiological roles in health and disease.

## Data Availability

All data produced in the present study are available upon reasonable request to the authors

https://gwas.mrcieu.ac.uk/

https://www.ebi.ac.uk/gwas/

https://jeffgranja.s3.amazonaws.com/MPAL-10x/Supplementary_Data/Healthy-Data/scRNA-Healthy-Hematopoiesis-191120.rds

https://www.ebi.ac.uk/arrayexpress/experiments/E-MTAB-10026/

https://storage.googleapis.com/linnarsson-lab-loom/l5_all.loom

https://www.ncbi.nlm.nih.gov/geo/query/acc.cgi?acc=GSE138852

https://www.ncbi.nlm.nih.gov/geo/query/acc.cgi?acc=GSE160936

https://www.ncbi.nlm.nih.gov/geo/query/acc.cgi?acc=GSE140231

## Resource Availability

### Lead contact

Further information and requests for resources and reagents should be directed to and will be fulfilled by the Lead Contact, Jianzhong Su.

### Materials availability

This study did not generate new unique reagents

### Data and Code Availability Statements

All GWAS summary datasets were downloaded from three publicly accessible databases of the IEU open GWAS project (https://gwas.mrcieu.ac.uk/), the COVID-19 Host Genetics Initiative (www.covid19hg.org/results), the Psychiatric Genomics Consortium website (https://pgc.unc.edu/), and the NHGRI-EBI GWAS Catalog (https://www.ebi.ac.uk/gwas/). The healthy BMMC scRNA-seq dataset was downloaded from a website (https://jeffgranja.s3.amazonaws.com/MPAL-10x/Supplementary_Data/Healthy-Data/scRNA-Healthy-Hematopoiesis-191120.rds). The healthy PBMC scRNA-seq dataset used to validate scPagwas performance was downloaded from the ArraryExpress database (https://www.ebi.ac.uk/arrayexpress/experiments/E-MTAB-10026/). The mouse brain scRNA-seq dataset was downloaded from the Mouse Brain Atlas (https://storage.googleapis.com/linnarsson-lab-loom/l5_all.loom). Human brain snRNA-seq dataset #1 (https://www.ncbi.nlm.nih.gov/geo/query/acc.cgi?acc=GSE138852), Human brain snRNA-seq dataset #2 (https://www.ncbi.nlm.nih.gov/geo/query/acc.cgi?acc=GSE160936), Human brain snRNA-seq dataset #3 (https://www.ncbi.nlm.nih.gov/geo/query/acc.cgi?acc=GSE140231), and two bulk-based transcriptomic profiles on AD (1. https://www.ncbi.nlm.nih.gov/geo/query/acc.cgi?acc=GSE109887; 2. https://www.ncbi.nlm.nih.gov/geo/query/acc.cgi?acc=GSE15222) were downloaded from the GEO database. The PBMC scRNA-seq dataset on COVID-19 severity was downloaded from the ArraryExpress database (https://www.ebi.ac.uk/arrayexpress/experiments/E-MTAB-9357). The scRNA-seq dataset on the human cell landscape (HCL) was downloaded from the GEO database (https://www.ncbi.nlm.nih.gov/geo/query/acc.cgi?acc=GSE134355). scPagwas is implemented as an R package and is available on GitHub (https://github.com/dengchunyu/scPagwas). The code to reproduce the results is available in a dedicated GitHub repository (https://github.com/dengchunyu/scPagwas_reproduce).

## Method details

### scPagwas methodology

The workflow of the scPagwas method is shown in Figure 1. In brief, scPagwas employs an optimized polygenic regression model to identify the associations of a subset of cells with a complex disease or trait of interest. The framework of the method is described in detail in the following steps.

### Linking SNPs to their corresponding pathways

Based on previous evidence^6^ indicating that most eQTLs consistently lie in a 20-kb window centered on the transcription start site of a gene, a window size of 20 kb is adopted as the default parameter of scPagwas to assign SNPs from GWAS summary statistics to associated genes. We use the notation *g*(*k*) to represent a gene *g* with an SNP *k*. With the assignment of SNPs to corresponding gene, there are a few SNPs with multiple associated genes. We duplicate these SNPs and consider them as independent SNP -gene pairs following an earlier study^13^. In our data applications, SNPs with minor allele frequencies smaller than 0.01 or on the sex chromosomes (ChrX-Y) were removed.

Based on pathways in the KEGG database^70^, we annotate these SNPs to associated gene *g* in corresponding pathway, and use the notation *S*_*i*_ = {*k* : *g*(*k*) ∈ *P*_*i*_ } to indicate the set of SNPs within the pathway *i*. The notation *P*_*i*_ indicates the set of genes in the pathway *i*. scPagwas provides other functional gene sets, such as Reactome ^72^ and MSigDB^73^, as alternative options. In our data applications, the 1,000 Genomes Project Phase 3 Panel ^74^ was applied to calculate the linkage disequilibrium (LD) among SNPs available in the GWAS summary statistics, and the major histocompatibility complex region (Chr6: 25–35 Mbp)^75^ was removed because of the extensive LD in this region.

### PAS matrix transformation

scPagwas uses the variance-stabilizing transformation method^76^ with a scale factor of 10,000 to normalize a sparse gene-by-cell matrix from scRNA-seq data as follows: 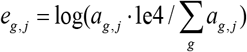, where *a*_*g, j*_ is the raw expression for gene *g* in cell *j* and *e*_*g, j*_ is the normalized expression of gene *g* in cell *j*. Pathways such as those from the KEGG database^70^ can be used as a gene set to calculate PASs. The SVD method can greatly improve the computational efficiency^22,77^ of analyzing a sparse matrix with high dimensionality and can be used to generate eigenvalues without calculating the covariance matrix. We apply the SVD method to transform a normalized gene-by-cell matrix into a pathway-by-cell matrix with reduced dimensional space.

For each pathway *i*, we extract a *N* × *M*_*i*_ sub-matrix ***A***_*i*_ from the normalized single-cell matrix ***A***, with *N* being the number of cells and *M*_*i*_ being the number of genes in pathway *i*. Applying the SVD method, ***A***_*i*_ can be decomposed as follows:

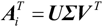

where ***U*** is an *N* × *N* orthogonal matrix, ***Σ*** is a diagonal matrix with all zeroes except for the elements on the main diagonal, and ***V*** ^*T*^ is an *M*_*i*_ × *M*_*i*_ orthogonal matrix. For the right orthogonal matrix 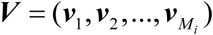, the *t*th column vector ***v***_*t*_ represents the *t*th principal component. In reference to previous studies ^28,35^, we use the projection of the characteristics of genes in each pathway on the direction of the PC1 eigenvalue to define PAS *s*_*i, j*_ for the pathway *i* in cell *j*, which reflects the main coordinated expression variability of genes in a given pathway among single-cell data.

### Polygenic regression model

According to a previous method^13,34^, we assume a linear regression model, ***y*** = ***Xb*** + ***ε***, where ***y*** is an *n* − vector of phenotypes, ***X*** denotes the *n*× *m* matrix of genotypes (standardized to mean 0 and variance 1 for each SNP vector), ***b*** indicates the per-normalized-genotype effect sizes vector of *m* SNPs when fitted jointly, and ***ε*** is the stochastic environmental error term. The released GWAS summary dataset contains per - SNP effect estimates, denoted as 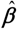. These estimates indicate the marginal regression coefficients from univerate models and can be calculated using the transformation equation 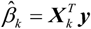, where 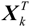 represents standardized genotypes for SNP *k* across *n* GWAS samples. After substituting the polygenic model ***y*** = ***Xb*** + ***ε*** into the estimation equation 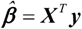, the estimated marginal effect sizes of SNPs can be written as:

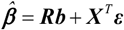

where ***R*** denotes the LD matrix.

As previously mentioned, *S*_*i*_ denotes an SNP set that contains SNPs mapped to genes in a pathway *i*. The polygenic model assumes that the effect sizes of SNPs in pathway *i* are random effects, which follow the multi-variable normal distribution 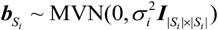, where 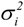 is the variance of effect sizes for SNPs in the pathway and ***I*** is the | *S*_*i*_ | × | *S*_*i*_ |identity matrix. Based on the prior assumption of above polygenic model, the distribution for the vector of the estimated effects of SNPs 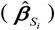 associated with a pathway follows:

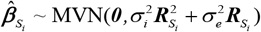

In reference to the extension of stratified LD score regression to continuous annotations ^34^, the per-normalized SNP estimates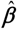 is a mean 0 vector whose variance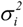 depends on continuous-valued annotations (in this case, expression levels of genes in a given pathway). Based on the assumption that a positive correlation between genetic associations and ge ne expression levels in each cell associated with a trait of interest, the variance 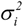 is modeled using the linear weighted sum method for each SNP *k*:

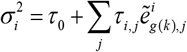

where *τ* _*0*_ is an intercept term, *τ*_*i, j*_ is the coefficient for the pathway *i* in cell *j*, which measures the strength of association between pathway-specific gene expression activity and the variance of GWAS effect sizes at the single-cell level, and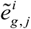is the adjusted gene expression for each gene *g* in the given pathway *i* calculated as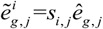 with *s*_*i, j*_ being the PAS of pathway *i*. For each gene *g* in the pathway *i*, the gene expression *e*_*g, j*_ is rescaled using the min-max rescaling method:

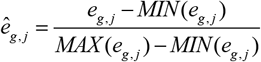

where *MAX* (*e*_*g, j*_) denotes the maximum gene expression in pathway *i* and *MIN* (*e*_*g, j*_) denotes the minimum gene expression in pathway *i*.

To optimize the coefficients for each pathway in cells under the polygenic regression model, scPagwas adopts the method-of-moments approach, which can prominently improve the computational efficiency and the estimated uniform convergence ^13^. Then, the observed and expected squared effects of SNPs relevant to each pathway are fitted, and the following equation is used to estimate the expected value:

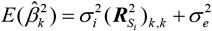

where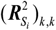 represents the *k*th diagonal element of matrix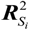 Then, the coefficient *τ*_*i, j*_ can be estimated using the following linear regression:

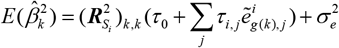

Of note, the estimated coefficient 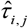 represents the per-SNP contribution of one unit of the pathway-specific activity to heritability. We define a gPAS for each pathway *i* that is calculated by the product between the estimated coefficient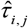 and weighted PAS using the following equation: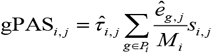, where *M*_*i*_ is the number of genes in the pathway *i*. Essentially, gPAS is a pathway-activity-based prediction of the genetic variance of a normal distribution of cis-GWAS effect sizes for pathway *i* (Figure 1D). Note that the larger gPASs would have larger *cis*-GWAS effects for each cell, and gPASs can be used to rank trait-relevant pathways (see Supplementary Methods).

### Identification of trait-relevant genes and individual cells

To optimize genes relevant to complex diseases/traits at single-cell resolution, we determine which gene *g* exhibits expression that is highly correlated with the summed gPASs across individual cells using the Pearson correlation method. To maximize the power, the expression of each gene *g* is inversely weighted by its gene-specific technical noise level, which is estimated by modeling the mean-variance relationship across genes in the scRNA-seq data^78^. By arranging the PCCs for all genes in descending order, we select the top-ranked risk genes as trait-relevant genes (default top 1,000 genes) according to a previous method^16^.

Subsequently, we quantify the aggregate expression of predefined trait-relevant genes in each cell to generate raw TRSs. For a given cell *j* and a trait-relevant gene set *B*, the cell-level raw TRS, TRS _*j*_, is defined as the average relative expression of the genes in *B*. However, such raw TRSs may be confounded by cell complexity, as cells with higher complexity would have more genes identified and consequently tend to have higher TRSs for any given gene set. To properly control for the effect of cell complexity, we calculate a control cell score with a control gene set *B*^*Ctrl*^, which is randomly selected in a manner that maintained a comparable distribution of expression levels to that of the predefined gene set. The process included two steps: 1) using the average expression levels to group all analyzed genes into 25 bins of equal size and 2) randomly selecting 100 genes from the same expression bin for each gene in the predefined gene set. The final TRS is defined as the initial raw TRS after subtracting its corresponding control cell score: 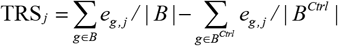. The *AddModuleScore* cell-scoring method in Seurat^37^ is employed to calculate the TRS with default parameters.

To further assess whether a cell is significantly associated with the trait of interest, we employ an approach^36^ to determine the statistical significance of individual cells by calculating the ranking distribution of trait-relevant genes. Initially, the percent ranks of these trait-relevant genes across the cells yielded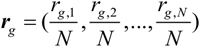, where *r*_*g, j*_ is the expression rank of gene *g* in cell *j* and *N* is the total number of cells. The percent ranks of genes follow a uniform distribution *U* (0,1). Under the null hypothesis that there is no relationship between the percent rank of genes, a statistic *T*_*j*_ for each cell is calculated using the formula

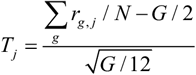

In view of a large number of cells is included in the single-cell data, the distribution of *T*_*j*_ can be deduced using the central limit theorem^36^: 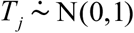, where *G* is the number of selected trait-relevant genes. The hypothesis for the significance test is *H*_0_ : *T*_*j*_ = 0 *vs H*_1_ : *T*_*j*_ > 0. The P value for each cell *j* can be written as *p* _*j*_ = Pr(*T*_*j*_ ≤ *t*).

### Inference analysis of trait-relevant cell types

scPagwas can also identify trait-relevant cell types, where the set of cells is treated as a pseudo-bulk transcriptomic profile and the expression of a gene across cells is averaged within a given cell type. For the cell type association, the block bootstrap metho d^79^ is used to estimate the standard error and compute a t -statistic with a corresponding P value for each cell type. Because the goal of the block bootstrap is to maintain data structures when sampling from the empirical distribution, we leverage all pathways in the KEGG database^70^ to partition the genome into multiple biologically-meaningful blocks and sample these pathway-based blocks with replacement. Under default parameters, scPagwas performs 200 block bootstrap iterations for each cell-type association analysis. The optional parameters are provided for the block bootstrap. Detailed information on scPagwas cell type –level inference analysis can be found in the Supplementary Methods.

### Simulations

We used scDesign2 (version 1.0.0) ^80^ to simulate a ground truth scRNA-seq dataset containing five cell types including monocytes, DCs, and B, NK, and T cells to assess the performance of scPagwas in identifying monocyte count trait-relevant individual cells. DC, a type of cell differentiated from monocytes^81^, was chosen as a non-trait-relevant cell type, which could be a confounding factor for distinguishing monocytes from all simulated cells. In the model-fitting step, we first fitted a multivariate generative model to a real dataset via the fluorescence-activated cell-sorted bulk hematopoietic populations downloaded from the GEO database (Accession No. GSE107011)^82^. Because there were five sorted cell types, we divided the datasets into five subsets according to the cell types and fitted a cell type-specific model to each subset. In the data-generation step, we generated a synthetic scRNA-seq dataset from the fitted model to represent trait-relevant cell populations (monocytes) and non-trait-relevant cell populations (non-monocyte cells including DCs and B, NK, and T cells) for the monocyte count trait. Finally, we obtained 2,000 cells with synthetic scRNA-seq data with cell proportions of 0.5 (monocytes), 0.05 (DCs), 0.2 (B cells), 0.05 (NK cells), and 0.2 (T cells).

### scRNA-seq datasets

Eight independent scRNA-seq or snRNA-seq datasets spanning 1.4 million human (*Homo sapiens*) and mouse (*Mus musculus*) cells were used in this study (Supplementary Table S1). For blood cell traits, we collected two scRNA-seq datasets based on human BMMCs (n = 35,582 cells)^39^ and human PBMCs (PBMC #1, n = 97,039 cells)^40^ to identify trait-relevant cell subpopulations or types. For AD, we collected four single-cell datasets including a mouse brain scRNA-seq dataset (n = 160,796 cells)^83^, a human brain entorhinal cortex snRNA-seq dataset (Human brain #1, n = 11,786 cells)^84^, a human brain snRNA-seq dataset (Human brain #2, n = 101,906 cells)^85^, and another human brain snRNA-seq dataset (Human brain #3, n = 14,287 cells)^54^. To identify severe COVID-19-related immune cell populations, we collected a large-scale PBMC scRNA-seq dataset (PBMC #2, n = 469,453 cells) containing 254 peripheral blood samples from patients with various COVID-19 severities (mild N = 109 samples, moderate N = 102 samples, and severe N = 50 samples) and 16 healthy controls^86^. The scRNA-seq dataset from the human cell landscape (HCL, n = 513,707 cells in 35 adult tissues)^87^, as well as the previously mentioned four scRNA-seq datasets (i.e., BMMC, PBMC #1, Human brain #1, and Mouse brain), were used to assess the performance of scPagwas in reducing the sparsity and technical noise.

### GWAS summary datasets for complex diseases and traits

We obtained GWAS summary statistics for 10 blood cell traits (average N = 307,772) and AD (21,982 cases and 41,944 controls) from the IEU OpenGWAS, for schizophrenia (67,390 cases and 94,015 controls) from the Psychiatric Genomics Consortium, and for severe COVID-19 (7,885 cases and 961,804 controls) from the COVID-19 Host Genetics Initiative (Supplementary Table S2). The 10 blood cell traits included monocyte count, lymphocyte count, lymphocyte percent, mean corpus volume, neutrophil count, white blood count, eosinophil count, basophil count, mean corpuscular hemoglobin, and hemoglobin concentration.

## Acknowledgement

We acknowledge funding support from the National Natural Science Foundation of China (32200535 to Y.M; 61871294 and 82172882 to J.S), the Scientific Research Foundation for Talents of Wenzhou Medical University (KYQD20201001 to Y.M.), the Science Foundation of Zhejiang Province (LR19C060001 to J.S), Leading Innovative and Entrepreneur Team Introduction Program of Zhejiang (2021R01013 to J.Y.), Research Program of Westlake Laboratory of Life Sciences and Biomedicine (202208013 to J.Y.), and Westlake Education Foundation (101566022001 to J.Y.). We thank Dr. Zhenhui Chen from Wenzhou Medical University for providing helpful suggestions and manuscript revisions. We also thank all of the authors who have deposited and shared GWAS summary data in public databases and the authors who publicly released the scRNA-seq and snRNA-seq datasets.

## Author contributions

J.S., J.Y., and Y.M. conceived and designed the study. Y.M., C.D. and Y.Z. developed the method. Y.M., C.D., Y.Z, Y.R.Z., F.Q., D.J., G.Z., J.Q., and J.L. managed data collection. Y.M., C.D., Y.Z, Y.R.Z., and J.L. conducted the bioinformatics analysis and data interpretation. Y.M., J.S., C.D., Y.Z, J.Y., and Y.Z. wrote the manuscript. All authors reviewed and approved the final manuscript.

