## Supplemental Figures for "Polygenic regression uncovers trait-relevant cellular contexts through pathway activation transformation of single-cell RNA sequencing data"

***Supplementary Figures***

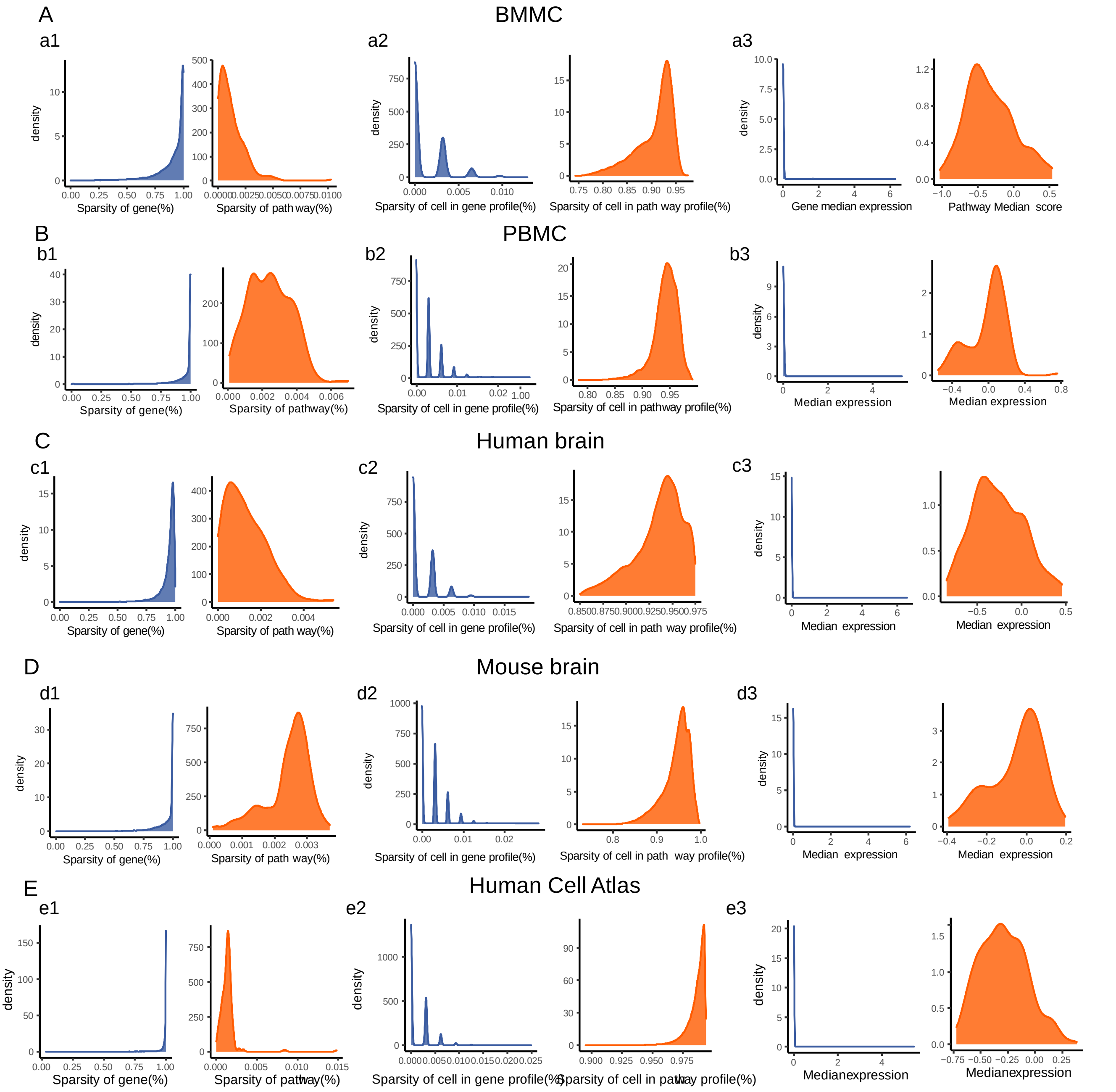


**Supplementary Figure S1. scPgwas remarkably reduce the sparsity among five distinct scRNA-seq datasets under three different conditions.** There were five scRNA-seq datasets derived from BMBC (n = 35,582 cells), PBMC (n = 97,039 cells), human brain (n = 11,786 cells), mouse brain (n = 160,796 cells), and human cell atlas (n = 513,707 cells) across human and mouse (A-E). Orange color represents plots related to PASs, and blue color represents plots related to the expression magnitude of individual genes. Condition #1: The sparsity of gene expression or PASs is defined as the proportion of cells that show no signal (zero-valued) for a given gene or pathway (a1, b1, c1, d1, and e1). Condition #2: The sparsity of cells is defined as the proportion of genes (expression magnitude) or pathways (PASs) that show no signal for a given cell (a2, b2, c2, d2, and e2). Condition #3: The distribution of the median gene expressions or median PASs across cells (a3, b3, c3, d3, and e3). The detailed information on these five scRNA-seq datasets are shown in the Supplementary Table S2.

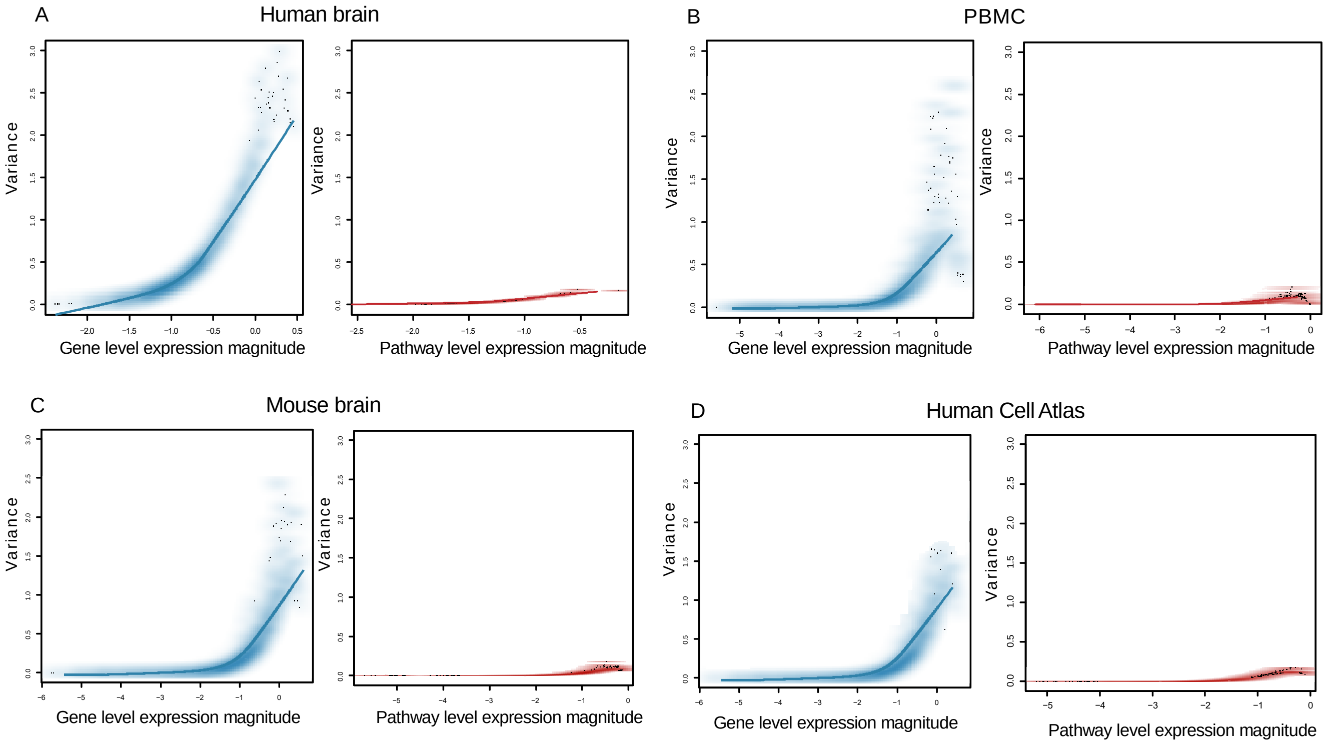


**Supplementary Figure S2. Plots demonstrating the variance of gene level expression magnitude and pathway level expression magnitude in four scRNA-seq datasets, related to Figure 2C.** There were four scRNA-seq datasets derived from human brain (n = 11,786 cells), PBMC (n = 97,039 cells), mouse brain (n = 160,796 cells), and human cell atlas (n = 513,707 cells) across human and mouse (A-D). Red color represents plots related to pathway level expression magnitude, and blue color represents plots related to the expression magnitude of individual genes. The left panel in each plot (A-D) shows a mean-variance fit, which demonstrates the relationship between average gene expression (x axis) and gene variance (y axis). The right panel in each plot (A-D) shows a mean-variance fit, which demonstrates the relationship between average expression of genes in a given pathway (x axis) and its corresponding variance (y axis). The detailed information on these four scRNA-seq datasets are shown in the Supplementary Table S2.

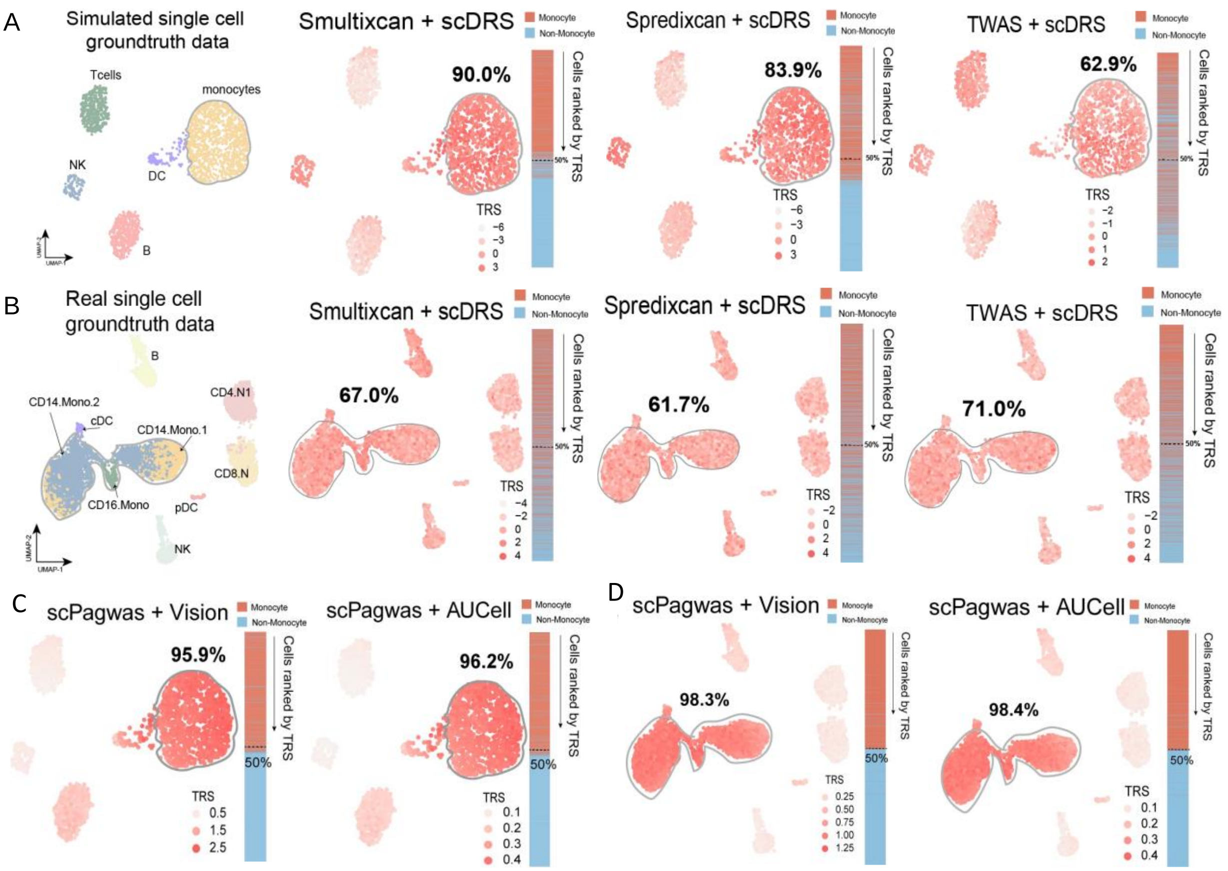


**Supplementary Figure S3. Assessment of the performance of different gene-scoring methods on scDRS, Vision, and AUCell for identifying trait-relevant cells, related to Figure 3.** A. UMAP embedding plot showing the performance of S-MultiXcan, S-PrediXcan, and TWAS on scDRS for identifying monocyte count trait-relevant cells in the simulated ground truth dataset (monocytes: n = 1,000 cells, and T, B, DC, and NK: n = 1,000 cells in total). B. UMAP embedding plot showing the performance of S-MultiXcan, S-PrediXcan, and TWAS on scDRS for identifying monocyte count trait-relevant cells in the real ground truth scRNA-seq dataset. The real BMMC scRNA-seq dataset contains 10,000 cells with seven cell types: monocytes (11_CD14.Mono.1, 12_CD14.Mono.2, and 13_CD16.Mono, n = 5,000 cells), DC (09_pDC and 10_cDC, n = 200 cells), T cells (19_CD8.N and 20_CD4.N1, n = 3,000 cells), B cells (17_B, n = 1,000 cells), and NK cells (25_NK, n = 800 cells). C-D). UMAP plot showing the performance of scPagwas-identified genes on Vision and AUCell for identifying monocyte count trait-relevant cells in the simulated (C) and real (D) ground truth scRNA-seq datasets. The UMAP projections of every cell colored by its TRS. The vertical bar exhibits cells descendingly ranked according to their corresponding TRSs (top-ranked 1,000 genes), where red color indicates monocyte cells and blue color indicates non-monocyte cells. The accuracy of each method represents the percentage of monocyte count trait-related cells (i.e., monocytes) for top-half cells that are ranked by TRS for all cells in a descending manner.

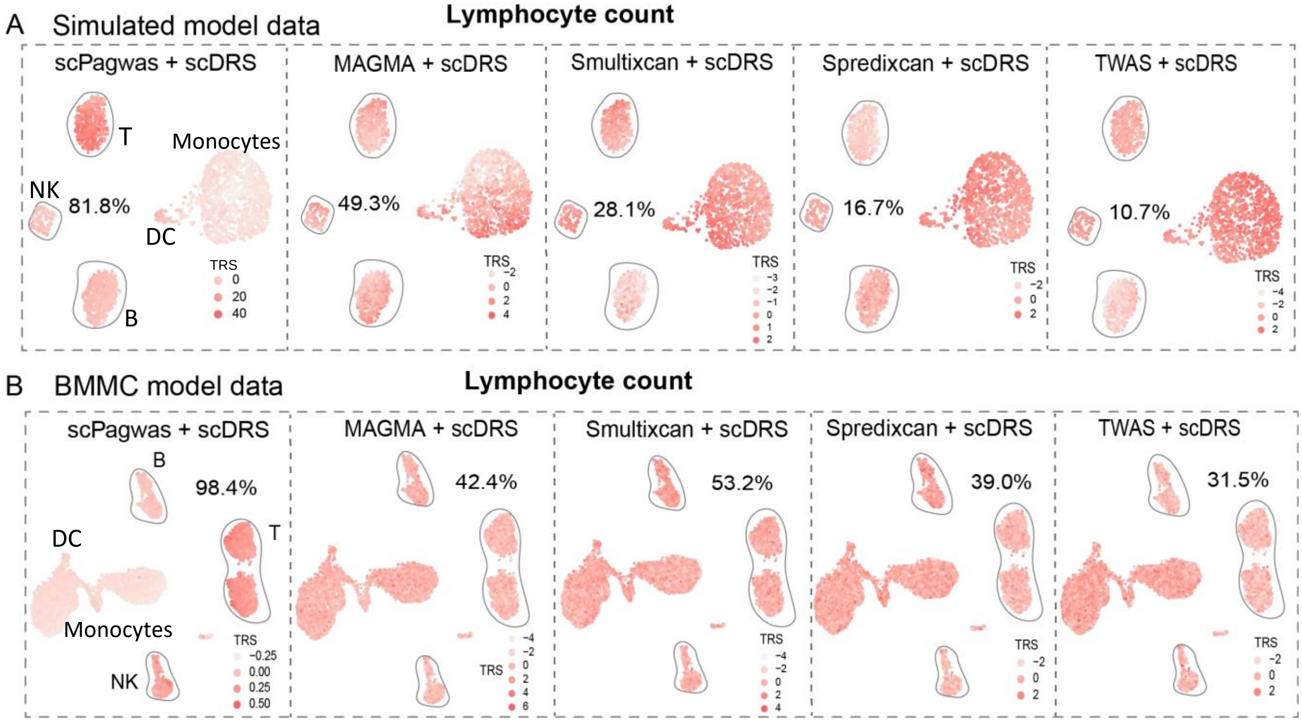


**Supplementary Figure S4. Benchmarking the performance of scPagwas with other four gene-based methods in both simulated and real ground truth scRNA-seq datasets.** A. Illustration of the performance of scPagwas and other four gene-based methods (i.e., MAGMA, S-MultiXcan, S-PrediXcan, and TWAS/FUSION) for lymphocyte count trait based on the scDRS cell-scoring method in a synthesized ground truth scRNA-seq dataset. This synthesized scRNA-seq dataset contained two main cell groups of monocyte cells (n = 1,000 cells) and non-monocyte cells (T, NK, B and DC, n = 1,000 cells). B. Illustration of the performance of scPagwas and other four gene-based methods (i.e., MAGMA, S-MultiXcan, S-PrediXcan, and TWAS/FUSION) for lymphocyte count trait based on the scDRS cell-scoring method in a real (BMMC) ground truth scRNA-seq dataset. The real dataset contains 10,000 cells with seven cell types: monocytes (11_CD14.Mono.1, 12_CD14.Mono.2, and 13_CD16.Mono, n = 5,000 cells), DC (09_pDC and 10_cDC, n = 200 cells), T cells (19_CD8.N and 20_CD4.N1, n = 3,000 cells), B cells (17_B, n = 1,000 cells), and NK cells (25_NK, n = 800 cells). The accuracy of each method represents the percentage of lymphocyte count trait-related cells (i.e., T, NK, and B) for top-half cells that are ranked by TRS for all cells in a descending order.

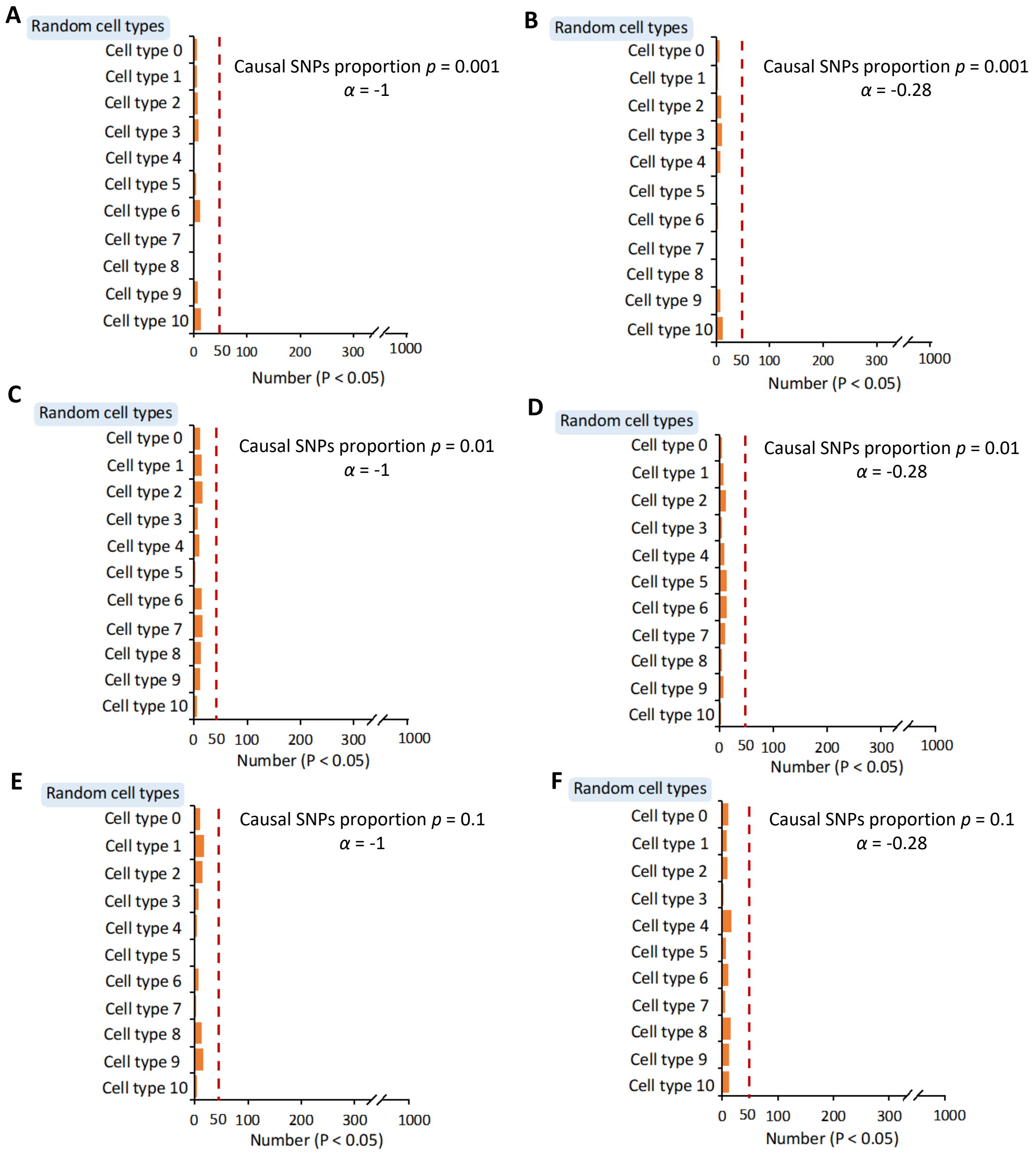


**Supplementary Figure S5. The performance of scPagwas approach on inferring cell-type relevance under six different genetic architectures.** Note: Number of associations with P < 0.05 after permuting 1,000 times of scPagwas analysis using randomly-selected scRNA-seq datasets (n = 5,000 cells) with six simulated GWAS summary statistics (see Supplementary Methods) according to the proportion of causal SNPs across the genome ($p$ = 0.001, 0.01 and 0.1), as well as the dependence of the SNPs' effects ($\alpha$ = -1 or -0.28). $\alpha$ = -1 represents MAF-independent where all SNPs equally contribute to variance, $\alpha$ = -0.28 represents MAF-dependent where the contributions of SNPs to variance were influenced by linkage disequilibrium. $p$ = 0.001, 0.01, and 0.1 represent sparse, medium, and dense polygenic genetic architecture, respectively (A-F). The red dash line stands for the number of results expected by chance at P < 0.05 for 1,000 permutations (n = 50). We found three times fewer significant results than expected by chance using the scPagwas model, indicating that scPagwas approach is conservative and credible.

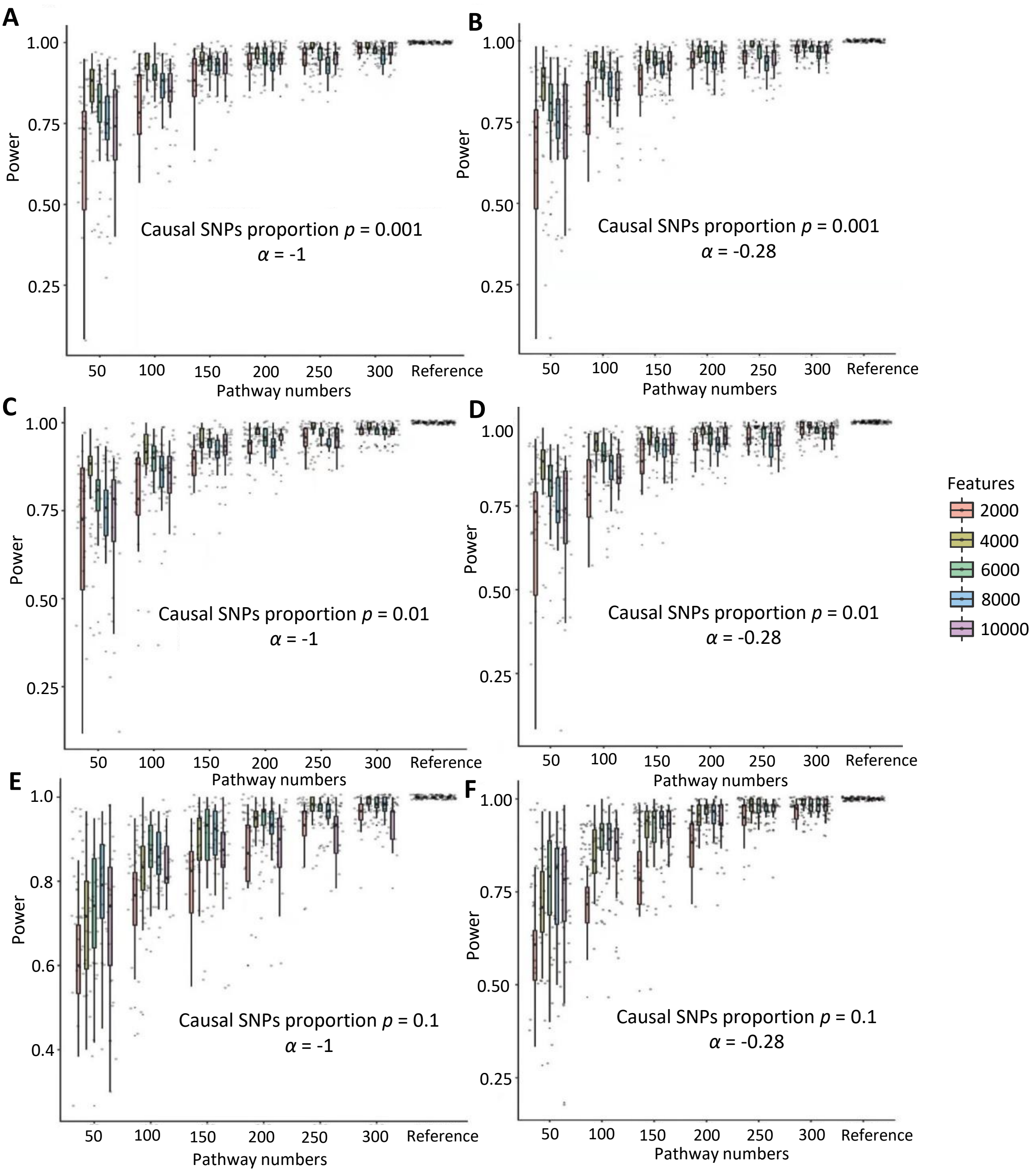


**Supplementary Figure S6. The power of various pathway numbers in scPagwas model.** Note: Based on five different features of selecting top-ranked genes with high expression variance (i.e., 2000, 4000, 6000, 8000, and 10000), we conducted scPagwas analyses to integrate a random scRNA-seq dataset with six simulated GWAS statistics (see Supplementary Methods) under sixs different pathway numbers (i.e., 50, 100, 150, 200, 250, and 300), which were randomly selected from the KEGG pathway database. All KEGG pathways were used as a reference. For each feature, we ran 20 times of scPagwas analyses based on different random selections. The results of scPagwas with different pathway numbers were correlated with that of the reference to calculate the correlation coefficients. We defined the correlation coefficient as “power” to reflect the effects of different pathway numbers on scPagwas performance at different conditions. We observed that scPagwas approach yielded a relatively stable and powerful performance under different pathway numbers.

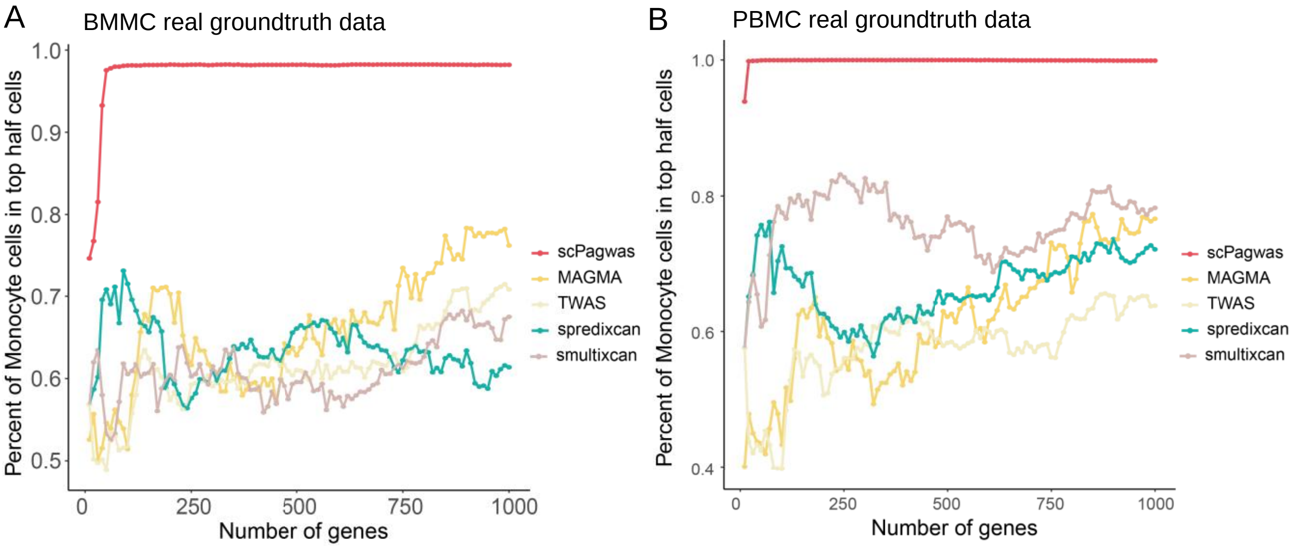


**Supplementary Figure S7. Assessing the performance of the number of included top trait-relevant genes from scPagwas and four other methods in scoring monocyte count trait-relevant cells, related to Figure 3.** A-B) Examination of the influence of included trait-relevant genes' number from scPagwas and four other methods (i.e., MAGMA, TWAS, S-PrediXcan, and S-MultiXcan) on the performance of scDRS cell-scoring method in two real (BMMC and PBMC) ground truth scRNA-seq datasets. A. The real BMMC dataset contains 10,000 cells with seven cell types: monocytes (11_CD14.Mono.1, 12_CD14.Mono.2, and 13_CD16.Mono, n = 5,000 cells), DC (09_pDC and 10_cDC, n = 200 cells), T cells (19_CD8.N and 20_CD4.N1, n = 3,000 cells), B cells (17_B, n = 1,000 cells), and NK cells (25_NK, n = 800 cells). B. The real PBMC dataset contains 10,000 cells with two main cell types: monocytes (CD14+monocytes, N = 5,000 cells) and non-monocytes (CD4+T cells and NK, N = 5,000 cells). For these assessments, the number of included trait-relevant genes started with top 10 genes and increased to top 1,000 genes by adding 10 genes at a time. For each analysis, the accuracy represents the percentage of monocyte count trait-related cells (i.e., monocytes) for top-half cells that are ranked according to the TRSs for all cells in a descending manner. x axis indicates the number of included genes, and y axis indicates the percent of monocyte cells in top half cells ranked by TRSs.

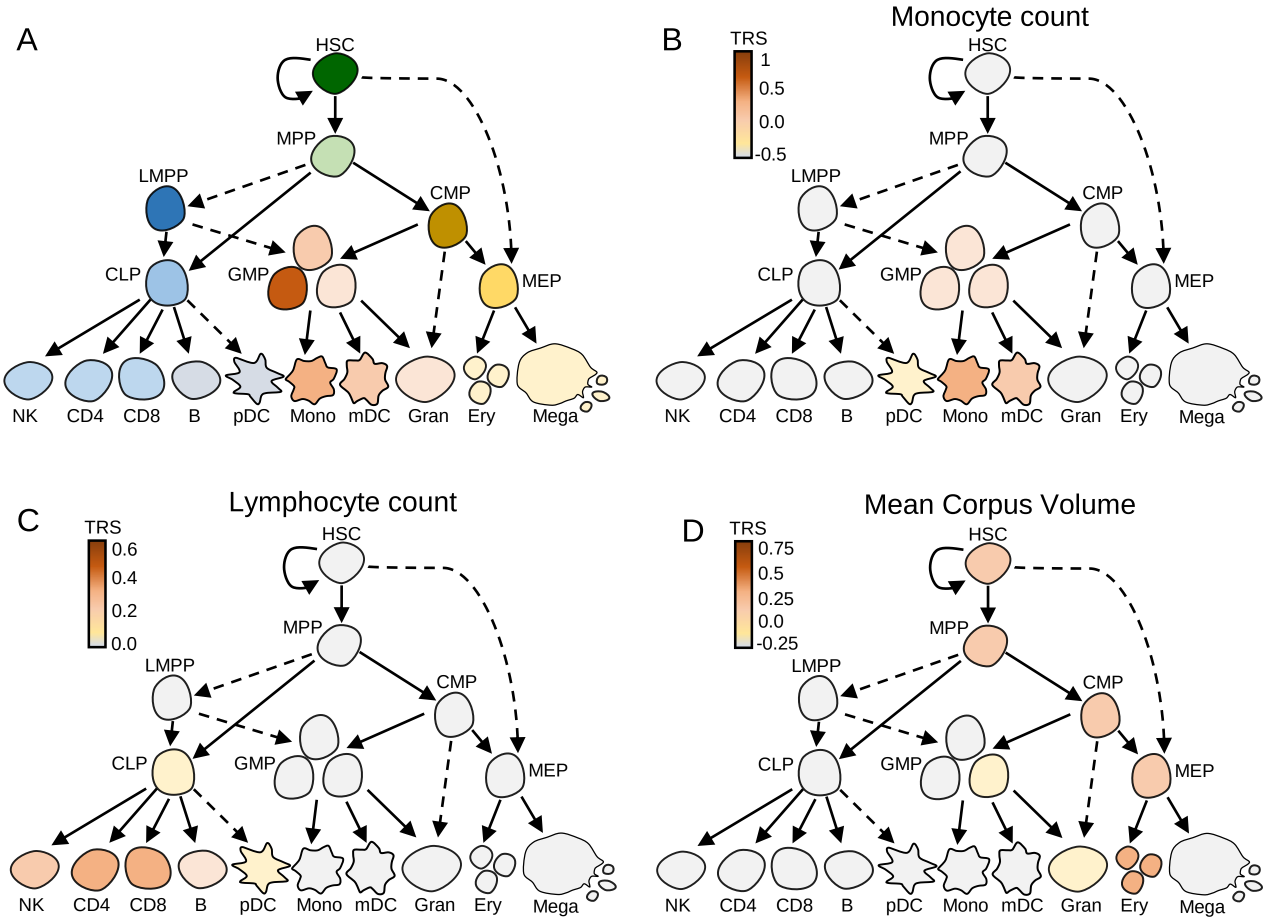


**Supplementary Figure S8. scPagwas TRSs of three representative traits in hematopoietic lineages at different stages of differentiation, related to Figure 3.** A) Schematic plot showing the hematopoietic lineages at different stages of differentiation. B-D) The TRSs of three representative blood cell traits (B: monocyte count, C: lymphocyte count, D: mean corpus volume) in relevant cell types across the hematopoietic lineages. These schematic plots showed the results in Figure 3C-E. HSC = hematopoietic stem cell; MPP = multipotent progenitor; LMPP = lymphomyeloid-restricted progenitors; CMP = common myeloid progenitor; CLP = common lymphoid progenitor; MEP = megakaryocyte and erythroblast progenitor; GMP = granulocyte macrophage progenitor; NK = natural killer; CD4 = CD4+T cell; CD8 = CD8+T cell; B = B-lymphocyte cell; pDC = plasmacytoid densdritic cell; Mono = monocyte; mDC = myeloid dendritic cell; Gran = granulocyte; Ery = erythrocyte; Mega = megakaryocyte.

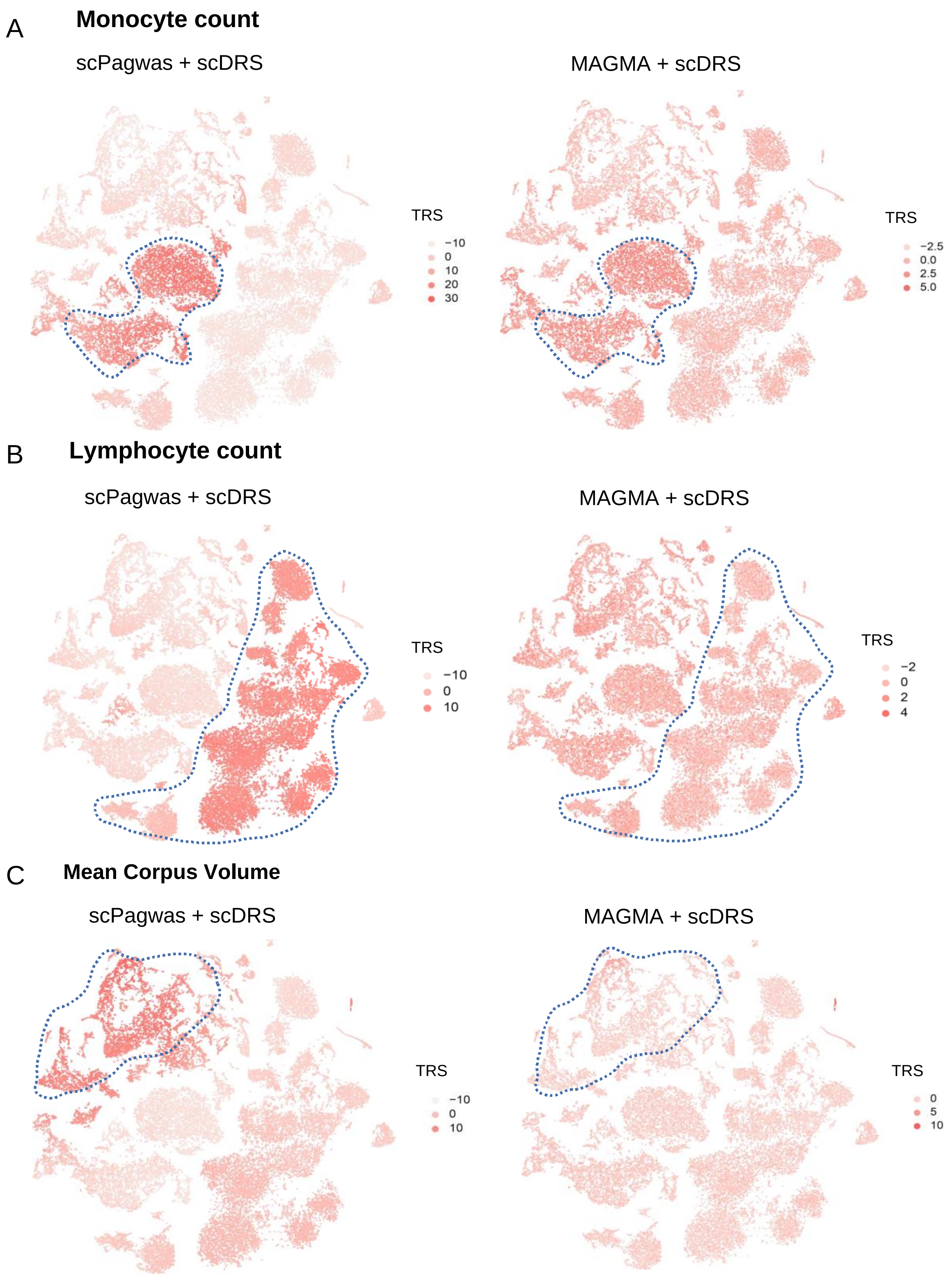


**Supplementary Figure S9. Illustration of per-cell scDRS TRS for three representative blood cell traits in the BMMC scRNA-seq dataset, related to Figure 4.** The tSNE plots showing the results of scDRS using top scPagwas 1,000 genes (left panel) and using top MAGMA 1,000 genes (right panel) for monocyte count (A), lymphocyte count (B), and mean corpus volume (C). Note: The analysis was performed based on the BMMC scRNA-seq dataset (n = 35,582 cells, see Figure 4 and Supplementary Table S2).

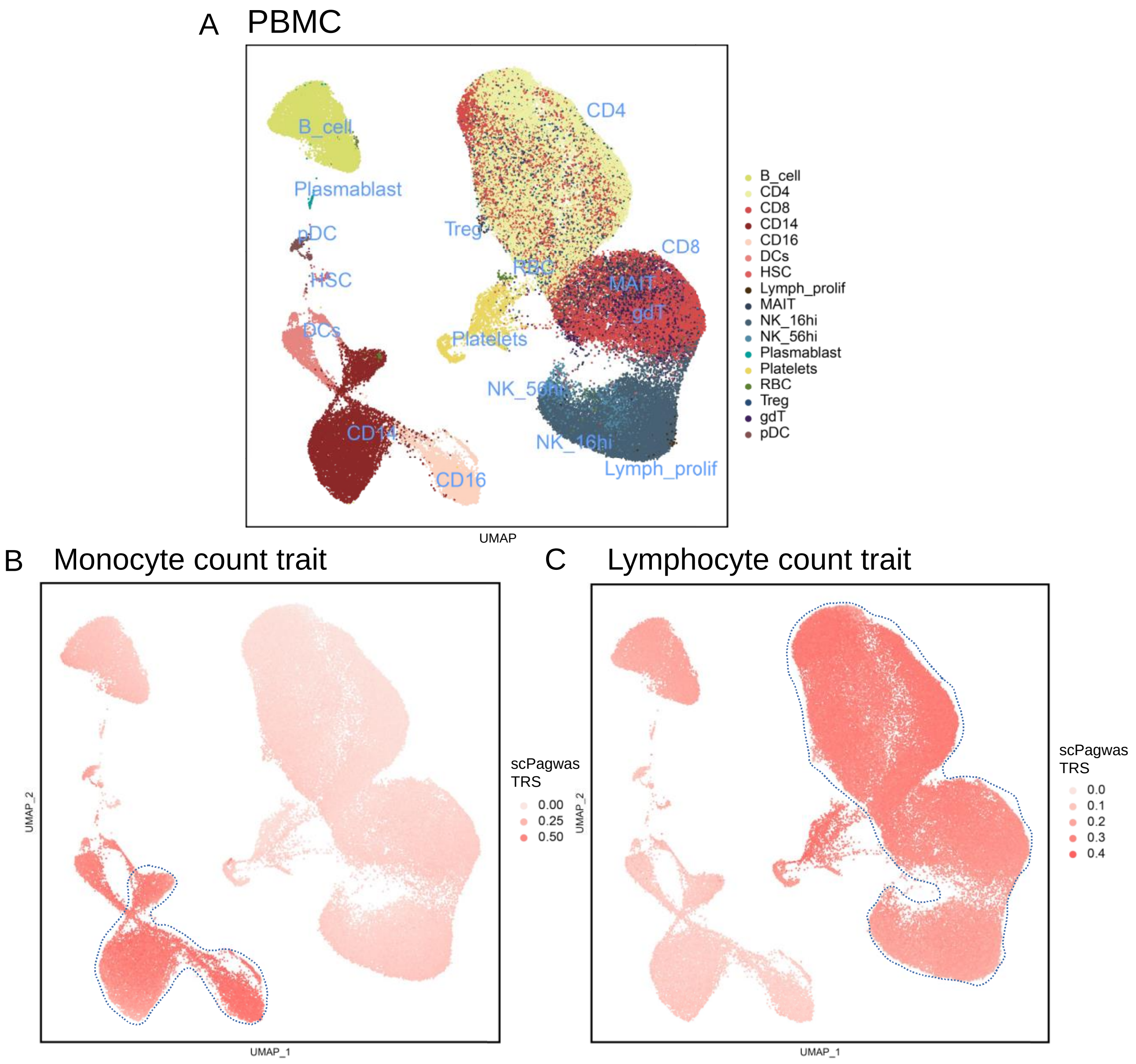


**Supplementary Figure S10. Validation evidence demonstrating that scPagwas (Seurat) captures the genetic associations of two blood cell traits with their relevant cell types, related to Figure 4.** A. The UMAP plot shows the cell type labels in a PBMC scRNA-seq dataset. B. Per-cell scPagwas TRS based on top 1,000 genes for the representative trait of monocyte count trait. C. Per-cell scPagwas TRS based on top 1,000 genes for the representative trait of lymphocyte count trait. Note: The analysis was performed based on the PBMC scRNA-seq dataset (n = 97,039 cells, see Supplementary Table S2). The cell counts of these cell types were listed as follow: B cell (n = 7,568 cells), CD4+T cells (n = 28,268 cells), CD8+T cells (n = 18,796 cells), CD14+T cell (n = 10,312 cells), CD16+mononcyte (n = 3,466), DCs (n = 2,115 cells), HSC (n = 144 cells), lymphocyte_prolif (n = 282 cells), MAIT (n = 2,014 cells), mono_prolif (n = 4 cells), NK_16hi (n = 12,640 cells), NK_56hi (n = 2,222 cells), Plasmablast (n = 115 cells), platelets (n = 2,145 cells), RBC (n = 327 cells), Treg (n = 2,340), gdT (n = 3,592 cells), and pDC (n = 689 cells).

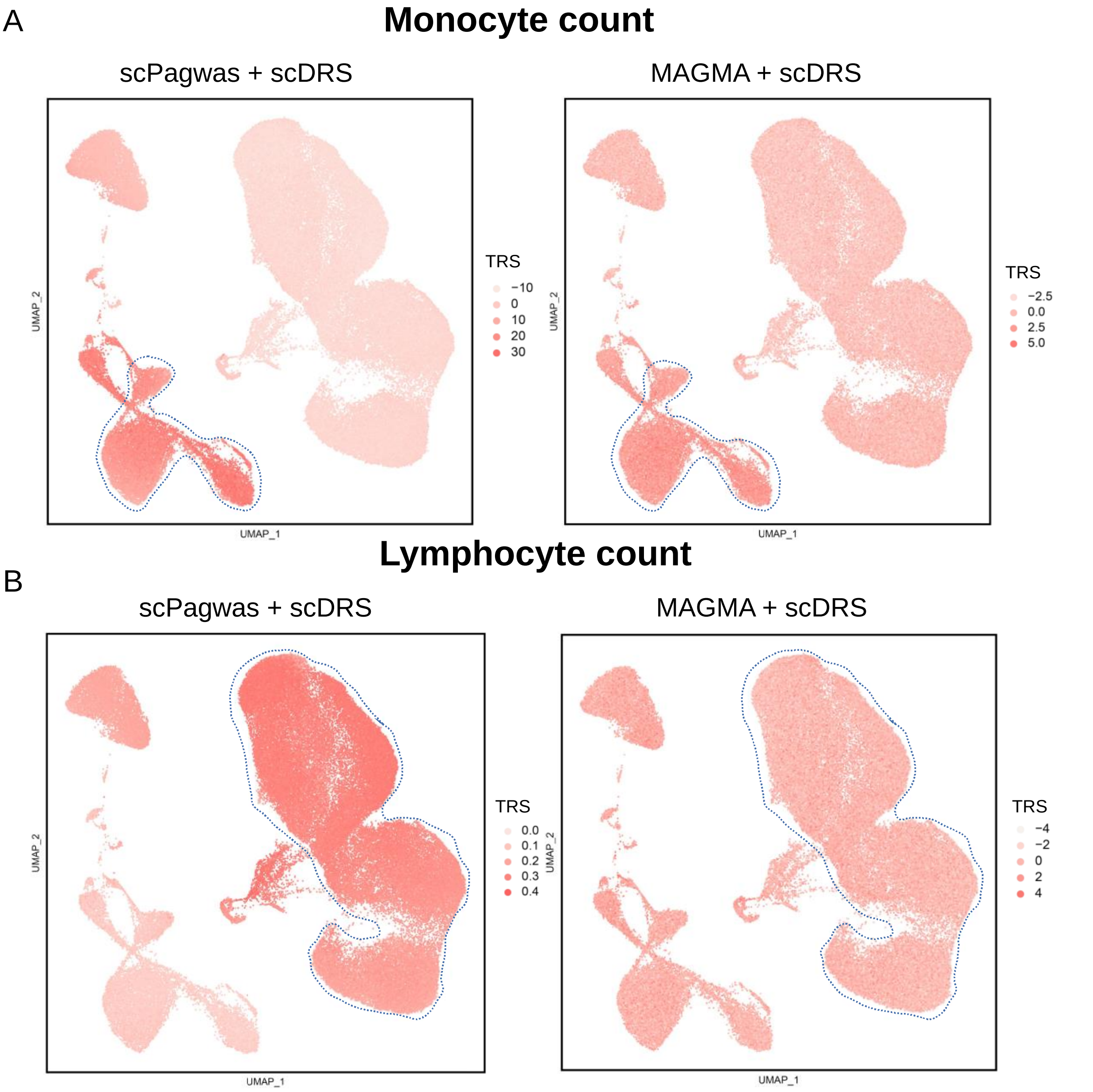


**Supplementary Figure S11. Illustration of per-cell scDRS TRS for two representative blood cell traits in the PBMC scRNA-seq dataset, related to Figure 4.** The UMAP embedding plots showing the results of scDRS using top scPagwas 1,000 genes (left panel) and using top MAGMA 1,000 genes (right panel) for lymphocyte count (A) and monocyte count (B). Note: The analysis was performed based on the PBMC scRNA-seq dataset (n = 97,039 cells, see Supplementary Table S2). The cell counts of these cell types were listed as follow: B cell (n = 7,568 cells), CD4+T cells (n = 28,268 cells), CD8+T cells (n = 18,796 cells), CD14+T cell (n = 10,312 cells), CD16+mononcyte (n = 3,466), DCs (n = 2,115 cells), HSC (n = 144 cells), lymphocyte_prolif (n = 282 cells), MAIT (n = 2,014 cells), mono_prolif (n = 4 cells), NK_16hi (n = 12,640 cells), NK_56hi (n = 2,222 cells), Plasmablast (n = 115 cells), platelets (n = 2,145 cells), RBC (n = 327 cells), Treg (n = 2,340), gdT (n = 3,592 cells), and pDC (n = 689 cells).

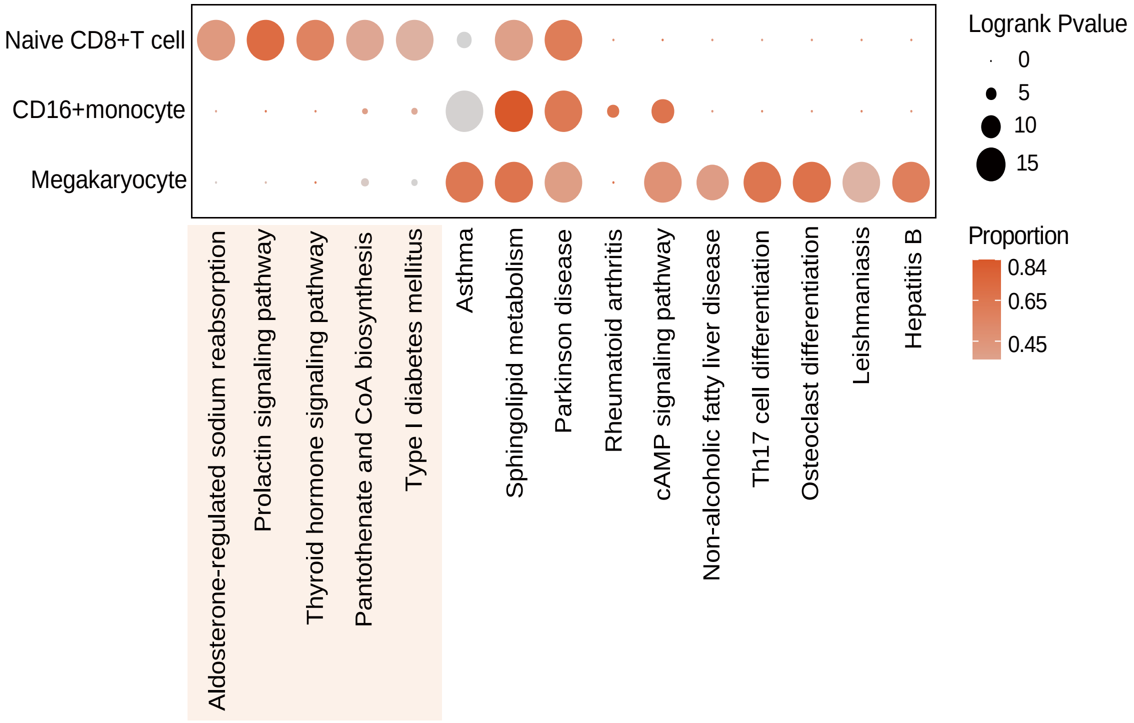


**Supplementary Figure S12. Dot plot showing the trait-risk pathways across three significant cell types by scPagwas among severe COVID-19 patients, related to Figure 5.** Note: scPagwas identified three cell types of naïve CD8+T cell, CD16+monocyte, and megakaryocyte significantly associated with severe COVID-19 by incorporating a large-scale GWAS summary dataset on severe COVID-19 (N = 969,689 samples, Supplementary Table S1) with a large PBMC scRNA-seq dataset (N = 469,453 cells, Supplementary Table S2). By adopting the central limit theorem, we identified top-ranked significant trait-relevant risk pathways with higher gPAS scores. Dot size represents the log-ranked P value for each pathway, and color intensity represents the proportion of cells within each cell type genetically influenced by a given pathway (i.e., pathway-level coefficient beta > 0, indicating that pathway-level genetic effects on cells).

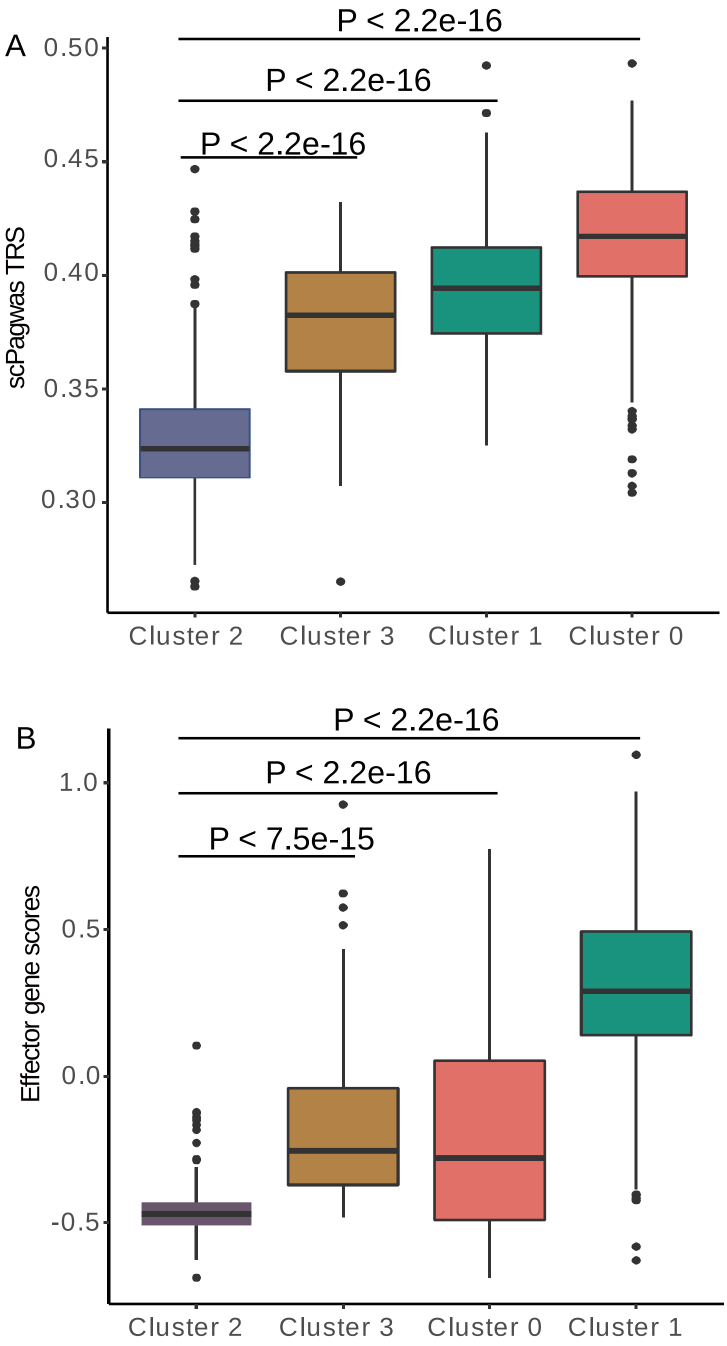


**Supplementary Figure S13. Boxplots showing the TRS and effector gene score among four cell clusters in naïve CD8+T cells, related to Figure 5.** A. Cells with scPagwas TRSs among four cell clusters. B. Cells with effector marker gene scores among four cell clusters. The effector marker gene scores were calculated by using the *AddModuleScore* function in Seurat (see Supplementary Methods). These effector marker genes are shown in Supplemental Table S5. The statistical significance among different cell clusters was calculated by using the two-sided Student' T test.

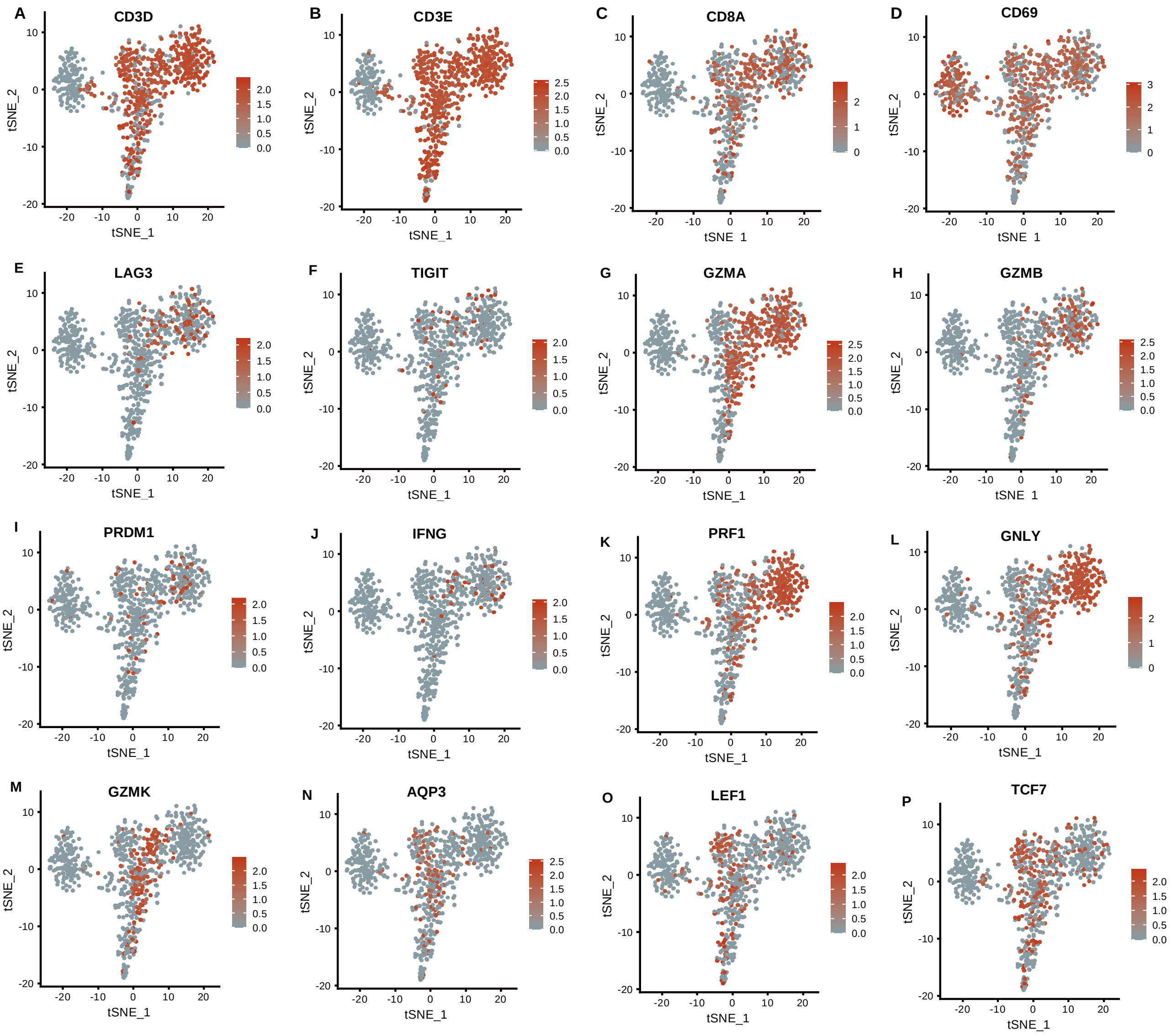


**Supplementary Figure S14. Feature plots demonstrating expression of important marker genes in 766 naïve CD8+T cells, related to Figure 5.** A-P) Color scale represents the scaled gene expression value (log10 scale) for each cell for a given marker gene. Note: CD3D, CD3E and CD8A represent markers for CD8+T cells, LAG3 and TIGIT represent exhaustion markers, GZMA, GZMB, PRDM1, IFNG, PRF1, and GNLY represent effector marker genes, GZMK, AQP3, and CD69 represent memory markers, LEF1 and TCF7 represent naïve markers.

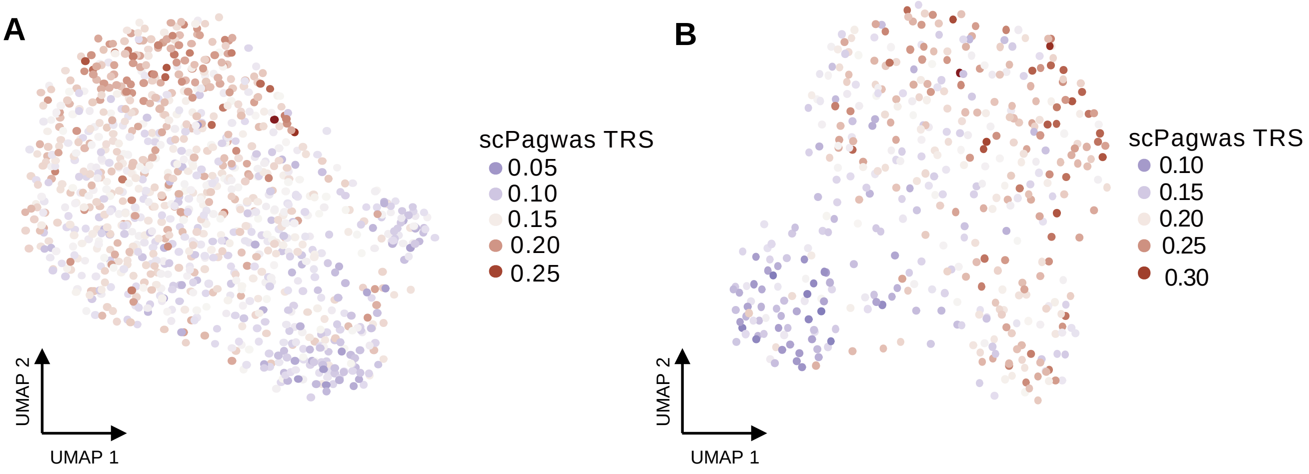


**Supplementary Figure S15. Associations of OPC and microglial cells with AD based on the human brain entorhinal cortex snRNA-seq dataset, related to Figure 6.** A. UMAP visualization of OPC cells with scPagwas TRSs. Using the Geary' C method (see Supplemental Methods), we examined the spatial autocorrelation of TRS across cells in the cell type of OPC. We found that the remarkable heterogeneous AD-associations among cells in OPC (FDR = 3.33e-4). The statistical significance was calculated by using the *getSignatureAutocorrelation*() function in VISION R package with default parameters. B. UMAP visualization of microglial cells with scPagwas TRSs. Color intensity represent the degree of scPagwas TRS for each cell. Cells with red color indicate a strong enrichment by genetic association signals of AD.

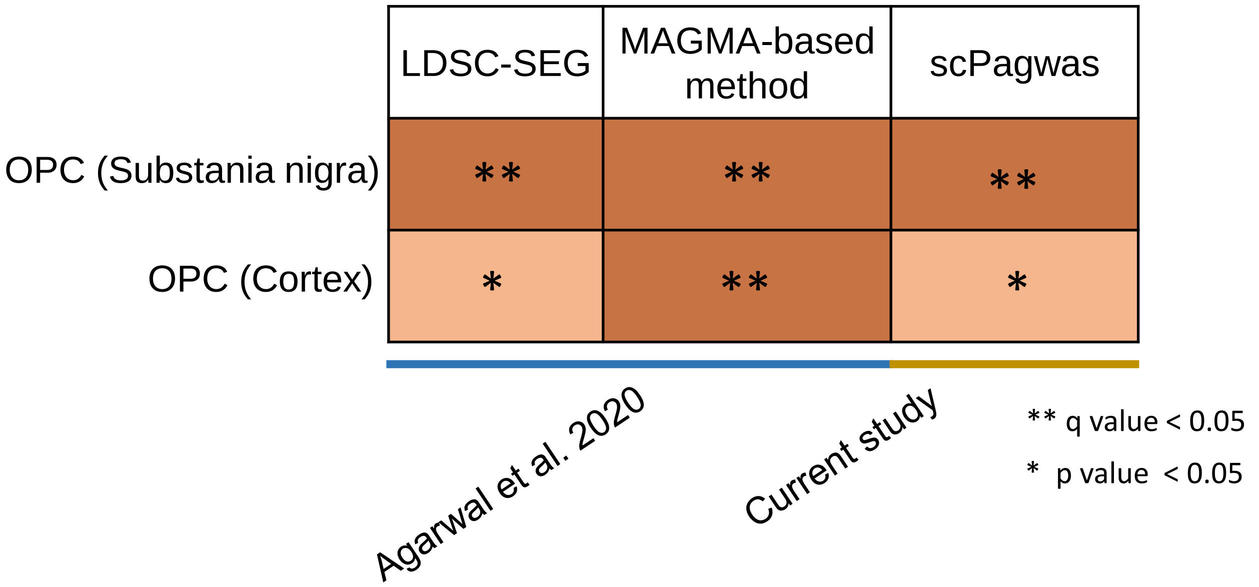


**Supplementary Figure S16. Validation of the association of OPC with schizophrenia by using scPagwas.** The human brain snRNA-seq dataset reported by Agarwal et al. 2020 was downloaded from the GEO database (Accession number: GSE140231). In the original paper, the authors used two methods of LDSC-SEG and MAGMA-based approach for identifying schizophrenia-associated brain cell types, and found that the OPC cell type was significantly associated with schizophrenia in both substania nigra and cortex brain regions. In the current study, we applied scPagwas to validate whether OPC is associated with schizophrenia in the same scRNA-seq dataset. The GWAS summary statistics on SCZ was based on the wave 3 of the Psychiatric Genomics Consortium (n = 161,405 samples with 67,390 cases and 94,015 controls, see Supplementary Table S1). The heatmap colors give different degrees of significance with each method, an asterisk (*) and double asterisks (**) indicate nominally significant p value (< 0.05) and q value (Bonferroni correction for each raw p value).

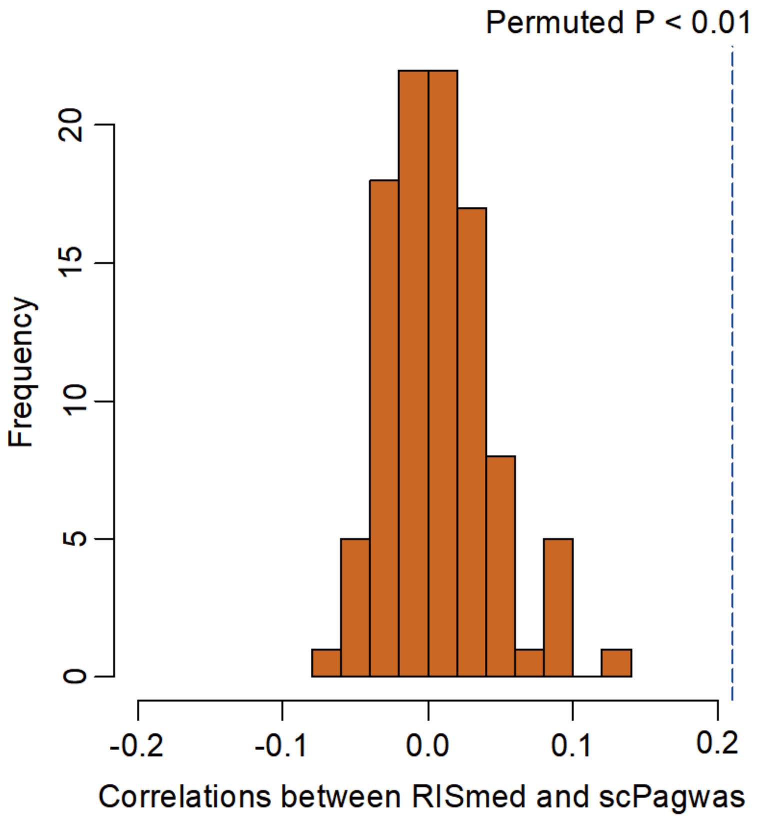


**Supplementary Figure S17. Permutation analysis for assessing the correlation between top 1,000 scPagwas-identified AD-relevant genes and RISmed-searching evidence counts, related to Figure 6F.** Note: We performed an *in silico* permutation analysis of 100 times by randomly selecting length-matched genes (n = 1,000 genes) from background genes and calculating their correlations with RISmed-searching results. Barplot shows the frequency of the correlation coefficients between RISmed-seaching results and  results from random selections, and blue vertical dotted-line indicates the observed correlation coefficient from scPagwas-identified AD-relevant genes. Please see the Supplementary Methods for detailed information.

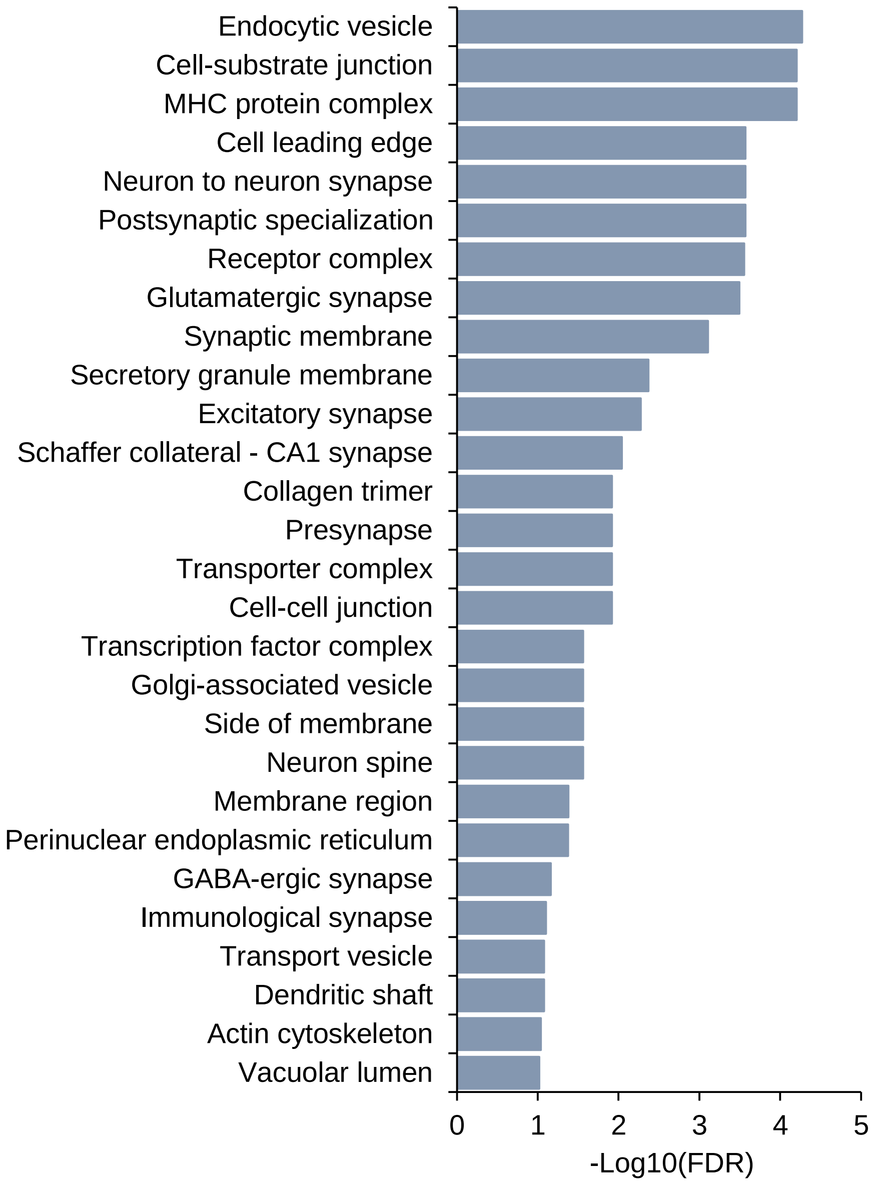


**Supplementary Figure S18. GO-term enrichment analysis using cellular component terms of top-ranked 1,000 risk genes for AD, related to Figure 6E.** Note: This enrichment analysis was performed by using the web-accessed tool of WebGestlat (http://www.webgestalt.org/). There were 28 significant GO-terms enriched by these top-ranked 1,000 risk genes for AD (FDR < 0.1, see Supplementary Table S8).

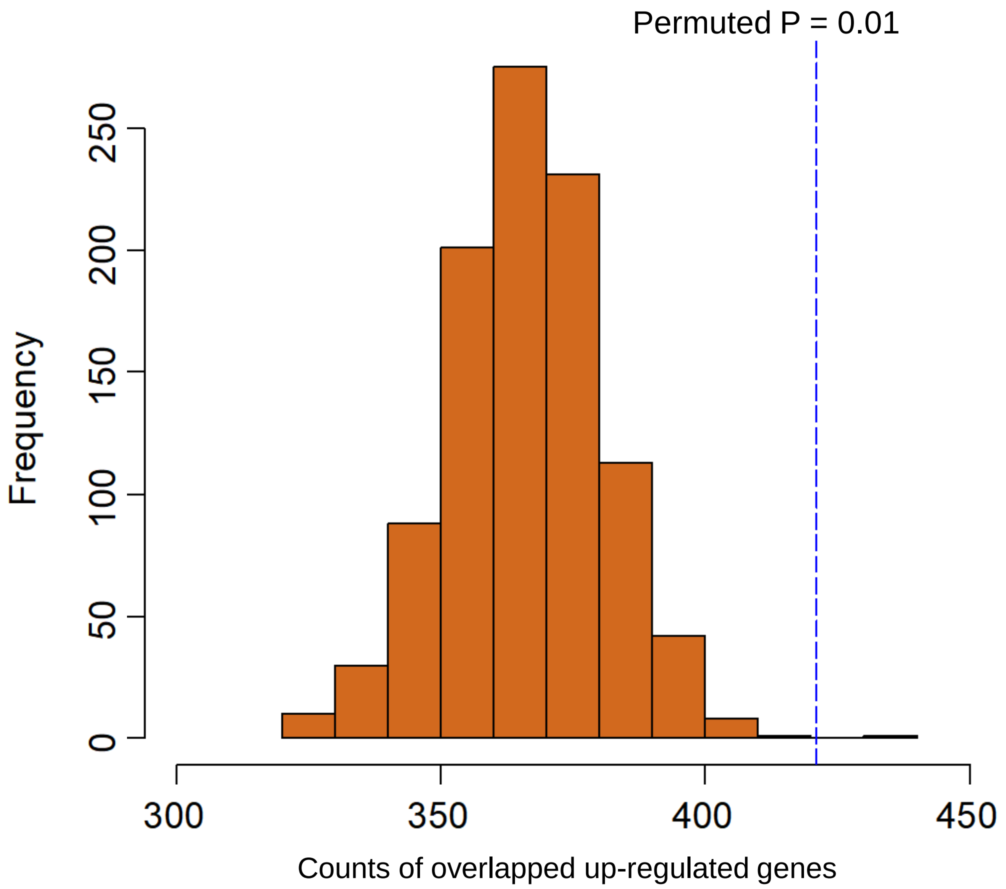


**Supplementary Figure S19. Permutation analysis for assessing the significance of top 1,000 scPagwas-identified AD-relevant genes that were up-regulated in both bulk RNA expression datasets on AD.** Note: We performed an *in silico* permutation analysis (n = 1,000 times) by randomly selecting length-matched genes (n = 1,000 genes) to count the overlapped genes that were significantly up-regulated in two bulk RNA-expression datasets on AD (Accession number: GSE15222 and GSE109887), and compare with the number of top scPagwas-identified AD-relevant genes that were significantly higher expressed in these two bulk datasets. Barplot shows the frequency of overlapped up-regulated genes from random selections, and blue vertical dotted-line indicates the observed overlapped up-regulated genes from scPagwas-identified AD-relevant genes. Please see the Supplementary Methods for detailed information.
