## Supplemental Tables for "Polygenic regression uncovers trait-relevant cellular contexts through pathway activation transformation of single-cell RNA sequencing data"

### *Supplementary Tables*

**Supplementary Table S1. Summary of GWAS summary statistics on 13 complex diseases and traits used in the current study**

| Phenotypes | Year | Data ID | Population | Sample size | Number of SNPs | Resources |
|---|---|---|---|---|---|---|
| Monocyte count | 2020 | ieu-b-31 | European | 563,946 | 26,946,895 | https://gwas.mrcieu.ac.uk/datasets/ieu-b-31/ |
| Lymphocyte count | 2016 | ebi-a-GCST004627 | European | 171,643 | 29,167,587 | https://gwas.mrcieu.ac.uk/datasets/ebi-a-GCST004627/ |
| Lymphocyte percent of white cells | 2016 | ebi-a-GCST004632 | European | 171,748 | 29,167,359 | https://gwas.mrcieu.ac.uk/datasets/ebi-a-GCST004632/ |
| Mean corpus volume | 2020 | ebi-a-GCST90002335 | European | 121,047 | 14,991,118 | https://gwas.mrcieu.ac.uk/datasets/ebi-a-GCST90002335/ |
| Neutrophil count | 2020 | ieu-b-34 | European | 563,946 | 26,935,743 | https://gwas.mrcieu.ac.uk/datasets/ieu-b-34/ |
| White blood cell count | 2020 | ieu-b-30 | European | 563,946 | 27,090,906 | https://gwas.mrcieu.ac.uk/datasets/ieu-b-30/ |
| Eosinophil count | 2020 | ieu-b-33 | European | 563,946 | 26,776,110 | https://gwas.mrcieu.ac.uk/datasets/ieu-b-33/ |
| Basophil count | 2020 | ieu-b-29 | European | 563,946 | 26,797,882 | https://gwas.mrcieu.ac.uk/datasets/ieu-b-29/ |
| Mean corpuscular hemoglobin concentration (MCHC) | 2020 | ebi-a-GCST90002329 | East Asian | 126,151 | 14,939,486 | https://gwas.mrcieu.ac.uk/datasets/ebi-a-GCST90002329/ |
| Hemoglobin concentration | 2020 | ebi-a-GCST90002311 | East Asian | 149,861 | 15,410,015 | https://gwas.mrcieu.ac.uk/datasets/ebi-a-GCST90002311/ |
| Hospitalized COVID-19 | 2021 | B2_ALL_leave_23andme | European | 969,689 | 9,368,170 | https://www.covid19hg.org/ |
| Alzheimer's disease | 2019 | ieu-b-2 | European | 63,926 | 10,528,610 | https://gwas.mrcieu.ac.uk/datasets/ieu-b-2/ |
| Schizophrenia | 2022 | PGC3_SCZ_wave3 | European (~80%) | 161,405 | 7,585,078 | https://figshare.com/articles/dataset/scz2022/19426775 |

**Supplementary Table S2. Summary of eight single-cell sequencing datasets spanning 1,404,556 cells from human and mouse used in the current study**

| Data set | Species | Number of cells | Number of cell types | Tissues /Organ | Description | Resources |
|---|---|---|---|---|---|---|
| Granjia et al. | Human | 35,582 | 24 | BMMC | This dataset contains the full spectrum of human hematopoietic differentiation from | https://jeffgranja.s3.amazonaws.com/MPAL-10x/Supplementary_Data/He |

| | | | | | | |
|---|---|---|---|---|---|---|
| | | | | | stem cells to their corresponding progenies | althy-Data/scRNA-Healthy-Hematopoiesis-191120.rds |
| Stephenson et al. | Human | 97,039 | 17 | PBMC | 24 healthy individuals with PBMC samples, using the 10X Genomics scRNA-seq platform | https://www.ebi.ac.uk/arrayexpress/experiments/E-MTAB-10026/ |
| Han et al. | Human | 513,707 | 60 | 35 adult tissues | Using microwell-seq platform to perfom scRNA-seq for determining the cell type composition of all major human organs | https://www.ncbi.nlm.nih.gov/geo/query/acc.cgi?acc=GSE134355 |
| Bryois et al. | Mouse | 160,796 | 39 | Brain | Single cell suspensions of all brain regions, the majority of sampling was sequenced by using the 10X Genomics scRNA-seq platform | https://storage.googleapis.com/linnarsson-lab-loom/l5_all.loom |
| Grubman et al. | Human | 11,786 | 5 | Brain | One region of entorhinal cortex from control and AD brains of 12 individuals, the Illumina HiSeq 500 according to the 10X Gnomics protocol for snRNA-seq | https://www.ncbi.nlm.nih.gov/geo/query/acc.cgi?acc=GSE138852 |
| Smith et al. | Human | 101,906 | 9 | Brain | Two brain regions of entorhinal and somatosensory cortex from 6 non-disease control and 6 AD cases, the Illumina HiSeq 4000 according to the 10X Gnomics protocol for snRNA-seq | https://www.ncbi.nlm.nih.gov/geo/query/acc.cgi?acc=GSE160936 |
| Agarwal et al. | Human | 14,287 | 8 | Brain | The first human single-nuclei RNA-seq for substantia nigra and matched cortical brain regions from 12 samples The 10X chromium system (V2) was used for single-cell sequencing | https://www.ncbi.nlm.nih.gov/geo/query/acc.cgi?acc=GSE140231 |

| | | | | | | |
|---|---|---|---|---|---|---|
| Su et al. | Human | 469,453 | 13 | PBMC | Containing the major immune cell classes in COVID-19 patients, 10X Genomics scRNA-seq platform | https://www.ebi.ac.uk/arrayexpress/experiments/E-MTAB-9357 |

**Supplementary Table S3. GO-term (biological process) enrichment analysis of top-ranked 1,000 genes from scPagwas and MAGMA (FDR < 0.01) for 10 blood cell traits**

| Blood cell trait | GO-terms enriched by scPagwas top 1,000 genes (FDR < 0.01) | GO-terms enriched by MAGMA top 1,000 genes (FDR < 0.01) | GO-terms enriched by TWAS top 1,000 genes (FDR < 0.01) | GO-terms enriched by S-PrediXcan top 1,000 genes (FDR < 0.01) | GO-terms enriched by S-MultiXcan top 1,000 genes (FDR < 0.01) |
|---|---|---|---|---|---|
| Monocyte count | GO:0036230, GO:0002446, GO:0002521, GO:0002764, GO:0002694, GO:0042110, GO:0030099, GO:0050867, GO:0006909, GO:0034341, GO:0019882, GO:0071706, GO:0007159, GO:0001819, GO:0007015, GO:0031349, GO:0045088, GO:0002250, GO:1903706, GO:0042113, GO:0045785, GO:0070661, GO:0051051, GO:0022407, GO:0060627, GO:0032970, GO:1901652, GO:0002237, GO:0050727, GO:1902903, GO:0071216, GO:0051258, GO:0051493, GO:0007249, GO:0072593, GO:0002449, GO:0002697, GO:0051090, GO:0052547, GO:0022604, GO:0043254, GO:0002576, GO:0001818, GO:0045576, GO:0032635, GO:2001233, GO:0002532, GO:0071887, GO:0032637, GO:0032612, GO:0032103, GO:0050817, GO:0032418, GO:0006898, GO:0050900, GO:0002448, GO:0072524, GO:0002285, GO:0061919, GO:0098542, GO:0050663, GO:0071900, GO:0090066, GO:0007229, GO:0016052, GO:0031589, GO:0007162, GO:0051098, GO:0042116, GO:0032606, GO:0002683, GO:0042107, GO:0072512, GO:2000147, GO:0042326, GO:1902532, GO:0090662, GO:0009141, GO:0035690, GO:0032633, GO:0034109, GO:0002440, GO:0071496, GO:0032602, GO:0050878, GO:1904951, GO:0033002, GO:0070671, GO:0032609, GO:2001057, GO:0060326, GO:0042092, GO:0010959, GO:0000041, GO:0097193, GO:0050866, GO:0010324, GO:0032615, GO:0048872, GO:0006979, | GO:0006909, GO:0050900, GO:0002521, GO:0030099, GO:0007159, GO:0042110, GO:1903706, GO:0045785, GO:0042113, GO:0060326, GO:0060249, GO:0002683, GO:0060627, GO:0046777, GO:0070741, GO:1990868, GO:0006968, GO:0002694, GO:0048872, GO:0071900, GO:0050867, GO:0036230, GO:0043254, GO:0048017 | Null | GO:0006968 | GO:0034341, GO:0019882, GO:0060326, GO:0007159, GO:0050900, GO:1990868, GO:0006968, GO:0002764, GO:0006909, GO:0032103, GO:0042110, GO:0050920, GO:0030099 |

| | | | | | |
|---|---|---|---|---|---|
| | GO:0051346, GO:0031529, GO:0007568, GO:0048013, GO:0032613, GO:0150076, GO:0045862, GO:0007033, GO:0070997, GO:0038061, GO:0070371, GO:0050690, GO:0140029, GO:0031098, GO:0099132, GO:0043112, GO:0007265, GO:0060759, GO:0061351, GO:0045730, GO:0048010, GO:0051047, GO:0031348, GO:0002269, GO:0032386, GO:0009636, GO:0032623, GO:1902600, GO:0002791, GO:0043900, GO:0104004, GO:0071800, GO:0051235, GO:0009123, GO:1901136, GO:0097305, GO:1990778, GO:0097191, GO:0003012, GO:0051348, GO:0022406, GO:0006090, GO:0046677, GO:0001906, GO:0046939, GO:0016197, GO:0009595, GO:0001667, GO:0045861, GO:0031099, GO:0034404, GO:0070482, GO:0051701, GO:0016050, GO:0018212, GO:0097581, GO:0006968, GO:0051156, GO:0006091, GO:0009615, GO:0090130, GO:0009620, GO:0009743, GO:0016049, GO:0019058, GO:0034330, GO:0097006, GO:0031532, GO:0016051, GO:0046434, GO:0009132, GO:0051271, GO:0010573, GO:0006732, GO:0038127, GO:0034340, GO:0006907, GO:0031579, GO:0097242, GO:0048771, GO:1904950, GO:0055076, GO:1901342, GO:0042063, GO:0001773, GO:0006959, GO:0031647, GO:0005996, GO:0050769, GO:0001525, GO:0010975, GO:0071241, GO:0032147, GO:0070849, GO:0048017, GO:1905475, GO:0009308, GO:0032102 | | | | |
| Mean Corpuscular Hemoglobin | GO:0044772, GO:0048285, GO:0051052, GO:1901987, GO:0045787, GO:0007059, GO:0010948, GO:0006310, GO:0006260, GO:0045930, GO:0044843, GO:0071103, GO:0000075, GO:0044839, GO:1902850, GO:0033044, GO:0051321, GO:0007051, GO:0071824, GO:0010639, GO:0034502, GO:0006333, GO:0061641, GO:0000910, GO:0051383, GO:0032200, GO:0006338, GO:0006302, GO:0042110, GO:0050000, GO:0002200, GO:0031023, GO:0072331, GO:0042770, GO:1904029, GO:0071897, GO:0050867, GO:0032886, GO:0002521, GO:2001020, | Null | GO:0036230, GO:0002446 | Null | Null |

| | | | | | |
|---|---|---|---|---|---|
| | GO:0051098, GO:0060249, GO:0032069, GO:0009262, GO:0002694, GO:1903829, GO:0051302, GO:0022407, GO:0051493, GO:0022616, GO:0007050, GO:0045785, GO:0048872, GO:0002250, GO:0007159, GO:0007018, GO:0045023, GO:0006298, GO:0042769, GO:0036297, GO:0006289, GO:0019692, GO:0051169, GO:0006403, GO:0040029, GO:0042113, GO:0006997, GO:0008380, GO:0009112 | | | | |
| Neutrophil count | GO:0036230, GO:0002446, GO:0002521, GO:0002764, GO:0002694, GO:0006909, GO:0019882, GO:0042110, GO:0007015, GO:0034341, GO:0051051, GO:0050867, GO:0002250, GO:0045088, GO:0042113, GO:0030099, GO:0060627, GO:0007159, GO:0071706, GO:0031349, GO:0032970, GO:0070661, GO:0001819, GO:1903706, GO:0051493, GO:0045785, GO:1902903, GO:0002237, GO:0022407, GO:0002449, GO:0001818, GO:1901652, GO:0051090, GO:0007249, GO:0051258, GO:0052547, GO:0006898, GO:0002697, GO:0050727, GO:0071216, GO:0002576, GO:0072593, GO:0045576, GO:0032635, GO:0002683, GO:0043254, GO:0022604, GO:2001233, GO:0032606, GO:0010324, GO:0007162, GO:0070671, GO:0007229, GO:0002440, GO:0002285, GO:0050817, GO:0090066, GO:1904950, GO:0050900, GO:0071887, GO:0051098, GO:0032418, GO:0072524, GO:0009141, GO:0010959, GO:0002448, GO:0031589, GO:0042116, GO:0042326, GO:0050866, GO:0032103, GO:0032612, GO:0042107, GO:0045862, GO:1902532, GO:0002532, GO:0034109, GO:0048010, GO:0032623, GO:0050663, GO:0050690, GO:0050878, GO:1904951, GO:0061919, GO:0016052, GO:0031348, GO:0032386, GO:0097193, GO:0071900, GO:0098542, GO:0009123, GO:1990778, GO:0033002, GO:0043112, GO:2000147, GO:0006091, GO:0048013, GO:0032613, GO:0150076, GO:0032633, GO:0002791, GO:0007033, GO:0048872, GO:0051346, GO:0070997, GO:0035690, GO:0140029, GO:0045730, | GO:0048872, GO:0002521, GO:0071216, GO:0006909, GO:0071887, GO:1903706, GO:0050900, GO:0042110, GO:0007159, GO:0006338, GO:2001233, GO:0002683, GO:0045785, GO:0030099, GO:0002694 | Null | GO:0050900 | GO:0019882, GO:0034341, GO:0001819, GO:0002764, GO:0002697, GO:0001906, GO:0002683 |

| | |
|---|---|
| | GO:0043900, GO:0032615, GO:0002269, GO:1905475, GO:0032602, GO:0051348, GO:0060326, GO:0031529, GO:0032637, GO:0007568, GO:0003012, GO:1902600, GO:2001057, GO:0090662, GO:0006907, GO:0071800, GO:0097242, GO:0034330, GO:0051235, GO:0060759, GO:0042176, GO:0061351, GO:0097191, GO:0016049, GO:0070371, GO:0022406, GO:0007265, GO:0031532, GO:0001906, GO:0016197, GO:0032609, GO:1903829, GO:0071496, GO:0009615, GO:0051047, GO:0019058, GO:0034340, GO:0007034, GO:0031099, GO:0038061, GO:0038127, GO:0051701, GO:0045861, GO:0016050, GO:0006959, GO:0097581, GO:0018212, GO:0006968, GO:0051156, GO:0031098, GO:0006979, GO:0001667, GO:0006090, GO:0097006, GO:0009636, GO:0008637, GO:0046939, GO:0034612, GO:0051271, GO:0042063, GO:0099132, GO:0046677, GO:0014812, GO:0031579, GO:1903828, GO:0043270, GO:0010821, GO:0048771, GO:0009259, GO:0009266, GO:0042092, GO:0009743, GO:0055076, GO:0032102, GO:0098876, GO:1901342, GO:0001773, GO:0046434, GO:0030865, GO:0072512, GO:0097305, GO:0034404, GO:0009132, GO:0001525, GO:0009896, GO:0104004, GO:1901136, GO:0090130, GO:0070849 |

| | | | | | |
|---|---|---|---|---|---|
| Hemoglobin concentration | GO:0036230, GO:0002446, GO:0044772, GO:0007059, GO:0006260, GO:0071103, GO:0009141, GO:0051052, GO:0009123, GO:0006091, GO:1901987, GO:0033044, GO:0048285, GO:0044843, GO:0006338, GO:0044839, GO:0071824, GO:0032200, GO:0006403, GO:0006333, GO:0034502, GO:0140053, GO:0010948, GO:0006310, GO:0045787, GO:0033108, GO:0071897, GO:0010257, GO:1902850, GO:0006414, GO:0051383, GO:0061641, GO:0019882, GO:0009259, GO:0000075, GO:0032984, GO:0007051, GO:0051169, GO:0010639, GO:0008380, GO:0045930, GO:0050000, GO:0006397, GO:0006979, GO:0006457, GO:0015931, GO:0050867, GO:1904951, GO:0060249, GO:0045023, GO:0042769, GO:0072593, GO:0051321, GO:0016999, GO:1903829, GO:0006302, GO:0009636, GO:0009262, GO:0042770, GO:1901657, GO:2001020, GO:0046677, GO:0006839, GO:0006909, GO:0010608, GO:0070585, GO:0015949, GO:0009895, GO:0006081, GO:0034404, GO:0048144, GO:0070646, GO:1990823, GO:0008637, GO:0006401, GO:0022613, GO:0018205, GO:0043687, GO:0072524, GO:0022616, GO:0034504, GO:0070661, GO:0040029, GO:0051098, GO:0016569, GO:0044282, GO:0045454, GO:0002764, GO:1904029, GO:0000910, GO:2001233, GO:0010498, GO:0071166, GO:0071706, GO:0019692, GO:0072331, GO:1903311, GO:0097066, GO:0031647, GO:0042113, | GO:0048706, GO:0048568, GO:0002521, GO:0048872, GO:0061448, GO:0061614, GO:0033044 | Null | Null | GO:0019882 |

| | | | | | |
|---|---|---|---|---|---|
| Basophil count | GO:0036230, GO:0002446, GO:0002521, GO:0002764, GO:0002694, GO:0042110, GO:0031349, GO:0001819, GO:0002250, GO:0030099, GO:0045088, GO:0007159, GO:0050867, GO:0006909, GO:0034341, GO:0002237, GO:1903706, GO:0070661, GO:0045785, GO:0007015, GO:0022407, GO:0019882, GO:0071216, GO:0071706, GO:0042113, GO:0051051, GO:0007249, GO:0002697, GO:0060627, GO:0001818, GO:1901652, GO:0032970, GO:0002285, GO:0050727, GO:0051090, GO:0098542, GO:0032635, GO:0045576, GO:0032103, GO:0071900, GO:0002683, GO:0022604, GO:0050900, GO:0002449, GO:0050663, GO:1902903, GO:0051493, GO:0071496, GO:0002576, GO:2000147, GO:0002532, GO:0072593, GO:0050817, GO:0007162, GO:0051258, GO:0050866, GO:0032612, GO:0032606, GO:2001233, GO:0002448, GO:0052547, GO:0007229, GO:0032637, GO:0042107, GO:0032418, GO:1902532, GO:0071887, GO:0042116, GO:0070482, GO:0097193, GO:0006898, GO:0060326, GO:0051098, GO:0043254, GO:0050878, GO:0072524, GO:0090066, GO:0031589, GO:0042326, GO:0032602, GO:0070997, GO:0001667, GO:0070371, GO:0090130, GO:0006979, GO:0033002, GO:0061919, GO:0032615, GO:0010959, GO:0035690, GO:0032633, GO:0032613, GO:0150076, GO:0002440, GO:0007265, GO:0048013, GO:0048872, GO:0002791, GO:0048771, GO:0042092, GO:0045730, GO:0046677, GO:0031098, GO:0009615, GO:0038061, GO:1904951, GO:0045862, GO:0010324, GO:0071800, GO:0031529, GO:0048010, GO:0060759, GO:0090662, GO:0032609, GO:0016052, GO:2001057, GO:0010573, GO:1904950, GO:0007033, GO:0009595, GO:0051047, GO:0007568, GO:0001773, GO:0009636, GO:0018212, GO:0051056, GO:0034330, GO:0104004, GO:0051235, GO:0031348, GO:0072512, GO:0002269, GO:0048871, GO:0031667, GO:0034340, GO:0032623, GO:0009141, GO:0006968, | GO:0036230, GO:0002446, GO:0030099, GO:0002521, GO:0006909, GO:0048872, GO:0045785, GO:0050900, GO:1903706, GO:0043112 | Null | GO:0036230, GO:0002446 | GO:0019882, GO:0002764, GO:0036230, GO:0034341, GO:0002446, GO:0006909, GO:0051090, GO:0001906, GO:0031349, GO:0030099, GO:0045785, GO:0060326, GO:0050900, GO:0002521, GO:0045088, GO:1903706, GO:0022407, GO:0002237, GO:0009620, GO:0052192, GO:0002683 |

| | | | | | |
|---|---|---|---|---|---|
| | GO:0000041, GO:0043900, GO:0009620, GO:0043112, GO:0035456, GO:0038127, GO:0032147, GO:0051346, GO:0051701, GO:0070671, GO:0140029, GO:0043087, GO:0006959, GO:0032386, GO:0097305, GO:1905475, GO:0099132, GO:0001906, GO:0031532, GO:0043620, GO:0050690, GO:1990845, GO:0034612, GO:0003012, GO:0001525, GO:0045861, GO:0032620, GO:0009612, GO:0042063, GO:0051348, GO:0097581, GO:0061351, GO:0051156, GO:0016049, GO:0046939, GO:0033028, GO:0070265, GO:0031099, GO:0034109, GO:0071241, GO:1901136, GO:0014812, GO:0010038, GO:0001562, GO:0031579, GO:0097242, GO:0010657, GO:0001101, GO:0048017, GO:0016197, GO:0009743, GO:0046777, GO:0071402, GO:0071559, GO:0048545, GO:1901342, GO:0051271, GO:0006090, GO:1903829, GO:0050769, GO:0031032, GO:0034404, GO:0097191, GO:0070555, GO:0010975, GO:0009132, GO:0009123, GO:0072511, GO:0070849, GO:0019058, GO:0032102, GO:0009266, GO:0016050, GO:1990778, GO:0032616 | | | | |
| Lymphocyte percent | GO:0044772, GO:0009141, GO:0071103, GO:0009123, GO:0007059, GO:0071824, GO:0006338, GO:0008380, GO:0006260, GO:0006091, GO:0006333, GO:0033044, GO:0006397, GO:0034502, GO:1901987, GO:0036230, GO:0006457, GO:0002446, GO:0051169, GO:0032200, GO:1902850, GO:0006403, GO:0048285, GO:0071897, GO:0044839, GO:0044843, GO:0010608, GO:0010257, GO:1903829, GO:0009259, GO:0042113, GO:0051383, GO:0019882, GO:0010948, GO:0007051, GO:0061641, GO:0051052, GO:0033108, GO:1903311, GO:0031647, GO:0006310, GO:0045787, GO:0070661, GO:1904951, GO:0034504, GO:0010639, GO:0045023, GO:0002764, GO:0010498, GO:0051098, GO:0042769, GO:0000075, GO:0071166, GO:0009262, GO:0031503, GO:0042176, | GO:0002521, GO:0030099, GO:1903706, GO:0042110, GO:1903829, GO:0042113, GO:0061919, GO:0006909, GO:0002683, GO:0001819, GO:0036230, GO:0010821, GO:0071216, GO:1990868, GO:0002244, GO:0050867, GO:0002237, GO:0022407, GO:1904951, GO:0048872, GO:0002694, GO:0002446, GO:0045785, GO:0070585, GO:0007159, GO:0002285, GO:0097191, | Null | Null | GO:0042110, GO:0002521, GO:0002237, GO:0034341, GO:0002764 |

| | | | | | |
|---|---|---|---|---|---|
| | GO:0006979, GO:0070585, GO:0060249, GO:0015931, GO:0045930, GO:0034248, GO:0009266, GO:0006289, GO:0006081, GO:0045862, GO:0048144, GO:0045454, GO:0043687, GO:0006301, GO:0008637, GO:0046677, GO:0050000, GO:0040029, GO:0006839, GO:0006909, GO:0052547, GO:0019058, GO:0019080, GO:0050867, GO:0070646, GO:0042770, GO:0022616, GO:1901998, GO:0032606, GO:0006401, GO:0016999, GO:0051051, GO:1902579, GO:0032984, GO:0048010, GO:0032418, GO:0019692, GO:0006302, GO:0006354, GO:0090559, GO:0031123, GO:1901657, GO:0072593, GO:0071826, GO:0010821, GO:0002250 | GO:0060759, GO:1901652 | | | |
| White blood cell | GO:0036230, GO:0002446, GO:0002521, GO:0002764, GO:0002694, GO:0007015, GO:0006909, GO:0034341, GO:0019882, GO:0051051, GO:0030099, GO:0045088, GO:0002250, GO:0050867, GO:0042110, GO:0001819, GO:0042113, GO:0031349, GO:0060627, GO:0070661, GO:0071706, GO:0032970, GO:0007159, GO:1903706, GO:0002237, GO:0051493, GO:0002697, GO:1902903, GO:0001818, GO:0002449, GO:1901652, GO:0071216, GO:0045785, GO:0050727, GO:0022407, GO:0051090, GO:0002683, GO:0002576, GO:0051258, GO:0032635, GO:0002440, GO:0050900, GO:0007249, GO:0032103, GO:0022604, GO:0002285, GO:0050817, GO:0072593, GO:0043254, GO:2001233, GO:0006898, GO:0045576, GO:0032606, GO:0052547, GO:1902532, GO:0071900, GO:0090066, GO:2000147, GO:0007162, GO:0002532, GO:0007229, GO:0072524, GO:0051098, GO:0050663, GO:0050878, GO:0042116, GO:0097193, GO:1904950, GO:0032612, GO:0042107, GO:0042326, GO:0071887, GO:0009141, GO:0032418, GO:0048872, GO:0031589, GO:0060326, GO:0002448, GO:1904951, GO:0098542, GO:0050866, GO:0010959, GO:0016052, GO:0010324, GO:0002791, GO:0035690, GO:0034109, GO:0032623, GO:0032602, GO:0032637, GO:0043112, GO:0070997, | GO:0048872, GO:0002521, GO:2001233, GO:0050900, GO:0006909, GO:0042110, GO:0042113, GO:0002694, GO:0030099, GO:0009612, GO:0002237, GO:0097191, GO:0022407, GO:0007159, GO:0001819, GO:0045785, GO:0071216, GO:0097193, GO:0002285, GO:0070482, GO:0071887, GO:0010821 | Null | GO:0006909 | GO:0019882, GO:0034341, GO:0001819, GO:0042110, GO:0070661, GO:0002764, GO:0002697, GO:0006909, GO:0001906, GO:0002694, GO:0022407, GO:0045785, GO:0034340, GO:0007159, GO:0002449, GO:0002521, GO:0048872, GO:0002683, GO:0060326, GO:0006352, GO:0050900, GO:0031349, GO:1903706 |

| | | | | | |
|---|---|---|---|---|---|
| | GO:0031348, GO:0032386, GO:0071496, GO:0038061, GO:0071800, GO:0032615, GO:0070371, GO:0048010, GO:0061351, GO:0060759, GO:0031098, GO:0090130, GO:0061919, GO:0051701, GO:0048013, GO:0007265, GO:0032613, GO:0150076, GO:0043900, GO:0010573, GO:0032633, GO:0051235, GO:0009123, GO:0001667, GO:0070671, GO:0006979, GO:1990778, GO:0070482, GO:0033002, GO:0140029, GO:0038127, GO:1903829, GO:0045862, GO:0045730, GO:0031532, GO:0032609, GO:1905475, GO:0002269, GO:0051348, GO:0001906, GO:0007033, GO:0031529, GO:0016049, GO:0007568, GO:0034340, GO:2001057, GO:0009615, GO:0009620, GO:0090662, GO:0019058, GO:0034330, GO:0051047, GO:0006907, GO:0051346, GO:0048771, GO:0018212, GO:0006090, GO:0046677, GO:0016197, GO:0046939, GO:1902600, GO:0104004, GO:0001773, GO:0050690, GO:0046434, GO:0003012, GO:0031099, GO:0034404, GO:0042063, GO:0051271, GO:0006959, GO:0097191, GO:0097581, GO:0006968, GO:0051156, GO:0022406, GO:0009636, GO:0010975, GO:0035456, GO:0016050, GO:1901342, GO:0009132, GO:0099132, GO:0097305, GO:0006091, GO:1901136, GO:0014812, GO:0050769, GO:1903828, GO:0031579, GO:0097242, GO:0010821, GO:0001525, GO:0009743, GO:0055076, GO:0042092, GO:0045861, GO:0032102, GO:0034612, GO:0040013, GO:0006732, GO:0030865, GO:0009595, GO:0005996, GO:0008637, GO:0009259, GO:0051056, GO:0072512, GO:0097006, GO:0010721, GO:0071241, GO:0043270, GO:0032147, GO:0070849, GO:0048017, GO:0000041, GO:0009308, GO:0031346, GO:0032616 | | | | |
| Lymphocyte count | GO:0002521, GO:0002694, GO:0042110, GO:0022407, GO:0002250, GO:0007159, GO:0050867, GO:0002449, GO:0045785, GO:0070661, GO:0002285, GO:0032609, GO:1903706, GO:0048872, GO:0001819, GO:0002440, GO:0001818, GO:0032633, | Null | GO:0051235 | Null | Null |

| | | | | | |
|---|---|---|---|---|---|
| | GO:0002697, GO:0042113, GO:0002683, GO:0001906, GO:0050727, GO:0002507, GO:0002764, GO:0051056, GO:0002437, GO:0031349, GO:0002200, GO:0050900, GO:0030099, GO:1902532, GO:0050866, GO:0043087, GO:0032623, GO:0032620, GO:0030101, GO:0009615, GO:0032613, GO:0007265, GO:0032103, GO:0051348, GO:0002791, GO:0070670, GO:0032615, GO:0042326, GO:0042107, GO:0019216, GO:0050690 | | | | |
| Eosinophil count | GO:0036230, GO:0002446, GO:0002521, GO:0002764, GO:0002694, GO:0042110, GO:0031349, GO:0001819, GO:0007015, GO:0002250, GO:0030099, GO:0045088, GO:0050867, GO:0006909, GO:0042113, GO:0034341, GO:0019882, GO:0071706, GO:0007159, GO:0070661, GO:0051051, GO:0060627, GO:0002237, GO:1903706, GO:0002697, GO:0032970, GO:1902903, GO:0071216, GO:0045785, GO:0022407, GO:0007249, GO:1901652, GO:0001818, GO:0050727, GO:0032103, GO:0002449, GO:0051493, GO:0071900, GO:0002683, GO:0051258, GO:0032635, GO:0002285, GO:0051090, GO:0022604, GO:0050817, GO:0050900, GO:0002440, GO:0006898, GO:0043254, GO:2000147, GO:0090066, GO:0002576, GO:0072593, GO:0045576, GO:0032606, GO:1902532, GO:0050663, GO:2001233, GO:0042107, GO:0002532, GO:0098542, GO:0042116, GO:0052547, GO:0032637, GO:0031098, GO:0007162, GO:0032612, GO:0060326, GO:0051098, GO:0050878, GO:0071887, GO:0010324, GO:0032418, GO:0072524, GO:0002791, GO:0097193, GO:0071496, GO:0032613, GO:0031589, GO:0042326, GO:0002448, GO:1904950, GO:0048010, GO:0070371, GO:0010959, GO:0006959, GO:0018212, GO:0007265, GO:1904951, GO:0016052, GO:0032615, GO:0007229, GO:0038061, GO:0032633, GO:0050866, GO:0150076, GO:0032623, GO:0032602, GO:0032609, GO:0070997, GO:0045730, GO:0104004, GO:0051235, GO:0048872, GO:0071800, GO:0007568, | GO:0002521, GO:0042110, GO:0002694, GO:0050867, GO:0007159, GO:0002285, GO:0022407, GO:1903706, GO:0030099, GO:0150076, GO:0001773, GO:0050727, GO:0042092, GO:0007162, GO:0097193, GO:0042107, GO:0042113, GO:0002683, GO:0045785, GO:0001819, GO:0032633, GO:0045862, GO:0050900, GO:0009896, GO:0002764, GO:0006338, GO:0002250, GO:0097191, GO:0002440, GO:0032623, GO:0034612, GO:0002244, GO:0061919, GO:0002697, GO:0104004, GO:0071887, GO:0050663, GO:0006909, GO:0038034, GO:0050866, GO:2001233 | Null | Null | GO:0002521, GO:0042110, GO:0002694, GO:0034341, GO:0002449, GO:0050867, GO:0002250, GO:0001819, GO:0002764, GO:1903706, GO:0002697, GO:0002683, GO:0150076, GO:0007159, GO:0050727, GO:0019882, GO:0022407, GO:0045088, GO:0031349, GO:0030099, GO:0002440, GO:0034340, GO:0002269, GO:0045785, GO:0048872, GO:0098542, GO:0097191, GO:0002285, GO:0042113, GO:0031348, GO:0001906, GO:0032103, GO:0042116, GO:0070661, GO:0042107, GO:0032615, GO:0032609, GO:0032633, GO:0002437, GO:0042176, GO:0034612, GO:2001233, GO:0045444 |

| | | | | | |
|---|---|---|---|---|---|
| | GO:0031529, GO:0060759, GO:0090130, GO:0006968, GO:0009615, GO:0048013, GO:0043900, GO:2001057, GO:0001667, GO:0035690, GO:0033002, GO:0042092, GO:0043112, GO:0051056, GO:0051701, GO:0032147, GO:0031348, GO:1905475, GO:0002269, GO:0032386, GO:0009141, GO:0045862, GO:0097581, GO:0051047, GO:0038127, GO:0090662, GO:0034612, GO:0051346, GO:0001562, GO:0006907, GO:0043087, GO:0061919, GO:0140029, GO:0048771, GO:0051348, GO:0001525, GO:0072511, GO:0001906, GO:0031532, GO:0046939, GO:0006979, GO:0009595, GO:0034330, GO:0001773, GO:0050690, GO:0046434, GO:0034340, GO:0000041, GO:0045861, GO:1901342, GO:1990778, GO:0097305, GO:0072512, GO:0046677, GO:0061351, GO:1903829, GO:0051260, GO:0048871, GO:0032616, GO:0051156, GO:0016197, GO:0050769, GO:0016049, GO:0009743, GO:0007033, GO:0009620, GO:0003012, GO:0010975, GO:0006090, GO:0070482, GO:0070671, GO:0009132, GO:0009612, GO:0034404, GO:0099132, GO:0031667, GO:0034109, GO:0097191, GO:0010573, GO:1904019, GO:0031579, GO:0097242, GO:0048017, GO:0002526, GO:0019058, GO:0055076, GO:0034765, GO:0042063, GO:0008037, GO:0005996, GO:0010038, GO:0031032, GO:0071241, GO:0043270, GO:0009123, GO:1901136, GO:0070849, GO:0046777, GO:0009266, GO:0016050, GO:0031346, GO:0051271, GO:0051262, GO:0007163, GO:0006732 | | | | |
| Mean Corpus Volume | GO:0042737, GO:0048872, GO:0051052, GO:0051187, GO:0009636 | GO:0002521, GO:0030099, GO:0034504 | Null | Null | Null |

### Supplementary Table S4. Benchmarking analyses of scPagwas with LDSC, MAGMA, and RolyPoly for severe COVID-19-associated immune cell types

| Cell types | LDSC-SEG P value | MAGMA P value | RolyPoly P value | scPagwas P value | COVID-19 status |
|---|---|---|---|---|---|
| Naïve CD8+T cells | 1.73E-01 | 5.81E-01 | 9.04E-01 | 4.56E-17 | Severe |
| Megakaryocytes | 3.55E-01 | 2.36E-02 | 1.80E-02 | 7.82E-06 | Severe |
| CD16+monocytes | 3.21E-01 | 1.28E-02 | 3.00E-03 | 1.14E-04 | Severe |
| Naïve CD4+T cells | 4.87E-01 | 3.36E-01 | 1.14E-01 | 2.54E-03 | Severe |

| | | | | | |
|---|---|---|---|---|---|
| Naïve B cells | 4.82E-01 | 3.62E-01 | 4.86E-01 | 4.19E-02 | Severe |
| NK | 6.43E-01 | 1.37E-02 | 8.34E-01 | 1.58E-01 | Severe |
| Effector CD8+T cells | 4.57E-01 | 3.70E-01 | 9.80E-01 | 2.78E-01 | Severe |
| Memory CD4 T cells | 2.20E-01 | 6.00E-01 | 9.60E-01 | 6.02E-01 | Severe |
| CD34+Progenitors | 2.42E-01 | 9.38E-01 | 1 | 8.17E-01 | Severe |
| Dendritic cells | 5.75E-01 | 5.36E-01 | 4.10E-02 | 9.65E-01 | Severe |
| CD14+monocytes | 4.37E-01 | 1.21E-01 | 9.69E-01 | 9.99E-01 | Severe |
| Mature B cells | 3.82E-02 | 7.53E-03 | 9.30E-01 | 1 | Severe |
| Memory CD8+T cells | 4.68E-01 | 5.16E-01 | 1.40E-02 | 1 | Severe |
| Naïve CD8+T cells | 1.92E-01 | 7.97E-01 | 7.70E-02 | 1.98E-22 | Moderate |
| Megakaryocytes | 2.54E-01 | 6.79E-02 | 3.60E-02 | 9.46E-07 | Moderate |
| CD16+monocytes | 3.35E-01 | 1.51E-02 | 6.00E-03 | 2.00E-08 | Moderate |
| Naïve CD4+T cells | 5.00E-01 | 5.86E-01 | 8.48E-01 | 9.07E-03 | Moderate |
| Naïve B cells | 5.00E-01 | 4.17E-01 | 8.24E-01 | 3.28E-03 | Moderate |
| NK | 5.67E-01 | 1.18E-02 | 7.67E-01 | 1.48E-01 | Moderate |
| Effector CD8+T cells | 3.72E-01 | 1.73E-01 | 9.54E-01 | 6.54E-02 | Moderate |
| Memory CD4 T cells | 8.39E-02 | 5.48E-01 | 8.20E-01 | 5.25E-01 | Moderate |
| CD34+Progenitors | 7.72E-02 | 7.93E-01 | 9.91E-01 | 9.98E-01 | Moderate |
| Dendritic cells | 4.06E-01 | 7.35E-02 | 9.00E-03 | 9.32E-01 | Moderate |
| CD14+monocytes | 4.42E-01 | 2.15E-01 | 1 | 9.99E-01 | Moderate |
| Mature B cells | 4.35E-01 | 1.57E-02 | 9.87E-01 | 1 | Moderate |
| Memory CD8+T cells | 2.86E-01 | 2.01E-01 | 3.15E-01 | 1 | Moderate |
| Naïve CD8+T cells | 7.77E-01 | 3.15E-02 | 1.53E-01 | 7.54E-20 | Mild |
| Megakaryocytes | 9.38E-02 | 1.71E-01 | 5.20E-02 | 2.52E-02 | Mild |
| CD16+monocytes | 6.31E-02 | 4.27E-02 | 2.90E-02 | 5.39E-06 | Mild |
| Naïve CD4+T cells | 2.43E-01 | 4.06E-02 | 6.90E-01 | 1.51E-03 | Mild |
| Naïve B cells | 4.98E-01 | 8.11E-01 | 9.58E-01 | 6.00E-02 | Mild |
| NK | 7.47E-01 | 4.60E-02 | 6.82E-01 | 5.14E-03 | Mild |
| Effector CD8+T cells | 2.19E-01 | 8.20E-02 | 9.96E-01 | 9.17E-01 | Mild |
| Memory CD4 T cells | 3.96E-02 | 2.82E-01 | 9.61E-01 | 9.72E-01 | Mild |
| CD34+Progenitors | 8.41E-02 | 6.38E-01 | 9.15E-01 | 4.78E-01 | Mild |
| Dendritic cells | 5.17E-01 | 4.82E-01 | 2.90E-02 | 1.00E+00 | Mild |
| CD14+monocytes | 2.21E-01 | 7.21E-01 | 9.57E-01 | 9.97E-01 | Mild |
| Mature B cells | 6.72E-01 | 5.58E-03 | 4.70E-02 | 1 | Mild |
| Memory CD8+T cells | 3.42E-01 | 1.46E-01 | 1.84E-01 | 1 | Mild |
| Naïve CD8+T cells | 4.88E-01 | 8.71E-02 | 1.71E-01 | 6.56E-01 | Normal |
| Megakaryocytes | 4.45E-01 | 1.07E-02 | 4.79E-01 | 8.10E-01 | Normal |
| CD16+monocytes | 2.45E-01 | 7.90E-02 | 1.30E-01 | 5.04E-03 | Normal |
| Naïve CD4+T cells | 2.54E-01 | 3.75E-01 | 8.06E-01 | 4.91E-22 | Normal |
| Naïve B cells | 2.68E-01 | 3.99E-01 | 9.82E-01 | 4.82E-04 | Normal |
| NK | 7.64E-01 | 2.59E-02 | 7.03E-01 | 1.26E-01 | Normal |
| Effector CD8+T cells | 2.86E-01 | 1.01E-01 | 6.31E-01 | 2.41E-02 | Normal |
| Memory CD4 T cells | 3.82E-02 | 7.33E-01 | 8.75E-01 | 9.66E-01 | Normal |
| CD34+Progenitors | 7.42E-02 | 9.33E-01 | 3.16E-01 | 5.46E-01 | Normal |
| Dendritic cells | 6.29E-01 | 4.74E-02 | 2.00E-02 | 1 | Normal |
| CD14+monocytes | 3.67E-01 | 8.52E-01 | 8.13E-01 | 7.55E-01 | Normal |
| Mature B cells | 6.72E-01 | 1.39E-01 | 3.28E-01 | 1 | Normal |
| Memory CD8+T cells | 2.54E-01 | 2.78E-02 | 2.85E-01 | 1 | Normal |

Note: For the benchmark analyses, four tools were used to integrate a large-scale meta-GWAS summary statistics on severe COVID-19 round 4 (N = 969,689 samples, Supplementary Table S1) with scRNA-seq dataset on PBMCs (N = 514,400 cells, Supplementary Table S2) with COVID-19 of varying clinical severity, including severe, moderate, mild, and healthy controls. The GWAS summary dataset were downloaded from COVID-19 Host Genetics Consortium (https://www.covid19hg.org/; analyzed file named: "COVID19_HGI_B2_ALL_leave_23andme_20201020.txt.gz"; released date of October 4 2020). There were 7,885 hospitalized COVID-19 patients and 961,804 control participants from 21 independent contributing studies. To compare the significant levels of these methods in Figure 5A-D, we used the log transformation for the P values that were adjusted by using the Bonferroni correction method.

**Supplementary Table S5. The effector molecules for calculating molecular signature scores of naïve CD8+T cells**

| Marker genes | Full name | Gene Cards Summary |
|---|---|---|
| *PRDM1* | PR/SET Domain 1 | It is a Protein Coding gene. Diseases associated with PRDM1 include B-Cell Lymphoma and Lymphoma. Among its related pathways are Nucleotide-binding oligomerization domain (NOD) pathway and Signaling by Receptor Tyrosine Kinases. |
| *PRF1* | Perforin 1 | It is a Protein Coding gene. Diseases associated with PRF1 include Hemophagocytic Lymphohistiocytosis, Familial, 2 and Hemophagocytic Lymphohistiocytosis, Familial, 1. Among its related pathways are IL2 signaling events mediated by STAT5 and TNFR1 Pathway. |
| *GZMB* | Granzyme B | It is a Protein Coding gene. Diseases associated with GZMB include Peripheral T-Cell Lymphoma and Aggressive Nk-Cell Leukemia. Among its related pathways are TNFR1 Pathway and Downstream signaling in naïve CD8+ T cells. |
| *GNLY* | Granulysin | It is a Protein Coding gene. Diseases associated with GNLY include Erythema Multiforme and Kyphoscoliotic Heart Disease. Among its related pathways are Innate Immune System and Allograft rejection. |
| *GZMA* | Granzyme A | It is a Protein Coding gene. Diseases associated with GZMA include Chediak-Higashi Syndrome and Smallpox. Among its related pathways are NF-kappaB Signaling and IL-9 Signaling Pathways. |

| | | |
|---|---|---|
| *IFNG* | Interferon Gamma | It is a Protein Coding gene. Diseases associated with IFNG include Immunodeficiency 69 and Hepatitis C Virus. Among its related pathways are Dendritic Cells Developmental Lineage Pathway and Toll Comparative Pathway. |
| *FASLG* | Fas Ligand | It is a Protein Coding gene. Diseases associated with FASLG include Autoimmune Lymphoproliferative Syndrome and Lung Cancer. Among its related pathways are TNF Superfamily - Human Ligand-Receptor Interactions and their Associated Functions and TNFR1 Pathway. |

**Supplementary Table S6. Benchmarking analyses of scPagwas with LDSC, MAGMA, and RolyPoly for AD-associated brain cell types**

| Cell types | LDSC-SEG P value | MAGMA P value | RolyPoly P value | scPagwas P value |
|---|---|---|---|---|
| OPC | 0.96 | 0.98 | 0.49 | 4.35E-22 |
| Microglia | 0.07 | 0.034 | 0.079 | 0.015 |
| Oligodendrocyte | 0.38 | 0.80 | 0.99 | 0.42 |
| Neuron | 0.79 | 0.13 | 0.39 | 0.76 |
| Astrocyte | 0.25 | 0.047 | 0.83 | 0.99 |

**Supplementary Table S7. Validation of the association of OPC and microglia with AD in three large and independent single-cell datasets using scPagwas**

| Dataset ID | Resources | Publication year | OPC P-value | Microglia P value | Cell counts | Reference |
|---|---|---|---|---|---|---|
| Dataset #1 | Mouse brain scRNA-seq data | 2018 | 0.01 | 0.12 | 160,796 | PMID: 30096314 |
| Dataset #2 | Human brain snRNA-seq data | 2022 | 0.06 | 0.04 | 101,906 | PMID: 34767070 |
| Dataset #3 | Human brain snRNA-seq data | 2020 | 0.0036 | 4.18E-46 | 14,287 | PMID: 32826893 |

Note: Dataset #1 contains 5,435 microglial cells and 820 OPCs, dataset #2 contains 29,130 microglial cells and 1,622 OPCs, and dataset #3 contains 661 microglial cells and 476 OPCs. Since these three datasets were used for independent validation, thus we only chose the nominally significant level as threshold for replicating significant AD-relevant cell types of OPC and microglia.

**Supplementary Table S8. GO-term enrichment analysis using cellular component terms of top-ranked 1,000 risk genes for AD**

| Gene Set | Description | Enrichment ratio | P value | FDR |
|---|---|---|---|---|
| GO:0030139 | Endocytic vesicle | 2.39 | 3.10E-07 | 5.34E-05 |
| GO:0042611 | MHC protein complex | 7.59 | 8.72E-07 | 6.26E-05 |

| | | | | |
|---|---|---|---|---|
| GO:0030055 | Cell-substrate junction | 2.07 | 1.09E-06 | 6.26E-05 |
| GO:0099572 | Postsynaptic specialization | 2.08 | 7.41E-06 | 2.69E-04 |
| GO:0098984 | Neuron to neuron synapse | 2.08 | 7.95E-06 | 2.69E-04 |
| GO:0031252 | Cell leading edge | 1.98 | 9.38E-06 | 2.69E-04 |
| GO:0043235 | Receptor complex | 1.97 | 1.14E-05 | 2.80E-04 |
| GO:0098978 | Glutamatergic synapse | 2.02 | 1.48E-05 | 3.19E-04 |
| GO:0097060 | Synaptic membrane | 1.85 | 4.09E-05 | 7.82E-04 |
| GO:0030667 | Secretory granule membrane | 1.93 | 2.47E-04 | 4.25E-03 |
| GO:0060076 | Excitatory synapse | 3.62 | 3.38E-04 | 5.28E-03 |
| GO:0098685 | Schaffer collateral - CA1 synapse | 2.81 | 6.31E-04 | 9.04E-03 |
| GO:0005911 | Cell-cell junction | 1.65 | 1.07E-03 | 1.20E-02 |
| GO:1990351 | Transporter complex | 1.76 | 1.07E-03 | 1.20E-02 |
| GO:0098793 | Presynapse | 1.61 | 1.07E-03 | 1.20E-02 |
| GO:0005581 | Collagen trimer | 2.65 | 1.12E-03 | 1.20E-02 |
| GO:0044309 | Neuron spine | 2.02 | 2.73E-03 | 2.75E-02 |
| GO:0098552 | Side of membrane | 1.55 | 3.06E-03 | 2.75E-02 |
| GO:0005798 | Golgi-associated vesicle | 1.99 | 3.13E-03 | 2.75E-02 |
| GO:0005667 | Transcription factor complex | 1.65 | 3.20E-03 | 2.75E-02 |
| GO:0098589 | Membrane region | 1.64 | 5.10E-03 | 4.18E-02 |
| GO:0097038 | Perinuclear endoplasmic reticulum | 4.22 | 5.39E-03 | 4.21E-02 |
| GO:0098982 | GABA-ergic synapse | 2.36 | 9.16E-03 | 6.85E-02 |
| GO:0001772 | Immunological synapse | 3.13 | 1.10E-02 | 7.85E-02 |
| GO:0030133 | Transport vesicle | 1.52 | 1.21E-02 | 8.33E-02 |
| GO:0043198 | Dendritic shaft | 3.04 | 1.26E-02 | 8.34E-02 |
| GO:0015629 | Actin cytoskeleton | 1.43 | 1.43E-02 | 9.13E-02 |
| GO:0005775 | Vacuolar lumen | 1.77 | 1.55E-02 | 9.53E-02 |

**Supplementary Table S9. Differential gene expression analysis of top-ranked trait-relevant genes in OPC cells among two independent bulk-based gene expression datasets**

| | **GSE15222 (n = 363)** | | | **GSE109887 (n =78 )** | | |
|---|---|---|---|---|---|---|
| **Gene** | **P value** | **Mean Control (n = 187)** | **Mean AD (n = 176)** | **P value** | **Mean Control (n =46)** | **Mean AD (n = 46)** |
| *MCM7* | 1.89E-21 | 8.69 | 9.44 | 1.20E-08 | 7.59 | 8.14 |
| *CREB1* | 1.48E-18 | 10.09 | 10.83 | 5.82E-05 | 11.20 | 11.47 |
| *GFAP* | 5.92E-16 | 13.51 | 14.24 | 9.02E-06 | 11.60 | 12.53 |
| *TBC1D2B* | 4.54E-17 | 7.21 | 7.63 | 1.02E-07 | 7.96 | 8.33 |
| *LRRFIP1* | 2.64E-17 | 9.97 | 10.80 | 3.02E-02 | 10.71 | 10.98 |
| *CAPN2* | 2.18E-14 | 10.47 | 10.81 | NA | NA | NA |
| *COLGALT1* | 5.76E-15 | 6.26 | 6.70 | NA | NA | NA |
| *PHKB* | 8.76E-14 | 7.07 | 7.42 | 1.99E-01 | 7.72 | 7.66 |
| *BHLHE41* | 5.08E-14 | 7.40 | 7.78 | NA | NA | NA |
| *SEMA3E* | 8.54E-15 | 10.25 | 10.86 | 3.99E-02 | 6.60 | 6.54 |
| *AXIN1* | 1.46E-12 | 7.19 | 7.61 | 6.87E-01 | 6.53 | 6.52 |

| | | | | | | |
|---|---|---|---|---|---|---|
| *SLCO4A1* | 2.33E-14 | 7.30 | 8.25 | 1.79E-04 | 7.52 | 7.98 |
| *ATOH8* | 1.66E-12 | 8.51 | 9.20 | NA | NA | NA |
| *BAX* | 5.89E-07 | 3.38 | 4.06 | 1.74E-01 | 6.99 | 7.03 |
| *SYNM* | 4.58E-09 | 9.94 | 10.27 | 6.61E-01 | 6.56 | 6.57 |
| *BBX* | 1.02E-10 | 7.33 | 7.92 | 1.39E-06 | 9.10 | 9.49 |
| *DYNC1LI2* | 7.54E-12 | 8.39 | 8.95 | NA | NA | NA |
| *ST5* | 6.33E-12 | 7.36 | 7.74 | 1.43E-02 | 7.03 | 7.11 |
| *MYH9* | 2.47E-11 | 9.10 | 9.41 | 1.76E-04 | 10.47 | 10.78 |
| *ZMAT3* | 2.34E-11 | 11.65 | 12.08 | 6.67E-01 | 12.05 | 12.08 |
| *SLC2A3* | 1.81E-13 | 9.32 | 9.84 | 6.49E-01 | 11.02 | 10.97 |
| *KAZN* | 7.99E-12 | 5.68 | 6.26 | NA | NA | NA |
| *SYK* | 1.28E-06 | 5.15 | 5.75 | 2.88E-06 | 6.93 | 7.23 |
| *HM13* | 1.28E-07 | 2.72 | 3.80 | 1.41E-01 | 6.69 | 6.71 |
| *FOXN3* | 2.30E-15 | 9.37 | 9.85 | 5.95E-01 | 6.57 | 6.58 |
| *PLEKHM1* | 4.98E-12 | 6.79 | 7.10 | NA | NA | NA |
| *RPS3* | 1.63E-08 | 9.63 | 9.99 | 1.12E-04 | 10.59 | 10.87 |
| *LYN* | 6.34E-12 | 6.23 | 6.76 | NA | NA | NA |
| *PLXDC2* | 2.31E-08 | 6.25 | 6.82 | 2.02E-07 | 8.06 | 8.60 |
| *ANGPT2* | 1.33E-12 | 4.53 | 5.65 | 1.92E-05 | 7.05 | 7.59 |
| *CTBP2* | 4.71E-11 | 8.31 | 8.73 | 2.45E-03 | 6.83 | 6.90 |
| *UPF2* | 2.85E-09 | 6.49 | 7.10 | 9.53E-03 | 9.68 | 9.78 |
| *CNOT1* | 3.88E-09 | 6.74 | 7.01 | 2.25E-01 | 8.81 | 8.87 |
| *INPP5D* | 4.81E-08 | 5.96 | 6.39 | 3.36E-08 | 7.35 | 7.91 |
| *CD151* | 2.49E-08 | 7.87 | 8.24 | 3.08E-07 | 9.32 | 9.94 |
| *NCOA3* | 3.62E-08 | 6.71 | 6.97 | 5.38E-02 | 6.60 | 6.63 |
| *GSK3B* | 5.30E-07 | 6.13 | 6.45 | 4.30E-02 | 8.18 | 8.29 |
| *ETV6* | 4.56E-06 | 6.11 | 6.58 | 1.37E-04 | 8.04 | 8.33 |
| *DBF4* | 3.19E-12 | 11.45 | 12.15 | 4.34E-01 | 6.64 | 6.63 |
| *LRFN4* | 1.44E-09 | 7.77 | 8.14 | 2.05E-01 | 7.67 | 7.74 |
| *CXCR4* | 1.29E-10 | 6.16 | 6.87 | 2.01E-03 | 6.60 | 6.69 |
| *COASY* | 6.76E-08 | 9.41 | 9.58 | 3.65E-04 | 9.16 | 9.43 |
| *MAP3K1* | 1.85E-09 | 7.43 | 7.88 | NA | NA | NA |
| *ITGA8* | 5.22E-07 | 4.70 | 5.34 | 1.81E-04 | 6.95 | 7.24 |
| *CREBBP* | 1.56E-08 | 7.57 | 7.80 | 8.22E-04 | 8.40 | 8.56 |
| *GLRX3* | 2.21E-08 | 2.68 | 3.70 | 2.82E-02 | 7.97 | 7.72 |
| *ZFHX3* | 6.67E-05 | 6.48 | 6.81 | NA | NA | NA |
| *TPD52L2* | 3.42E-09 | 9.16 | 9.42 | 5.44E-03 | 9.88 | 10.10 |
| *TNS3* | 4.35E-09 | 9.83 | 10.26 | 3.88E-06 | 10.88 | 11.50 |
| *CEBPD* | 3.87E-08 | 9.10 | 9.62 | 2.34E-05 | 10.30 | 10.93 |
| *POU2F2* | 4.14E-10 | 5.51 | 6.03 | 9.41E-01 | 6.44 | 6.45 |
| *UNC13D* | 4.08E-08 | 4.59 | 5.21 | 3.92E-01 | 6.57 | 6.59 |
| *LHFPL2* | 1.03E-07 | 7.21 | 7.55 | NA | NA | NA |
| *RBM42* | 5.10E-08 | 8.89 | 9.03 | 8.36E-02 | 8.68 | 8.52 |
| *ATP11C* | 3.79E-08 | 4.95 | 5.39 | 8.33E-07 | 6.91 | 7.08 |
| *ARPC1B* | 1.17E-07 | 6.78 | 7.26 | 3.19E-01 | 6.72 | 6.73 |
| *RXRA* | 9.25E-07 | 10.03 | 10.33 | 3.91E-06 | 10.21 | 10.77 |

| | | | | | | |
|---|---|---|---|---|---|---|
| *HLA-DPB1* | 3.41E-09 | 10.11 | 10.51 | 7.23E-01 | 6.88 | 6.90 |
| *ID3* | 8.55E-05 | 7.36 | 7.87 | 3.92E-04 | 8.05 | 8.60 |
| *AK2* | 1.05E-07 | 8.26 | 8.49 | 3.84E-04 | 8.08 | 8.28 |
| *LAPTM5* | 2.05E-07 | 6.38 | 6.78 | 8.10E-06 | 7.29 | 7.79 |
| *ZRANB1* | 7.20E-08 | 7.52 | 8.14 | 2.78E-02 | 8.80 | 9.09 |
| *GRN* | 3.63E-07 | 7.10 | 7.32 | 1.34E-02 | 7.14 | 7.25 |
| *KLF6* | 4.93E-07 | 5.37 | 5.80 | 1.70E-06 | 8.30 | 8.70 |
| *EBF4* | 3.45E-03 | 6.34 | 6.58 | 7.43E-02 | 6.77 | 6.83 |
| *ZFP36L1* | 7.31E-06 | 11.11 | 11.44 | 3.01E-06 | 9.61 | 10.32 |
| *TSHZ1* | 4.21E-08 | 7.33 | 7.65 | 7.92E-01 | 8.48 | 8.46 |
| *RHBDF2* | 3.60E-07 | 5.53 | 5.98 | 3.54E-06 | 7.64 | 8.14 |
| *CSF2RA* | 1.96E-05 | 4.84 | 5.16 | 1.13E-06 | 10.46 | 11.05 |
| *PLCG1* | 2.94E-10 | 10.16 | 10.54 | 9.19E-08 | 8.42 | 8.79 |
| *BTG1* | 2.96E-11 | 9.11 | 9.63 | 1.74E-05 | 10.19 | 10.49 |
| *PABPC1* | 6.71E-06 | 9.54 | 9.85 | 4.85E-05 | 9.87 | 10.13 |
| *ETV1* | 2.38E-04 | 3.95 | 4.34 | 4.91E-03 | 6.71 | 6.61 |
| *RHBDD3* | 1.27E-07 | 6.51 | 6.81 | NA | NA | NA |
| *PELI2* | 1.59E-07 | 5.56 | 5.97 | 7.40E-04 | 7.81 | 8.03 |
| *SUGT1* | 7.78E-08 | 8.43 | 8.77 | 2.13E-01 | 8.29 | 8.38 |
| *ZIC2* | 6.21E-10 | 8.67 | 9.42 | 3.17E-03 | 9.33 | 9.73 |
| *BEST3* | 2.67E-08 | 4.68 | 5.28 | NA | NA | NA |
| *TRAFD1* | 7.76E-07 | 7.25 | 7.48 | 1.14E-02 | 7.32 | 7.41 |
| *LAT2* | 1.80E-09 | 5.86 | 6.46 | NA | NA | NA |
| *RASAL2* | 4.17E-04 | 4.53 | 4.78 | 1.78E-04 | 6.76 | 6.67 |
| *44810* | 2.19E-04 | 4.77 | 5.21 | NA | NA | NA |
| *NCOA5* | 8.02E-05 | 4.46 | 4.80 | NA | NA | NA |
| *TRIO* | 8.55E-04 | 5.11 | 5.62 | 4.89E-02 | 7.92 | 7.83 |
| *RPS6KB2* | 1.64E-06 | 6.96 | 7.24 | 9.59E-04 | 8.41 | 8.79 |
| *SCN1A* | 1.73E-05 | 6.95 | 7.42 | 6.09E-01 | 9.11 | 9.18 |
| *IKZF1* | 1.92E-03 | 3.24 | 3.74 | 3.53E-03 | 6.72 | 6.90 |
| *JDP2* | 2.03E-09 | 7.19 | 7.53 | 2.58E-04 | 7.41 | 7.71 |
| *FRMD6* | 1.16E-05 | 5.81 | 6.24 | 4.44E-02 | 7.21 | 7.32 |
| *CHD1* | 6.64E-06 | 5.64 | 5.98 | NA | NA | NA |
| *ASCL1* | 1.06E-04 | 4.64 | 5.08 | 1.79E-05 | 9.24 | 9.82 |
| *CDC25B* | 3.81E-08 | 7.92 | 8.26 | 8.51E-03 | 8.13 | 8.28 |
| *GPRIN3* | 9.07E-07 | 5.79 | 6.16 | 5.92E-02 | 6.55 | 6.58 |
| *ANKRD44* | 2.01E-08 | 12.36 | 12.84 | 6.82E-03 | 6.49 | 6.53 |
| *RAPGEF1* | 4.85E-06 | 8.77 | 8.97 | 6.96E-04 | 8.13 | 8.44 |
| *CD81* | 1.69E-05 | 12.43 | 12.64 | 7.25E-02 | 12.99 | 13.08 |
| *EBF1* | 7.41E-05 | 3.21 | 4.09 | 1.03E-01 | 7.24 | 7.37 |
| *PRPSAP1* | 7.61E-06 | 8.98 | 9.09 | 3.77E-01 | 9.94 | 9.99 |
| *CEBPG* | 3.48E-07 | 7.83 | 8.07 | NA | NA | NA |
| *CYBA* | 2.21E-05 | 6.89 | 7.30 | 3.67E-01 | 7.19 | 7.29 |
| *GPC6* | 1.04E-05 | 2.44 | 3.37 | 4.27E-01 | 7.64 | 7.71 |
| *ANO6* | 3.45E-04 | 6.20 | 6.48 | NA | NA | NA |
| *ZNHIT6* | 1.35E-05 | 8.39 | 8.57 | 1.34E-05 | 8.92 | 9.17 |

| | | | | | | |
|---|---|---|---|---|---|---|
| *CD44* | 3.47E-10 | 6.12 | 6.61 | 1.94E-03 | 8.10 | 8.68 |
| *THAP4* | 4.09E-06 | 9.92 | 10.04 | 2.68E-01 | 6.49 | 6.47 |
| *ARHGAP17* | 1.26E-05 | 7.29 | 7.48 | 1.65E-05 | 8.02 | 8.33 |
| *MYH14* | 8.01E-06 | 7.13 | 7.37 | 6.18E-01 | 6.83 | 6.81 |
| *RHOC* | 1.13E-04 | 9.53 | 9.82 | 9.30E-06 | 9.55 | 10.08 |
| *SLC25A39* | 1.41E-05 | 9.47 | 9.65 | 1.05E-05 | 10.00 | 10.40 |
| *GLB1* | 1.98E-05 | 7.18 | 7.33 | 6.10E-02 | 7.86 | 7.95 |
| *SLC7A6OS* | 8.49E-03 | 3.77 | 4.13 | 2.77E-01 | 6.88 | 6.92 |
| *RGS17* | 4.07E-05 | 8.79 | 9.04 | 6.20E-01 | 9.29 | 9.33 |
| *PAG1* | 1.87E-06 | 7.54 | 7.92 | 7.97E-03 | 8.01 | 8.15 |
| *CMTM7* | 7.34E-06 | 5.81 | 6.21 | 1.40E-01 | 6.87 | 6.95 |
| *CLK1* | 2.23E-05 | 8.92 | 9.15 | 4.96E-02 | 9.43 | 9.64 |
| *VCAN* | 1.10E-05 | 8.14 | 8.48 | 6.97E-09 | 8.87 | 9.66 |
| *IL6ST* | 8.62E-08 | 1.65 | 2.92 | 7.97E-01 | 6.90 | 6.91 |
| *C6orf89* | 6.93E-03 | 3.72 | 4.11 | 6.60E-03 | 6.91 | 6.84 |
| *IL13RA1* | 7.76E-03 | 7.38 | 7.59 | NA | NA | NA |
| *FYB* | 1.70E-04 | 3.91 | 4.29 | 3.87E-02 | 7.31 | 7.57 |
| *ZNF431* | 5.30E-06 | 3.06 | 3.70 | 1.59E-02 | 6.74 | 6.81 |
| *SCIN* | 2.24E-07 | 13.13 | 13.58 | 1.15E-04 | 6.91 | 7.35 |
| *MECP2* | 4.03E-05 | 6.90 | 7.10 | 4.24E-04 | 7.20 | 6.93 |
| *PRKCH* | 1.82E-04 | 5.35 | 5.68 | 4.70E-05 | 7.76 | 8.09 |
| *RUNX1* | 3.72E-08 | 3.96 | 4.78 | 1.96E-06 | 6.55 | 6.66 |
| *GTF3C1* | 2.39E-05 | 7.52 | 7.69 | 2.35E-01 | 8.05 | 7.96 |
| *CHIC2* | 3.58E-05 | 6.98 | 7.15 | 4.08E-01 | 8.90 | 8.96 |
| *STAB1* | 4.21E-07 | 7.00 | 7.47 | 3.48E-03 | 6.89 | 7.15 |
| *CARD11* | 2.51E-06 | 3.84 | 4.25 | 5.81E-04 | 6.94 | 7.16 |
| *ACSL1* | 1.42E-05 | 6.57 | 6.89 | 1.19E-04 | 8.58 | 9.00 |
| *MYOF* | 7.69E-04 | 5.43 | 5.69 | 1.74E-03 | 7.17 | 7.50 |
| *SH3TC1* | 1.04E-04 | 4.67 | 5.07 | 1.46E-04 | 6.94 | 7.23 |
| *PALD1* | 2.86E-04 | 7.12 | 7.36 | NA | NA | NA |
| *JUND* | 3.56E-05 | 13.08 | 13.26 | 4.36E-01 | 13.43 | 13.39 |
| *DUSP1* | 1.94E-07 | 8.14 | 8.60 | 1.97E-02 | 9.81 | 10.32 |
| *SLC6A13* | 6.15E-06 | 5.85 | 6.35 | 4.24E-02 | 6.53 | 6.57 |
| *PLOD3* | 1.20E-05 | 6.57 | 6.90 | NA | NA | NA |
| *SF3A3* | 3.65E-05 | 8.28 | 8.37 | 2.97E-02 | 9.78 | 9.95 |
| *CCND1* | 9.61E-04 | 7.19 | 7.46 | 2.70E-04 | 9.26 | 9.80 |
| *PRPF38B* | 7.89E-06 | 3.12 | 3.86 | 5.49E-02 | 7.12 | 7.24 |
| *MED13L* | 3.13E-04 | 6.32 | 6.58 | 1.08E-03 | 7.52 | 7.71 |
| *HCLS1* | 2.32E-03 | 6.99 | 7.28 | 3.41E-02 | 7.99 | 8.31 |
| *SEC24C* | 2.35E-05 | 9.12 | 9.31 | 1.31E-02 | 8.92 | 9.07 |
| *PEAK1* | 1.50E-04 | 9.43 | 9.57 | NA | NA | NA |
| *XRN2* | 3.00E-03 | 6.73 | 6.87 | 4.65E-01 | 7.08 | 7.11 |
| *IL18* | 1.11E-07 | 11.68 | 12.29 | 1.17E-05 | 13.41 | 13.61 |
| *SOCS5* | 7.13E-04 | 7.11 | 7.32 | 4.34E-02 | 7.04 | 6.96 |
| *BRAF* | 1.42E-03 | 4.68 | 4.89 | 4.73E-01 | 6.80 | 6.84 |
| *CREB3* | 6.21E-04 | 8.78 | 8.94 | NA | NA | NA |

| | | | | | | |
|---|---|---|---|---|---|---|
| *SPAG1* | 1.46E-07 | 1.86 | 2.82 | 1.32E-03 | 6.65 | 6.72 |
| *CD86* | 3.67E-03 | 4.16 | 4.49 | 6.99E-04 | 6.99 | 7.17 |
| *TBC1D1* | 5.58E-05 | 7.68 | 7.89 | NA | NA | NA |
| *TXNIP* | 3.29E-04 | 9.20 | 9.60 | 1.59E-03 | 11.06 | 11.66 |
| *TAL1* | 1.10E-03 | 5.04 | 5.31 | 2.85E-06 | 6.61 | 6.74 |
| *ATF4* | 8.83E-05 | 6.94 | 7.20 | 3.64E-05 | 10.16 | 10.66 |
| *CSF1R* | 4.84E-03 | 9.22 | 9.45 | 3.72E-06 | 9.71 | 10.38 |
| *DVL1* | 1.23E-02 | 4.12 | 4.34 | 5.84E-02 | 6.57 | 6.55 |
| *CHN2* | 2.48E-03 | 5.88 | 6.30 | 3.37E-01 | 8.43 | 8.53 |
| *RBM47* | 1.94E-04 | 3.17 | 3.77 | 1.89E-03 | 6.90 | 7.21 |
| *GGA2* | 3.62E-04 | 4.76 | 4.98 | 5.37E-06 | 8.40 | 8.77 |
| *FBXW7* | 3.41E-04 | 6.04 | 6.44 | 3.60E-06 | 9.77 | 9.37 |
| *TANC1* | 5.69E-03 | 5.16 | 5.40 | 8.22E-02 | 7.87 | 8.00 |
| *RPIA* | 9.54E-04 | 6.09 | 6.26 | 1.76E-01 | 7.90 | 7.97 |
| *SLCO2B1* | 4.82E-03 | 8.50 | 8.70 | 5.82E-05 | 8.92 | 9.44 |
| *SOAT1* | 1.90E-04 | 4.50 | 4.79 | 3.72E-02 | 6.77 | 6.83 |
| *RAB3IP* | 5.96E-06 | 7.81 | 8.21 | 1.87E-01 | 8.37 | 8.46 |
| *PARP14* | 7.00E-05 | 6.38 | 6.65 | 1.24E-04 | 7.34 | 7.63 |
| *IRAK3* | 1.66E-04 | 4.34 | 4.76 | 2.65E-03 | 6.95 | 7.12 |
| *ADAM15* | 2.77E-04 | 6.90 | 7.11 | 1.52E-01 | 7.10 | 7.17 |
| *MSR1* | 5.84E-05 | 1.67 | 2.42 | 1.58E-03 | 6.59 | 6.65 |
| *ALOX5* | 2.89E-05 | 6.88 | 7.30 | 7.98E-01 | 6.51 | 6.51 |
| *NAA16* | 5.40E-04 | 2.46 | 3.13 | NA | NA | NA |
| *KCNMB1* | 1.04E-02 | 4.79 | 5.02 | 8.45E-01 | 6.99 | 7.01 |
| *KLHL5* | 2.37E-03 | 8.50 | 8.68 | 2.16E-03 | 9.12 | 9.40 |
| *CPVL* | 2.39E-04 | 5.12 | 5.57 | 1.10E-01 | 7.59 | 7.88 |
| *STK32A* | 1.66E-03 | 3.15 | 3.53 | 1.95E-01 | 6.46 | 6.48 |
| *CACNA1A* | 1.51E-04 | 6.64 | 7.03 | 9.38E-03 | 7.21 | 7.02 |
| *PTGFR* | 1.18E-03 | 2.05 | 2.63 | 1.41E-02 | 6.63 | 6.67 |
| *PTEN* | 2.56E-02 | 5.97 | 6.11 | NA | NA | NA |
| *SRGAP2* | 2.49E-05 | 7.99 | 8.38 | 8.12E-01 | 6.85 | 6.85 |
| *SUSD6* | 7.66E-03 | 5.58 | 5.82 | NA | NA | NA |
| *IL16* | 1.43E-06 | 4.14 | 4.92 | 4.29E-01 | 6.55 | 6.56 |
| *ID1* | 2.97E-02 | 6.37 | 6.62 | 1.60E-03 | 7.62 | 7.97 |
| *NCKAP1L* | 1.89E-04 | 5.00 | 5.40 | 1.71E-04 | 7.19 | 7.55 |
| *MS4A6A* | 2.49E-05 | 5.86 | 6.26 | 8.35E-03 | 7.60 | 8.03 |
| *AKIP1* | 6.30E-04 | 5.89 | 6.12 | NA | NA | NA |
| *ARHGAP22* | 9.38E-04 | 6.97 | 7.17 | 7.93E-05 | 8.00 | 8.34 |
| *MTSS1* | 8.00E-03 | 5.93 | 6.11 | 3.66E-02 | 9.47 | 9.57 |
| *TIAM1* | 3.47E-03 | 2.53 | 3.02 | 8.95E-01 | 7.49 | 7.48 |
| *TRPC3* | 3.07E-02 | 4.61 | 4.78 | 8.05E-01 | 6.83 | 6.84 |
| *CRKL* | 3.58E-04 | 9.28 | 9.39 | 9.64E-03 | 10.84 | 10.98 |
| *RBMS1* | 4.36E-03 | 6.38 | 6.54 | 6.76E-02 | 6.86 | 6.92 |
| *PILRA* | 6.31E-03 | 3.48 | 3.78 | 6.70E-02 | 6.91 | 6.96 |
| *ADAM28* | 1.71E-02 | 3.78 | 4.02 | 1.00E-03 | 6.53 | 6.58 |
| *OLIG1* | 7.10E-03 | 9.71 | 9.94 | 4.45E-05 | 9.56 | 10.13 |

| | | | | | | |
|---|---|---|---|---|---|---|
| *ASIC4* | 5.76E-04 | 5.98 | 6.21 | NA | NA | NA |
| *SLU7* | 4.02E-04 | 7.21 | 7.35 | 9.16E-01 | 7.90 | 7.89 |
| *LRBA* | 9.32E-04 | 6.39 | 6.53 | 6.75E-02 | 7.46 | 7.52 |
| *C3* | 1.10E-03 | 8.88 | 9.22 | 7.99E-01 | 6.73 | 6.71 |
| *EML4* | 2.45E-03 | 6.86 | 6.96 | 6.41E-03 | 8.05 | 8.17 |
| *GOLGA8A* | 4.79E-07 | 10.03 | 9.58 | 7.18E-01 | 8.73 | 8.77 |
| *BLNK* | 9.84E-03 | 2.38 | 2.87 | 2.53E-01 | 6.49 | 6.50 |
| *HGS* | 4.06E-04 | 10.07 | 10.18 | 2.30E-01 | 10.86 | 10.94 |
| *HSPA1A* | 5.72E-06 | 8.83 | 9.42 | 7.97E-04 | 10.48 | 11.24 |
| *ABHD12* | 3.52E-03 | 7.06 | 7.22 | 9.96E-02 | 7.87 | 7.78 |
| *AKAP13* | 1.82E-02 | 3.79 | 4.08 | 1.13E-04 | 7.98 | 8.12 |
| *RASSF2* | 2.44E-02 | 2.06 | 2.50 | 1.71E-04 | 8.94 | 9.39 |
| *TSPAN9* | 5.57E-06 | 8.33 | 8.83 | 3.05E-02 | 9.78 | 9.97 |
| *SLC25A37* | 1.91E-03 | 7.53 | 7.75 | 1.68E-02 | 7.79 | 7.98 |
| *PPARD* | 3.99E-02 | 3.20 | 3.47 | 2.95E-01 | 7.01 | 7.04 |
| *AFAP1L2* | 1.33E-02 | 7.98 | 8.12 | 2.35E-01 | 8.68 | 8.78 |
| *GSDMB* | 2.04E-04 | 7.54 | 7.78 | 7.76E-05 | 8.32 | 8.74 |
| *CRTAM* | 3.05E-03 | 2.62 | 5.18 | 8.02E-01 | 6.49 | 6.49 |
| *COL9A1* | 2.99E-03 | 7.51 | 7.75 | 3.06E-03 | 7.03 | 7.24 |
| *SMOC1* | 4.46E-03 | 7.68 | 7.91 | 1.12E-02 | 7.25 | 7.48 |
| *VSIG4* | 3.53E-03 | 5.52 | 5.94 | 3.36E-02 | 7.12 | 7.34 |
| *ARL5A* | 2.73E-02 | 4.64 | 4.82 | 6.32E-02 | 10.12 | 10.24 |
| *PLCB4* | 4.89E-02 | 7.38 | 7.59 | 2.65E-01 | 6.79 | 6.82 |
| *RAB9B* | 2.48E-02 | 6.54 | 6.81 | 3.65E-03 | 8.46 | 8.20 |
| *PIK3R5* | 6.95E-03 | 3.03 | 3.42 | 7.80E-02 | 6.54 | 6.57 |
| *FAM110B* | 5.41E-03 | 7.80 | 7.95 | 4.41E-01 | 8.36 | 8.41 |
| *C1QA* | 1.64E-03 | 8.40 | 8.74 | 1.62E-01 | 6.93 | 7.01 |
| *HSPA1B* | 1.10E-03 | 10.12 | 10.47 | 3.23E-01 | 11.31 | 11.54 |
| *HLA-DMB* | 1.20E-02 | 8.53 | 8.77 | 1.18E-06 | 8.67 | 9.37 |
| *CALCRL* | 3.58E-04 | 3.57 | 3.99 | 2.53E-02 | 6.92 | 7.05 |
| *HLA-DPA1* | 4.93E-03 | 9.16 | 9.41 | 2.94E-01 | 8.32 | 8.53 |
| *CD74* | 1.11E-02 | 9.41 | 9.65 | 6.90E-01 | 8.75 | 8.84 |
| *TYROBP* | 1.31E-02 | 7.23 | 7.49 | 5.54E-02 | 8.03 | 8.35 |
| *SLC22A3* | 1.01E-02 | 3.35 | 3.67 | 5.87E-02 | 6.61 | 6.66 |
| *CTNNB1* | 8.58E-03 | 4.81 | 5.02 | 1.03E-01 | 6.86 | 6.81 |
| *CDKN1A* | 4.61E-04 | 8.78 | 9.18 | 3.29E-04 | 8.63 | 9.21 |
| *PTPRE* | 1.98E-02 | 1.58 | 2.21 | 1.31E-02 | 8.32 | 8.16 |
| *CLCN6* | 1.30E-02 | 3.49 | 3.74 | 3.16E-01 | 8.85 | 8.81 |
| *DOCK2* | 1.68E-02 | 5.52 | 5.73 | 1.53E-03 | 7.62 | 7.93 |
| *MFSD5* | 4.37E-03 | 6.74 | 6.85 | 7.37E-03 | 7.52 | 7.61 |
| *RSAD2* | 9.51E-10 | 8.43 | 9.11 | 6.61E-03 | 7.01 | 6.84 |
| *HIRA* | 4.88E-03 | 4.18 | 4.51 | 8.02E-01 | 7.10 | 7.09 |
| *TNK2* | 5.50E-03 | 10.73 | 10.85 | 2.74E-03 | 9.73 | 9.98 |
| *SH3GL1* | 6.60E-03 | 5.73 | 5.86 | 1.55E-02 | 8.19 | 8.41 |
| *C1QB* | 4.85E-03 | 9.92 | 10.24 | NA | NA | NA |
| *GNB4* | 1.11E-11 | 6.49 | 7.35 | 2.52E-04 | 7.01 | 7.18 |

| | | | | | | |
|---|---|---|---|---|---|---|
| *GRIA4* | 1.39E-03 | 4.63 | 5.02 | 5.74E-02 | 7.53 | 7.37 |
| *KCTD16* | 3.84E-02 | 3.89 | 4.16 | 5.67E-04 | 7.44 | 7.15 |
| *NELFB* | 2.09E-03 | 8.51 | 8.64 | NA | NA | NA |
| *SLC7A5* | 8.13E-04 | 7.41 | 7.66 | 7.15E-04 | 9.38 | 9.85 |
| *PDZD2* | 5.82E-03 | 1.85 | 2.42 | 3.65E-01 | 9.63 | 9.53 |
| *FERMT1* | 9.28E-03 | 7.07 | 7.30 | 5.14E-01 | 6.81 | 6.84 |
| *IGFBP5* | 7.60E-04 | 2.20 | 2.93 | 4.88E-05 | 9.70 | 10.14 |
| *RALGPS2* | 1.48E-04 | 2.23 | 3.04 | 3.50E-01 | 6.76 | 6.80 |
| *HNMT* | 4.12E-02 | 6.01 | 6.15 | 4.49E-02 | 7.55 | 7.63 |
| *OPCML* | 1.10E-02 | 7.82 | 7.98 | 6.44E-05 | 9.78 | 9.26 |
| *UNC80* | 3.25E-02 | 3.05 | 3.35 | NA | NA | NA |
| *ITGAL* | 2.52E-02 | 3.53 | 3.79 | 6.62E-03 | 6.64 | 6.75 |
| *CCDC26* | 2.19E-04 | 2.87 | 3.42 | 9.28E-01 | 6.56 | 6.56 |
| *COL7A1* | 9.11E-03 | 7.40 | 7.61 | 9.83E-02 | 8.08 | 8.22 |
| *MYO18A* | 1.49E-04 | 1.66 | 2.39 | 1.80E-01 | 6.74 | 6.84 |
| *KYNU* | 1.22E-02 | 3.42 | 3.71 | 1.94E-01 | 6.72 | 6.77 |
| *EXOSC10* | 9.68E-03 | 9.18 | 9.28 | 2.97E-03 | 8.76 | 8.91 |
| *C4orf19* | 6.66E-03 | 4.52 | 4.82 | 2.02E-03 | 6.85 | 6.98 |
| *RAP1A* | 1.55E-03 | 6.88 | 7.08 | 2.29E-02 | 6.86 | 6.94 |
| *MFSD14A* | 4.34E-02 | 6.99 | 7.11 | NA | NA | NA |
| *FAM129A* | 9.36E-04 | 2.47 | 2.98 | 3.36E-02 | 7.37 | 7.54 |
| *MAF* | 1.00E-02 | 5.67 | 5.82 | 2.38E-03 | 7.20 | 7.33 |
| *HS6ST3* | 2.73E-02 | 2.50 | 2.87 | 3.41E-01 | 6.48 | 6.47 |
| *VCL* | 3.74E-02 | 6.17 | 6.29 | 7.53E-06 | 8.82 | 9.13 |
| *SCN2A* | 8.56E-03 | 11.53 | 11.70 | 2.22E-02 | 6.46 | 6.43 |
| *WBP1L* | 2.77E-02 | 3.14 | 3.49 | NA | NA | NA |
| *RPS3A* | 7.76E-03 | 12.56 | 12.69 | 8.00E-02 | 12.05 | 12.15 |
| *C1QC* | 6.95E-03 | 6.70 | 7.04 | 4.53E-01 | 8.48 | 8.63 |
| *C10orf10* | 4.61E-06 | 7.74 | 8.28 | 8.61E-04 | 8.44 | 9.14 |
| *CTSS* | 1.99E-02 | 5.47 | 5.69 | 7.89E-02 | 6.60 | 6.65 |
| *RPL13* | 1.49E-02 | 10.45 | 10.58 | 2.26E-03 | 8.70 | 8.92 |
| *THOC1* | 3.08E-02 | 8.74 | 8.81 | 2.51E-02 | 8.83 | 8.95 |
| *IER2* | 5.68E-03 | 6.20 | 6.36 | 1.66E-04 | 6.89 | 7.00 |
| *NTNG1* | 1.07E-08 | 7.31 | 6.86 | 1.29E-04 | 8.32 | 7.87 |
| *JUNB* | 7.64E-03 | 2.54 | 2.97 | 4.98E-01 | 6.79 | 6.76 |
| *ARPP21* | 2.93E-17 | 8.16 | 7.22 | NA | NA | NA |
| *ZNF385D* | 2.18E-02 | 6.42 | 6.64 | 7.51E-02 | 8.42 | 8.26 |
| *HLA-DQA1* | 1.88E-02 | 4.82 | 5.31 | NA | NA | NA |
| *SP100* | 1.88E-02 | 11.04 | 11.10 | 3.03E-04 | 6.71 | 6.83 |
| *PTPRC* | 2.90E-04 | 1.64 | 2.42 | 2.74E-04 | 6.49 | 6.51 |
| *TLR2* | 3.77E-02 | 5.71 | 6.00 | 1.25E-02 | 6.47 | 6.52 |
| *PKIB* | 2.11E-01 | 0.70 | 1.15 | 1.32E-01 | 6.77 | 6.71 |
| *FCGR1A* | 2.48E-02 | 2.55 | 3.07 | 6.01E-02 | 7.12 | 7.32 |
| *PSMA1* | 1.99E-09 | 8.68 | 8.38 | 7.49E-04 | 8.53 | 8.26 |
| *PELI1* | 4.44E-02 | 6.35 | 6.46 | 8.50E-01 | 8.00 | 8.02 |
| *CEMIP* | 1.82E-02 | 7.06 | 7.33 | NA | NA | NA |

| | | | | | | |
|---|---|---|---|---|---|---|
| *MRPL27* | 8.38E-03 | 8.85 | 8.96 | 1.99E-01 | 7.73 | 7.78 |
| *LIMD1* | 3.39E-02 | 2.65 | 2.98 | 5.94E-01 | 6.48 | 6.48 |
| *GDAP2* | 3.43E-02 | 2.30 | 2.67 | 3.08E-04 | 6.68 | 6.77 |
| *ACER2* | 3.01E-02 | 3.69 | 3.95 | 8.95E-01 | 6.69 | 6.69 |
| *CUL9* | 1.67E-02 | 8.08 | 8.19 | 6.53E-01 | 8.16 | 8.19 |
| *NEU4* | 3.02E-02 | 6.44 | 6.60 | 8.01E-02 | 8.02 | 8.27 |
| *SLC7A1* | 1.79E-02 | 3.36 | 3.66 | 1.99E-01 | 9.85 | 9.96 |
| *HLA-B* | 2.97E-02 | 8.75 | 8.92 | 1.79E-01 | 8.92 | 9.12 |
| *MAP3K8* | 3.88E-02 | 4.69 | 4.93 | 4.46E-03 | 7.14 | 7.38 |
| *UBE3D* | 4.44E-02 | 4.95 | 5.05 | NA | NA | NA |
| *ZNF445* | 4.84E-03 | 2.98 | 3.37 | 8.65E-03 | 6.48 | 6.54 |
| *ZFPM2* | 6.13E-03 | 3.46 | 4.12 | 2.27E-03 | 7.33 | 7.10 |
| *ITPKC* | 9.04E-03 | 2.20 | 2.73 | 1.12E-03 | 6.74 | 6.84 |
| *RXRB* | 1.81E-02 | 7.71 | 7.78 | 6.55E-02 | 10.03 | 10.15 |
| *SORL1* | 4.24E-02 | 8.70 | 8.78 | 1.98E-02 | 9.55 | 9.38 |
| *NFYC* | 2.87E-02 | 9.03 | 9.11 | 1.04E-01 | 8.40 | 8.30 |
| *CAMKK2* | 4.80E-02 | 8.50 | 8.72 | 4.91E-03 | 9.15 | 8.77 |
| *ZFP36* | 1.56E-02 | 8.55 | 8.84 | 2.56E-03 | 8.26 | 8.81 |
| *CCL2* | 1.37E-02 | 4.73 | 5.32 | 7.67E-03 | 7.34 | 7.83 |
| *PHLDB2* | 3.79E-02 | 2.76 | 3.22 | 1.65E-01 | 6.86 | 6.94 |
| *NTN1* | 8.47E-03 | 3.10 | 3.53 | 9.91E-01 | 6.47 | 6.47 |
| *IREB2* | 2.08E-02 | 3.19 | 3.52 | 7.87E-02 | 7.15 | 7.03 |
| *CD14* | 1.26E-02 | 5.79 | 6.10 | 2.82E-04 | 9.60 | 10.23 |
| *IFI16* | 1.04E-02 | 6.67 | 6.89 | 3.78E-02 | 7.87 | 8.12 |
| *TBXAS1* | 3.60E-01 | 4.72 | 4.83 | 1.59E-02 | 7.30 | 7.52 |
| *RRP1* | 1.46E-04 | 2.23 | 2.85 | 2.29E-01 | 7.17 | 7.21 |
| *AIF1* | 4.92E-01 | 7.31 | 7.37 | 5.35E-01 | 7.58 | 7.64 |
| *SNX22* | 3.79E-02 | 1.21 | 1.75 | 2.53E-03 | 7.87 | 8.19 |
| *PGM5* | 3.15E-02 | 1.30 | 2.16 | 2.63E-04 | 6.97 | 7.20 |
| *SYN1* | 7.60E-01 | 1.80 | 1.89 | 5.25E-05 | 10.77 | 10.29 |
| *HPS3* | 2.20E-02 | 4.91 | 5.19 | 8.96E-02 | 6.97 | 7.07 |
| *FGFR1* | 1.57E-01 | 0.99 | 1.48 | 5.16E-01 | 6.49 | 6.50 |
| *EPS8* | 1.08E-02 | 5.51 | 5.80 | 1.23E-04 | 7.65 | 8.03 |
| *PYGL* | 4.00E-02 | 6.66 | 6.81 | 1.72E-03 | 8.11 | 8.44 |
| *TXLNA* | 3.19E-02 | 1.17 | 1.97 | 3.91E-05 | 9.71 | 10.15 |
| *MEGF11* | 1.42E-02 | 2.27 | 2.72 | 1.40E-01 | 6.93 | 7.01 |
| *DPP6* | 3.05E-04 | 11.72 | 11.58 | 1.24E-07 | 7.86 | 7.43 |
| *TRIM14* | 1.50E-02 | 2.42 | 2.83 | 3.05E-01 | 6.57 | 6.55 |
| *IL1RAP* | 2.69E-03 | 4.37 | 4.07 | 3.52E-01 | 6.66 | 6.68 |
| *TFEC* | 3.32E-02 | 1.92 | 2.38 | 6.96E-01 | 6.56 | 6.57 |
| *MAML3* | 1.06E-02 | 2.05 | 2.61 | 1.05E-04 | 6.52 | 6.58 |
| *CD69* | 4.28E-02 | 1.52 | 2.01 | 9.62E-02 | 6.61 | 6.66 |
| *CHST15* | 8.89E-05 | 8.31 | 8.04 | 2.13E-01 | 9.73 | 9.63 |
| *SKIL* | 1.90E-02 | 2.13 | 2.53 | 8.60E-03 | 6.59 | 6.62 |
| *SYNE2* | 1.97E-01 | 2.52 | 2.87 | 5.81E-01 | 6.51 | 6.52 |
| *LIMD2* | 2.29E-02 | 2.76 | 3.04 | 4.57E-02 | 6.59 | 6.54 |

| | | | | | | |
|---|---|---|---|---|---|---|
| *MCF2L* | 4.31E-04 | 1.75 | 2.40 | 1.59E-03 | 8.58 | 8.79 |
| *TMEM156* | 2.43E-04 | 6.99 | 7.47 | 2.22E-04 | 8.48 | 9.03 |
| *KCNJ3* | 4.41E-03 | 1.40 | 2.09 | 3.47E-02 | 6.57 | 6.53 |
| *A2M* | 4.23E-01 | 8.54 | 8.66 | 2.57E-03 | 10.30 | 10.67 |
| *ADAP2* | 5.13E-01 | 7.00 | 7.04 | 9.48E-04 | 7.73 | 8.05 |
| *ADCY7* | 3.02E-01 | 4.75 | 4.83 | 2.83E-04 | 7.22 | 7.41 |
| *ADPGK* | 2.24E-01 | 6.14 | 6.07 | 2.04E-03 | 7.87 | 7.96 |
| *ALPK1* | 9.64E-02 | 4.68 | 4.84 | 2.20E-05 | 6.88 | 7.08 |
| *AOAH* | 1.43E-01 | 2.65 | 2.91 | 3.36E-06 | 6.72 | 6.88 |
| *APBB1IP* | 6.76E-02 | 5.32 | 5.52 | 4.14E-02 | 8.30 | 8.54 |
| *ARHGAP10* | 5.35E-02 | 5.54 | 5.70 | 4.04E-04 | 7.24 | 7.43 |
| *ARHGAP15* | 2.65E-01 | 3.50 | 3.65 | 1.99E-04 | 7.00 | 7.19 |
| *B3GNT5* | 1.62E-01 | 2.40 | 2.64 | 2.85E-02 | 6.52 | 6.57 |
| *BAIAP2L1* | 8.46E-02 | 1.93 | 2.25 | 3.88E-03 | 6.56 | 6.63 |
| *BLM* | 3.29E-01 | 3.29 | 3.42 | 1.57E-03 | 6.91 | 7.06 |
| *BNC2* | NA | NA | NA | 3.32E-05 | 6.63 | 6.76 |
| *C10orf11* | 5.05E-02 | 3.29 | 3.57 | 2.52E-05 | 6.80 | 6.93 |
| *CCDC50* | 3.61E-01 | 3.45 | 3.33 | 9.83E-03 | 7.46 | 7.66 |
| *CD53* | 4.09E-01 | 6.56 | 6.48 | 4.96E-02 | 7.21 | 7.35 |
| *COBLL1* | 8.58E-01 | 7.28 | 7.27 | 2.05E-03 | 9.35 | 9.66 |
| *COL11A1* | 1.40E-01 | 1.45 | 2.05 | 6.12E-05 | 7.09 | 7.28 |
| *COL20A1* | 2.67E-01 | 6.83 | 6.93 | 3.27E-02 | 7.56 | 7.79 |
| *CREB3L2* | 1.26E-01 | 1.22 | 1.75 | 7.25E-03 | 8.38 | 8.60 |
| *CRTAP* | 4.32E-01 | 4.57 | 4.49 | 1.32E-04 | 7.20 | 7.35 |
| *CSF3R* | 1.22E-01 | 6.30 | 6.45 | 5.35E-05 | 7.24 | 7.63 |
| *CSPG4* | 1.60E-01 | 5.75 | 5.91 | 6.72E-04 | 7.68 | 8.16 |
| *CTSB* | 1.77E-03 | 9.17 | 9.01 | 1.13E-04 | 10.81 | 11.11 |
| *DIAPH2* | 4.59E-10 | 7.36 | 6.77 | 4.22E-02 | 7.33 | 7.46 |
| *DISC1* | 6.16E-02 | 6.02 | 6.15 | 6.35E-05 | 6.56 | 6.64 |
| *DOCK8* | 9.14E-01 | 3.56 | 3.58 | 8.96E-03 | 7.37 | 7.68 |
| *EDA* | 7.60E-01 | 2.46 | 2.53 | 4.92E-04 | 6.48 | 6.51 |
| *EIF2B5* | 8.49E-02 | 7.06 | 7.14 | 3.56E-03 | 8.54 | 8.77 |
| *EPAS1* | 1.11E-01 | 1.46 | 1.90 | 7.25E-03 | 10.04 | 10.34 |
| *FAM105A* | 2.85E-01 | 4.45 | 4.55 | 4.56E-03 | 6.91 | 7.00 |
| *FAM160A1* | NA | NA | NA | 1.30E-03 | 6.56 | 6.63 |
| *FCGR2A* | 1.20E-01 | 4.86 | 5.06 | 5.26E-03 | 6.95 | 7.19 |
| *FGD2* | 5.18E-01 | 1.72 | 1.93 | 1.59E-04 | 7.26 | 7.62 |
| *FLI1* | 1.12E-01 | 5.43 | 5.61 | 4.50E-05 | 7.31 | 7.64 |
| *FMNL3* | 2.57E-01 | 2.13 | 1.91 | 2.35E-02 | 6.81 | 6.88 |
| *FOS* | 1.10E-01 | 7.32 | 7.53 | 7.28E-03 | 8.64 | 9.21 |
| *GMFG* | 3.23E-01 | 6.17 | 6.08 | 2.93E-05 | 7.75 | 8.06 |
| *GNS* | 8.42E-01 | 1.32 | 1.19 | 8.91E-03 | 10.74 | 10.89 |
| *GPNMB* | 7.06E-01 | 6.75 | 6.71 | 4.55E-02 | 8.11 | 8.42 |
| *HLA-DRA* | 1.31E-01 | 6.41 | 6.59 | 1.81E-02 | 9.34 | 9.83 |
| *ICA1* | 2.75E-16 | 7.78 | 7.13 | 3.22E-04 | 8.77 | 9.12 |
| *INTS8* | 5.04E-02 | 5.79 | 5.90 | 4.71E-02 | 8.26 | 8.37 |

| | | | | | | |
|---|---|---|---|---|---|---|
| *IPO8* | 3.79E-01 | 3.67 | 3.76 | 4.57E-02 | 7.90 | 8.09 |
| *IRAK2* | 7.50E-01 | 1.37 | 1.19 | 2.28E-05 | 7.49 | 7.79 |
| *ITGAX* | 5.06E-02 | 5.84 | 6.00 | 8.24E-04 | 6.65 | 6.75 |
| *KAT2A* | 3.98E-01 | 7.86 | 7.82 | 1.14E-05 | 10.24 | 10.60 |
| *KDELR1* | 2.01E-01 | 6.49 | 6.44 | 4.83E-02 | 8.80 | 8.91 |
| *LY86* | 9.34E-01 | 6.72 | 6.73 | 1.45E-03 | 7.40 | 7.68 |
| *LYPD1* | 4.47E-01 | 3.91 | 3.82 | 4.94E-02 | 8.49 | 8.73 |
| *MGAT5* | 8.48E-01 | 2.13 | 2.20 | 2.13E-02 | 6.42 | 6.45 |
| *MLST8* | 2.19E-01 | 5.84 | 5.91 | 1.92E-02 | 7.93 | 8.07 |
| *MLXIPL* | NA | NA | NA | 3.25E-04 | 7.59 | 7.91 |
| *MPZL1* | 7.63E-01 | 4.17 | 4.19 | 4.49E-02 | 7.13 | 7.21 |
| *NCK2* | 2.58E-01 | 1.38 | 1.71 | 2.77E-02 | 9.17 | 9.35 |
| *OLIG2* | 6.85E-01 | 9.25 | 9.28 | 1.42E-02 | 9.59 | 9.89 |
| *PDGFRA* | 1.98E-04 | 5.98 | 5.62 | 1.38E-02 | 8.67 | 8.99 |
| *PIK3AP1* | 1.49E-01 | 2.16 | 2.43 | 6.06E-05 | 6.79 | 7.01 |
| *PIK3CA* | 8.57E-01 | 4.45 | 4.47 | 2.75E-02 | 7.69 | 7.81 |
| *PLCE1* | 9.56E-02 | 5.73 | 5.92 | 9.76E-09 | 7.54 | 7.98 |
| *PLCG2* | 2.18E-01 | 3.30 | 3.49 | 1.31E-05 | 7.36 | 7.71 |
| *PRRX1* | 6.66E-01 | 2.99 | 3.06 | 1.51E-05 | 7.24 | 7.55 |
| *PXDN* | 2.06E-04 | 6.91 | 6.72 | 9.49E-03 | 7.26 | 7.38 |
| *RCSD1* | 8.89E-01 | 5.40 | 5.41 | 1.50E-02 | 6.88 | 7.01 |
| *RGS1* | 1.08E-01 | 5.76 | 6.07 | 9.09E-03 | 7.50 | 8.13 |
| *RGS10* | 8.32E-01 | 5.63 | 5.64 | 8.73E-03 | 7.38 | 7.56 |
| *ROCK1* | 7.93E-02 | 2.45 | 2.82 | 4.76E-03 | 6.83 | 7.00 |
| *RPS6KA3* | 1.27E-04 | 4.02 | 3.58 | 1.09E-02 | 6.88 | 6.94 |
| *S100Z* | 5.66E-01 | 2.47 | 2.57 | 4.45E-02 | 6.49 | 6.53 |
| *SH3RF3* | 2.38E-01 | 1.54 | 1.83 | 3.65E-03 | 7.29 | 7.44 |
| *SLC2A5* | 4.43E-01 | 7.04 | 7.11 | 1.78E-02 | 8.36 | 8.73 |
| *SLC9A9* | 4.22E-01 | 4.30 | 4.38 | 7.20E-04 | 7.83 | 8.09 |
| *SNAP23* | 1.16E-01 | 5.93 | 6.16 | 2.32E-05 | 7.19 | 7.41 |
| *SSTR2* | 1.05E-01 | 3.84 | 4.14 | 5.87E-04 | 9.60 | 9.94 |
| *STK32B* | 5.24E-01 | 5.09 | 5.04 | 1.59E-04 | 7.28 | 7.55 |
| *TGFBR1* | 8.71E-01 | 1.57 | 1.66 | 6.29E-04 | 6.59 | 6.67 |
| *TGFBR2* | NA | 0.82 | 1.14 | 1.57E-03 | 8.17 | 8.60 |
| *TRPM2* | 5.75E-01 | 3.12 | 3.03 | 9.18E-03 | 7.39 | 7.59 |
| *TSEN34* | 2.34E-01 | 6.07 | 6.00 | 1.21E-02 | 9.20 | 9.32 |
| *USP24* | NA | NA | NA | 7.34E-03 | 8.92 | 9.06 |
| *WDFY4* | NA | NA | NA | 8.41E-03 | 6.57 | 6.61 |
| *WIPF1* | 8.36E-01 | 7.65 | 7.68 | 4.26E-02 | 6.78 | 6.86 |
| *ZC3HAV1* | 4.65E-01 | 6.46 | 6.50 | 3.01E-04 | 7.32 | 7.53 |
| *ZNF493* | 7.87E-01 | 1.57 | 1.47 | 1.14E-02 | 7.88 | 8.03 |
| *ZYX* | 7.04E-02 | 8.16 | 8.24 | 2.03E-02 | 8.41 | 8.53 |

Note: We chose top-ranked 1,000 risk genes for AD from the genetic co-expression analysis to perform differential gene expression analyses based on two bulk-based transcriptomic profiles (GSE109887, n = 78; GSE15222, n = 363). We found that 421 genes showed significantly positive associations with AD (42.1%) among two independent bulk-based transcriptomic datasets. We used two-sided Student’s T test to assess the significance for each gene. NA: Not applicable.
